# Genome-wide multi-trait analysis of irritable bowel syndrome and related mental conditions identifies 38 new genetic hits

**DOI:** 10.1101/2023.01.30.23285080

**Authors:** Silvia Alemany, María Soler Artigas, Judit Cabana, Dana Salim Fakhreddine, Natalia Llonga, Laura Vilar-Ribó, Amanda Rodríguez-Urrutia, Judit Palacio, Ana María González-Castro, Beatriz Lobo, Carmen Alonso-Cotoner, Magnus Simrén, Javier Santos, Josep Antoni Ramos-Quiroga, Marta Ribasés

**Author notes:** **Corresponding authors:** Marta Ribasés and Silvia Alemany. Psychiatric Genetics Unit, Group of Psychiatry Mental Health and Addiction, Vall d’Hebron Research Institute (VHIR), Universitat Autònoma de Barcelona, Barcelona Spain. Passeig Vall d’Hebron, 119-129, 08035 Barcelona, Spain.

## Abstract

**Background:** Irritable bowel syndrome (IBS) is a chronic disorder of gut-brain interaction frequently accompanied by mental conditions, including depression and anxiety. Despite showing substantial heritability and being partly determined by a genetic component, the genetic underpinnings explaining the high rates of comorbidity remain largely unclear and there are no conclusive data on the temporal relationship between them. Exploring the overlapping genetic architecture between IBS and mental conditions may help to identify novel genetic loci and biological mechanisms underlying IBS and causal relationships between them.

**Methods:** We quantified the genetic overlap between IBS, neuroticism, depression and anxiety, conducted a multi-trait genome-wide association study (GWAS) considering these traits and investigated causal relationships between them by using the largest GWAS to date.

**Results:** IBS showed to be a highly polygenic disorder with extensive genetic sharing with mental conditions. Multi-trait analysis of IBS and neuroticism, depression and anxiety identified 42 genome-wide significant hits for IBS, of which 38 are novel. Fine-mapping risk loci highlighted 289 genes upregulated during early embryonic brain development and gene-sets related with psychiatric, digestive and autoimmune disorders. IBS-associated genes were enriched for target genes of anti-inflammatory and antirheumatic drugs, anesthetics and opioid dependence pharmacological treatment. Mendelian-randomization analysis accounting for correlated pleiotropy identified bidirectional causal effects between IBS and neuroticism and depression and causal effects of the genetic liability of IBS on anxiety.

**Conclusions:** These findings provide evidence of the polygenic architecture of IBS, identify novel hits for IBS and extend previous knowledge on the genetic overlap and relationship between gastrointestinal and mental disorders.

## INTRODUCTION

Irritable bowel syndrome (IBS) is one of the most prevalent disorders of gut-brain interaction with a population lifetime risk of 11% ^1^ and a point prevalence of 4.1% according to the strict Rome IV criteria ^2^. IBS research is extremely challenging due to the multifactorial etiology of the disease and the heterogeneity of patients, who present high comorbidity rates for mental disorders, particularly, anxiety and depression, which impacts negatively on the patients’ quality of life ^1,3,4^.

A recent systematic review revealed that the prevalence of anxiety and depression symptoms among IBS patients is 39.1% and 28.8%, respectively ^5^. In addition, IBS has been associated with more severe depressive symptoms compared to healthy controls and, when co-existing with psychiatric disorders, gastrointestinal symptoms are more severe and disabling ^6–11^. This close association between IBS, anxiety and depression is also supported by neuroimaging studies and might be related to the bi-directional communication between the brain and the digestive system, termed the brain-gut-axis, which occurs through microbiota, neural, neuroimmune and neuroendocrine pathways ^12–14^. This idea agrees with evidence indicating that psychiatric interventions, including antidepressants or cognitive-behavioral therapy, improve IBS patients functioning and suggests that common pathophysiological mechanisms may be underlying these conditions ^15^.

IBS, anxiety and depression are partly determined by a genetic component and show substantial heritability ranging from 6% for IBS to 30%-50% for anxiety and depression ^16–18^. The largest GWAS on IBS conducted to date included 53,400 cases and 433,201 controls and identified six genome-wide significant loci ^18^ which represents an improvement over the previous study identifying four independent hits ^19^. Interestingly, among 173 traits, three mental conditions (neuroticism, depression and anxiety) were the most genetically correlated traits with IBS ^18^. Despite these strong genetic correlations, the genetic underpinnings explaining the high rates of comorbidity between IBS and mental conditions remain largely unclear and there are no conclusive data on the temporal and causal relationship between them ^18,19^.

In the present study we investigated the shared genetic architecture and the nature of the relationship between IBS and three highly genetically correlated conditions (neuroticism, depression and anxiety) using summary statistics of the largest GWAS datasets available so far by (i) estimating the genetic correlation and overlap between them, (ii) conducting a Multi-Trait Analysis of GWAS (MTAG) to identify novel genetic loci for IBS and (iii) performing downstream analyses to explore the overlaping genetic basis with other disorders and traits as well as causal relationships between them.

## MATERIALS AND METHODS

### Samples

We used publicly available SNP-level GWAS summary statistics for IBS ^18^, neuroticism ^20^, depression ^21^ and anxiety (Table 1). For further details see Supplementary Note 1.

**Table 1.** Summary of the GWAS datasets used in the current study.

| Phenotype | N cases | N controls | N total | N effective <sup>a</sup> | GWAS Hits<sup>b</sup> | Reference |
| --- | --- | --- | --- | --- | --- | --- |
| **IBS** | 53,400 | 433,201 | 486,601 | 190,159 | 6 | 18 |
| **Neuroticism** | - | - | □390,278<sup>c</sup> | □390,278 | 136<sup>d</sup> | 20 |
| **Depression** | 170,756 | 329,443 | 500,199<sup>c</sup> | 449,856 | 102<sup>d</sup> | 21 |
| **Anxiety nerves or GAD** | 16,730 | 101,021 | 117,751 | 57,412 | 1 | UKBB phenotype code: 20544_15 |

NOTE: GAD generalized anxiety disorder; UKBB UK Biobank.
<sup>a</sup> N effective sample sizes were calculated following the equation: $N_{eff}=4/(1/N_{cases}+1/N_{controls})$ .
<sup>b</sup> Number of genome-wide significant independent loci.
<sup>c</sup> Sample size excluding the 23andMe cohort.
<sup>d</sup> Hits including the 23andMe cohort.

### SNP-based heritability genetic correlation and overlap

SNP heritability (*h*^*2*^_SNP_) and pair-wise genetic correlation between IBS and each mental condition was calculated using linkage disequilibrium score regression (LDSC) analysis ^22^. Conversion of *h*^*2*^_SNP_ estimates from observed to liability scale was done using a population prevalence of 11%, 25%, 30% and 14% for IBS, neuroticism, depression and anxiety, respectively. Polygenic overlap between IBS and each mental condition was quantified using MiXeR ^23^. MiXeR caclulates the number of trait-influencing loci for each trait (univariate model) and for both traits (bivariate model) and the proportion of variants with concordant direction of effects for both traits. The proportion of SNPs shared by two traits is indicated by the Dice coefficient. Model fit was assessed using the Akaike Information Criterion (AIC). For further details see Supplementary Note 2.

### Multi-Trait Analysis of GWAS (MTAG)

To identify new loci for IBS, SNP-level GWAS for IBS, neuroticism, depression and anxiety were meta-analyzed using MTAG ^24^. To discard inflation in our results we calculated the max-false discovery rate (max-FDR) using the default settings as previously described ^24,25^. Independent lead SNPs from MTAG-IBS results (*P*-value<5-E08) were identified through clumping (r2=0.05, kb=5000) using the 1000 Genomes Project Phase 3 European reference panel (http://www.internationalgenome.org/) and PLINK1.09 as described by Eijsbouts et al. ^18^. We carried out conditional analyses to evaluate independence between secondary (within 5000kb and r2<0.2) and index variants within each locus using COJO implemented in Genome-wide Complex Trait Analysis (GCTA) ^26^. For further details on conditional analysis see Supplementary Note 3.

### Credible variants and functional annotation

Sets of credible variants (credible-sets) were identified by fine-mapping the independent lead SNPs of MTAG-IBS using three different tools, FINEMAP 1.3.1 ^27^, PAINTOR v3.0 ^28^ and CAVIARBF v.0.2.1 ^29^ following the pipeline available elsewhere ^30^. Variants located in a region of 5000 kb around the lead SNPs were included in the analysis and we assumed that there was only one causal variant per locus. We used the recommended parameters of each tool and only variants identified by all three methods were considered. Functional annotation of the credible variants was conducted using FUMA^31^. For further details see Supplementary Note 4.

### Gene-based and gene-set analyses of MTAG-IBS results

Gene-based and gene-set analyses of MTAG-IBS risk loci were performed using MAGMA v1.08 ^32^ implemented in FUMA ^31^. Tissue specific gene expression was explored using MAGMA gene-property analysis of expression data from GTEx v8 and BrainSpan available in FUMA (databases detailed in Supplementary Note 5). All gene sets were obtained from the Molecular Signatures Database (MSigDB v6.2) and included GO, KEGG, BIOCARTA and Reactome representing a total of 11,960 gene sets. The Bonferroni-corrected significance threshold was 0.05/11960 gene sets=4.18E-06.

### Drug target identification

To explore whether finemapped genes related with IBS were enriched for target genes of drugs (druggable genes) we performed enrichment analysis based on information from the PharmGKB using WebGestAlt ^33^. Identified drugs were classified according to available information from the Anatomical Therapeutic Chemical (ATC) classification system.

### Partitioned heritability and genetic correlations

We partitioned *h*^*2*^_*SNP*_ of MTAG-IBS results by functional annotation categories using stratified LDSC ^34^. We calculated whether any of the 28 specific genomic categories included in the analysis was enriched for variants that contribute to *h*^*2*^_SNP_. Annotations for these functional genomic categories (e.g. coding or regulatory regions) were obtained from LDSC website (https://github.com/bulik/ldsc/wiki/Partitioned-Heritability). We focused on categories extended by 500bp in either direction. Enrichment/depletion of heritability in each category is calculated as the proportion of heritability attributable to SNPs in the specified category divided by the proportion of total SNPs annotated to that category. The Bonferroni-corrected significance threshold was 0.05/28 annotations=0.0021.

We explored genetic correlations between our MTAG-IBS results and gastrointestinal, immunological and psychiatric disorders using LDSC analysis ^22^. We selected all GWAS summary statistics of gastrointestinal/abdominal, immunological/systemic (UK Biobank: 21 phenotypes) and psychiatric disorders (PGC: 7 phenotypes) available in the MR-Base database (Supplementary Table 14) ^35^. We used GWAS summary statistics including both males and females of European ancestry. If several GWAS were available for the same disorder, we chose the study with the largest effective sample size (N effective = 4NcaNco/(Nca+Nco)). The Bonferroni-corrected significance threshold used was 0.05/28 traits=0.0018.

### Causal analysis using summary effect estimates (CAUSE)

Causal relationships between IBS and correlated traits were assessed considering independent variants (r2=0.05; kb=5000) associated with the exposure with *P*<1.0E-03 using CAUSE ^36^. Bidirectional relationships were tested considering IBS as exposure and depression, anxiety or neuroticism as outcomes and vice-versa. Given that SE was not available from the largest study on neuroticism to date 37, we used the GWAS dataset on neuroticism by Luciano et al. in 329,821 subjects as an alternative 38. The strengths of CAUSE involve accounting for correlated horizontal pleiotropic effects (i.e. when a variant affects the outcome and the mediator through shared heritable factors) and using a less stringent significance threshold (P<1.0E-3) allowing the incorporation of more variants to the analyses. CAUSE compares two nested models, a sharing and a causal model. Both models allow for horizontal pleiotropy (correlated pleiotropy (eta)) but only the casual model includes a causal effect parameter (gamma). The sharing and the causal model are compared against a null model and against each other. Model comparisons are carried out using the expected log pointwise posterior density (ELPD), a Bayesian model comparison approach that estimates how well the posterior distributions of a particular model are expected to predict a new set data. When P <0.05 the second model fits the data better than the first model. There is evidence of causal effects when the causal model represents a significant improvement in the model fit of the sharing model. For further details see Supplementary Note 9.

## RESULTS

### SNP-based heritability, genetic correlation and overlap

The latest GWAS on IBS ^18^, neuroticism ^20^, depression ^21^ and anxiety used herein are summarized in Table 1 and Supplementary Note 1. The estimated SNP heritability (*h*^*2*^_SNP_) was 6.9% (SE=0.004) for IBS, 14.6% (SE=0.005) for neuroticism, 9.9% (SE=0.004) for depression and 8.3% (SE=0.011) for anxiety (Table 2). We found evidence of strong genetic correlation between IBS and all three mental conditions, ranging from 53% to 68% (Table 2). Univariate MiXeR analysis revealed that IBS and neuroticism were highly polygenic, with around twelve thousand variants explaining 90% of SNP heritability (12,438 variants for IBS and 12,308 for neuroticism; Supplementary Table 1a). Bivariate MiXeR analysis showed that the majority of the variants influencing IBS were shared with neuroticism (10,793 (SE=1.094) out of 12,438 (SE=1.305) variants, Dice coefficient=0.87), with a high proportion of variants being concordant (71%) (Supplementary Table 1a and Supplementary Figure 1). Unfortunately, MiXeR was unable to accurately quantify the genetic overlap between IBS and depression or anxiety according to the Akaike Information Criterion (AIC; Supplementary Table 1b).

**Table 2.** Genetic correlation estimates for IBS and neuroticism, depression and anxiety using Linkage Disequilibrium Score Regression (LDSC).

| Trait 1 | Trait 2 | Genetic<br>Correlation | SE | Z | P-value | Intercept (SE) | Trait 1 | Trait 2 |
| --- | --- | --- | --- | --- | --- | --- | --- | --- |
| | | | | | | | $h^2$ (SE) | $h^2$ (SE) |
| IBS | Neuroticism | 0.526 | 0.027 | 19.298 | 5.54E-83 | 1.013 (0.013) | 0.069 (0.004) | 0.146 (0.005) |
| IBS | Depression | 0.587 | 0.026 | 22.714 | 3.23E-114 | 0.992 (0.01) | 0.069 (0.004) | 0.099 (0.004) |
| IBS | Anxiety | 0.677 | 0.065 | 10.360 | 3.75E-25 | 0.999 (0.74) | 0.068 (0.004) | 0.083 (0.011) |

NOTE: SE, standard error; $h^2$ , heritability.

### Multi-Trait Analysis of GWAS (MTAG)

To identify novel loci for IBS, we combined the summary statistics from the GWAS on IBS, neuroticism, depression and anxiety using MTAG, increasing the estimated effective sample size from 486,601 in the original IBS dataset to 887,490. The max-FDR of MTAG-IBS analysis was low (0.020) suggesting no inflation, consistent with the similar mean chi-square values for the different GWAS, ranging from 1.08 for anxiety to 1.69 for neuroticism. After MTAG analysis, the number of genome-wide significant SNPs for IBS increased from six in the original GWAS to 42 independent SNPs in 37 loci in the current study (Figure 1, Supplementary Figure 2, Table 3 and Supplementary Table 2). Comparing these results with the ones originally described for IBS ^18^, 38 out of the 42 SNPs identified herein were novel for IBS and all of them showed consistent direction of the association (Figure 1a and Supplementary Table 3). Of them, 11 were not previously associated with neuroticism, depression or anxiety (Figure 1d). The remaining signals, 27 in total, were novel risk loci for IBS but previously reported for neuroticism and/or depression (Table 3, Figure 1d) and overall showed consistent direction of association with that reported in the original studies (Figure 1a). Of the six loci previously identified in IBS ^18^, four of them, on chromosome 3, 6, 9 and 11, were among the significant loci for IBS in the current study and the two additional ones, in chromosome 13, showed suggestive evidence of association (*P*<5E-07; Table 3). Among top findings, we found lead SNPs nearby genes involved in transcriptional regulation, including non-coding RNAs (*RP11*-*629G13*.*1* and *MSH5-SAPCD1*), RNA splicing (*CELF4*), chromatin remodeling (*EP300* and *HIST1H3J*), mRNA transport (*FAM120A*) or nucleic acid binding (*TCF4* and *ELAVL2*), as well as in brain development (*TMEM161B*) or presynaptic activity (*PCLO*).

**Figure 1.**
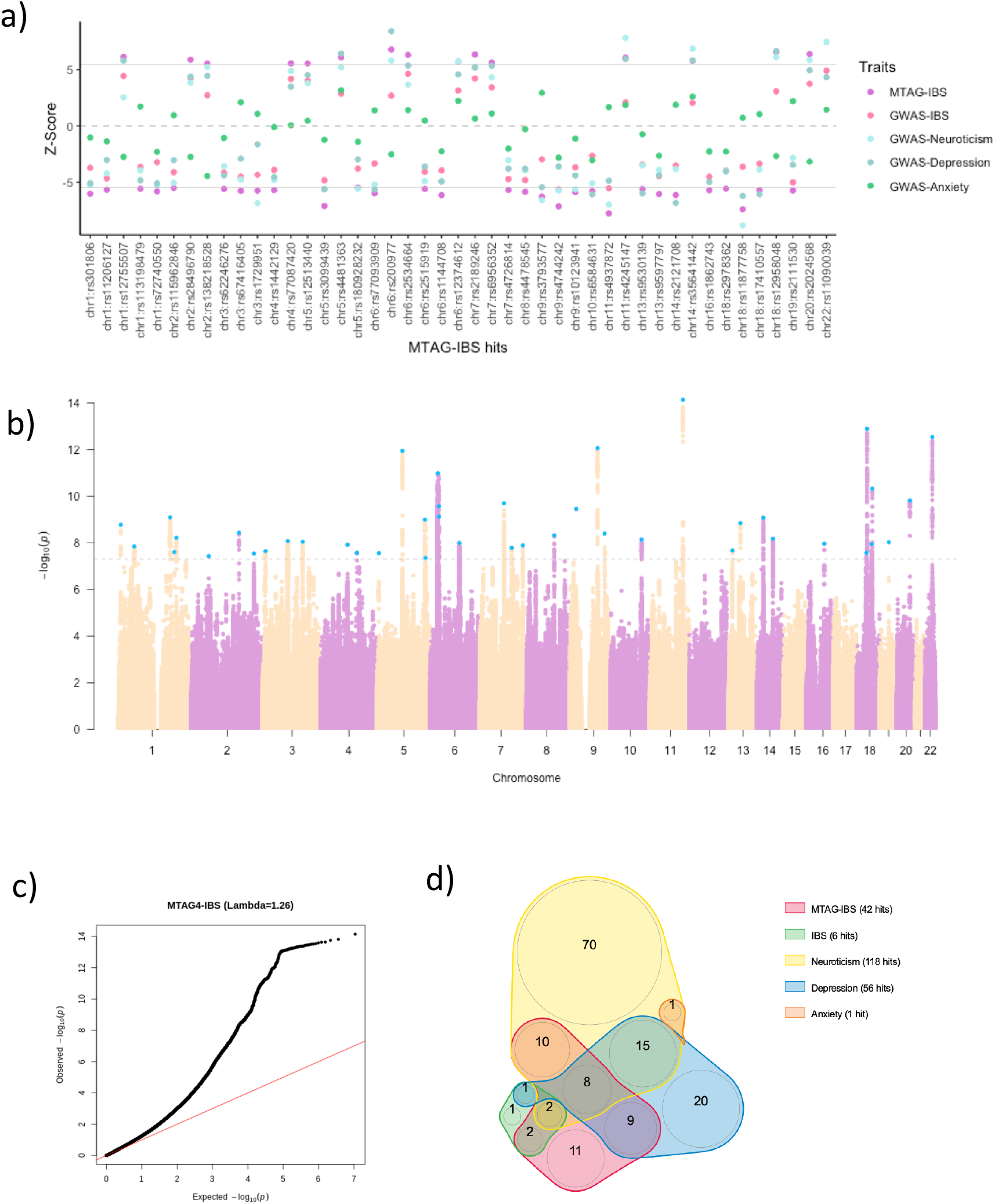
MTAG results of IBS and overlap with previous GWAS on IBS, neuroticism, depression and anxiety. a) Z-scores of MTAG-IBS and original GWAS on IBS, neuroticism, depression and anxiety for each of the independent lead SNPs (n=42) found in MTAG-IBS results. Dotted grey line indicates 0 Z-score and solid grey lines indicate statistical significance at P<5-E08. b) Manhattan plot of the MTAG-IBS results. Dotted grey line indicates statistical significance at P<5-E08. c) QQ plot of the MTAG-IBS results. d) Venn diagram depicting overlap among MTAG-IBS independent lead SNPs and genome-wide significant hits in the original GWAS.

**Table 3.**
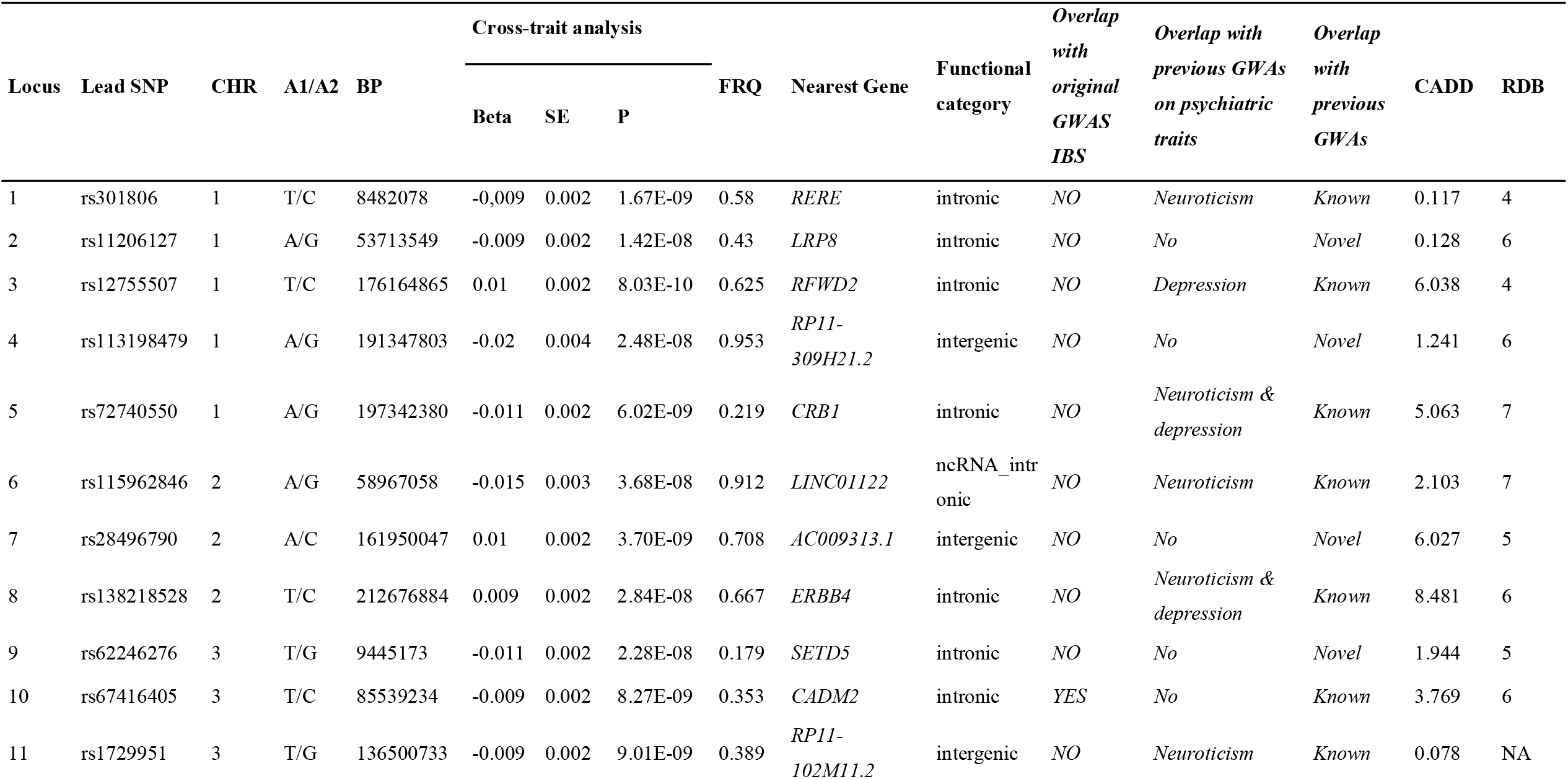
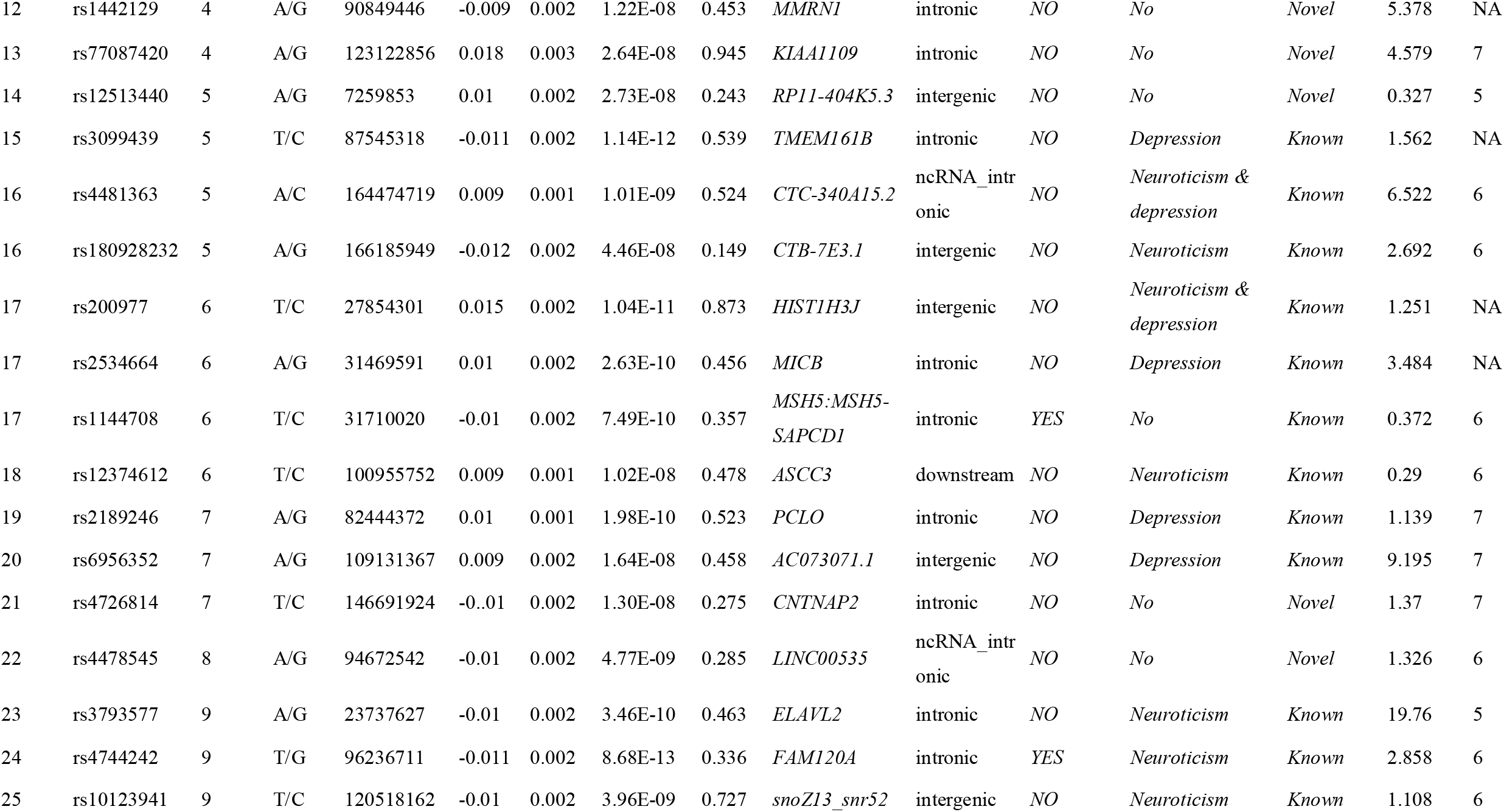
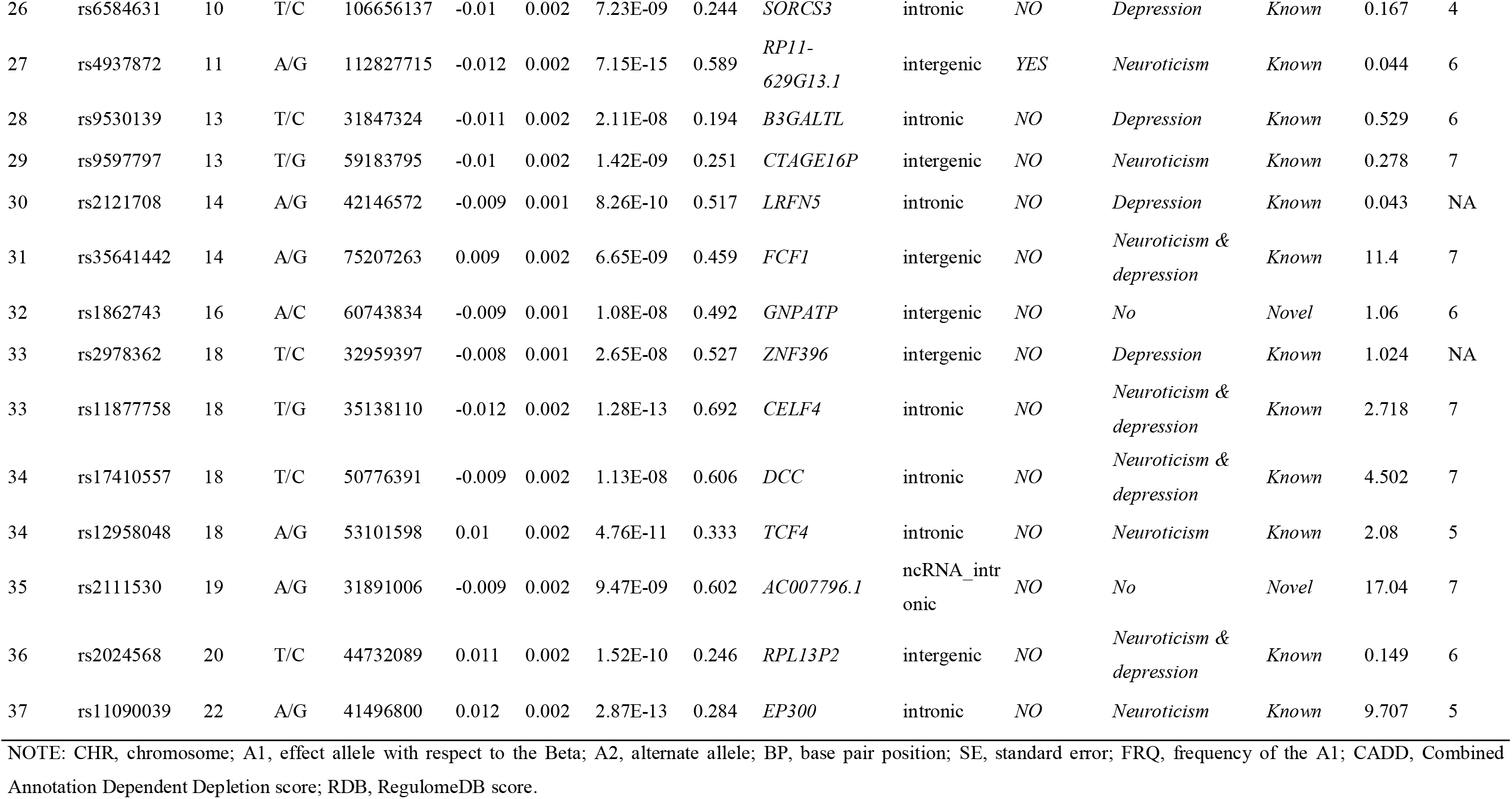
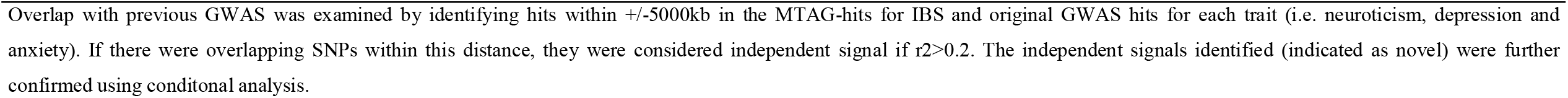
Results for the 42 independent lead SNPs identified in the MTAG-IBS analysis.

### Credible variants and functional annotation

We identified a total of 1,818 Bayesian credible variants in the 37 independent loci for IBS (Supplementary Table 4). Their functional annotation revealed over-presentation of SNPs in introns (64.6%), intergenic regions (21.7%) or located in non-coding RNA (9.4%) (Figure 2 and Supplementary Table 5). A total of 75% of the variants within credible sets were located in open chromatin regions (minimum chromatin state ≤ 7), 3% were likely to affect the binding of transcription factors (RegulomeDB scores from 1b to 2c) and 0.05% may be deleterious (Combined Annotation Dependent Depletion (CADD) score > 12.37) (Figure 2 and Supplementary Table 5). Forty-eight variants were previously related by GWAS (*P*<5E-07) to digestive-related phenotypes (e.g. inflammatory bowel disease, gastroesophageal reflux or gut microbiota relative abundance), lifestyle factors (e.g. alcohol consumption, lifetime smoking, coffee consumption or moderate to vigorous physical activity levels) and brain and neuropsychiatric phenotypes (e.g. neuroticism, depression, anxiety, cognition or brain morphology) (Supplementary Table 6). In addition, we found that more that half of the credible variants (n=953; 52%) were expression quantitative trait loci (eQTL) for at least one gene in one brain area (n=895; 49%) and/or digestive tissue (n=690; 38%; Supplementary Table 7).

**Figure 2.**
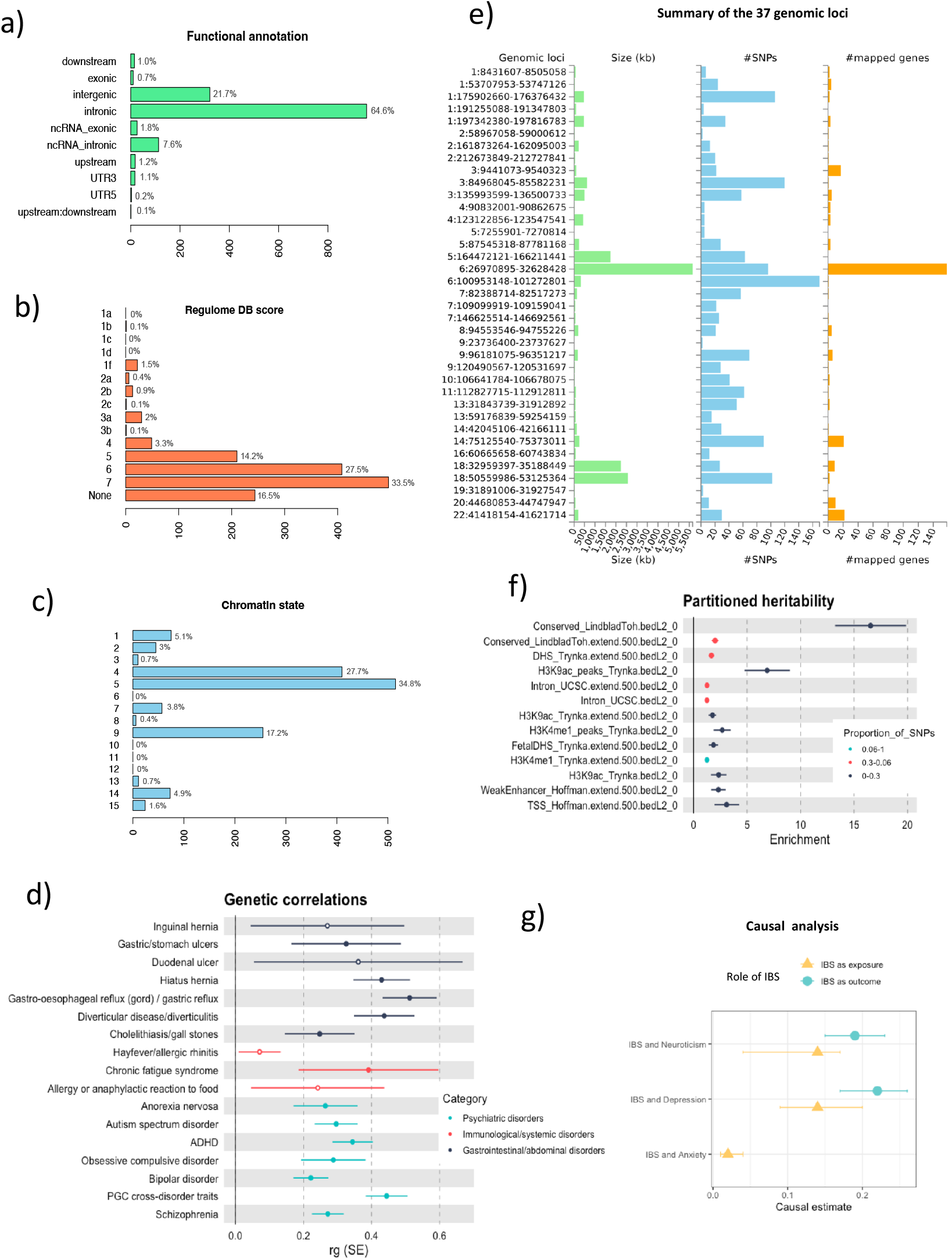
Follow-up analysis of MTAG-IBS results and causal analysis. a) Functional annotation of the credible variants associated with MTAG-IBS. b) RegulomeDB scores of the credible variants associated with MTAG-IBS. Low scores indicate increasing likelihood of having regulatory function. c) Distribution of the credible variants associated with MTAG-IBS across 15 categories of minimum chromatin state. Lower state indicating higher accessibility and states from 1 to 7 refer to open chromatin states. d) Genetic correlations (rg) between MTAG-IBS results and 17 phenotypes involving digestive, immunological and psychiatric disorders. Only significant correlations after Bonferroni correction are displayed. e) Bar graphs depicting the size of the genomic locus (left), number of candidate SNPs in the locus (center) and number of mapped genes in the genomic locus (right). Genomic loci are displayed by “chromosome: start position-end position “. f) Partitioning of the SNP heritability of the MTAG-IBS results using LD Score regression. Enrichment was calculated by dividing the partial heritability of a category by the proportion of SNPs in that category (proportion indicated by color). Only significant enrichments are displayed. g) Causal relationships between IBS and neuroticism, depression and anxiety assessed using Causal Analysis Using Summary Effect estimates (CAUSE). Only associations with evidence of causal relationship are displayed.

Credible variants were mapped to 289 unique genes (Supplementary Table 8) that were significantly enriched in genes upregulated during early embryonic brain development (8th post conceptual week; Supplementary Figure 3) and in several gene-sets (Supplementary Table 9). Among the most significant ones, we found psychiatric disorders (GWAS catalog: autism spectrum disorder or schizophrenia, *P*-adjusted=4.96E-193), digestive disorders (GWAS catalog: ulcerative colitis, *P*-adjusted=1.13E-57 and inflammatory bowel disease, *P*-adjusted=7.05E-40), autoimmune disease (KEGG: Systemic lupus erythematosus, *P*-adjusted=7.91E-61) and histone deacetylases (Reactome: HDACS deacetylate histones, *P*-adjusted=3.09E-46) (Supplementary Table 9).

### Gene-based and gene-set analyses of MTAG-IBS risk loci

The gene-based analysis identified 76 significant genes, which were associated with expression changes in the cerebellum (*P*=5.2E-09), frontal cortex (*P*=9.8E-07), anterior cingulate cortex (*P*=1.8E-05), basal ganglia nuclei (nucleus accumbens: *P*=6.9E-05; caudate: *P*=9.7E-04) and hypothalamus (*P*=4.3E-04) (Supplementary Table 10, Supplementary Figure 4) as well as with gene expression during the 21^st^ post conceptual week (*P*=8.5E-04) (Supplementary Figure 5). Among top findings, we found genes with a role in brain development and synaptic function, including *CADM2* and *NCAM1*, previously identified in the latest GWAS on IBS, and also genes involved in transcriptional regulation through mRNA transport or chromatin structure, including *FAM120A, PHF2* and different histone coding genes. When we conducted the gene-set analysis we found the *branching morphogenesis of a nerve* pathway significantly associated with IBS (gene-set size=10 genes; *P*= 1.7E-06) (Supplementary Table 11).

### Drug target identification

The enrichment analysis on druggable genes showed enrichment of MTAG-IBS-finemapped credible genes in druggable genes for 21 drugs (Supplementary Table 12), being l-lysine (*P* < 2.2E-16), belinostat (*P*=8.6E-10), s-adenosylmethionine (*P*=7.0E-09) and allopurinol (*P*=1.5E-07), the top ones (Supplementary Table 12). They also included drugs related to musculo-skeletal system, such as anti-inflammatory and antirheumatic drugs, or related to the nervous system, such as anesthetics and drugs used in opioid dependence (Supplementary Table 12).

### Partitioned heritability and genetic correlations

SNP When we partitioned the *h*^*2*^ of IBS, we observed significant heritability enrichment in seven functional categories (Figure 2 and Supplementary Table 13), with the strongest enrichment of variants in conserved regions (enrichment=2.01; *P*=4.0E-09), DNase I hypersensitive sites (DHSs) regions (enrichment=1.66; *P*=9.1E-08) and histone H3 lysine 9 acetylation (H3K9ac) peaks (enrichment=6.88; *P*=1.1E-07).

We found significant genetic correlations between IBS and 13 gastrointestinal, immunological or psychiatric disorders using GWAS summary statistics available in the MR-Base database ^35^, including gastric reflux (rg=0.51; *P*=2.6E-36), the cross-disorder GWAS from the PGC involving schizophrenia, bipolar disorder, major depressive disorder, autism spectrum disorders (ASD) and attention-deficit/hyperactivity disorder (ADHD) (rg=0.44, *P*=9.7E-46), diverticulitis (rg=0.44, *P*=7.4E-22), hiatus hernia (rg=0.43; *P*=4.7E-20) and chronic fatigue syndrome (rg=0.39, *P*=2.0E-04), among others (Figure 2 and Supplementary Table 14).

### Causal analysis using summary effect estimates (CAUSE)

CAUSE ^36^ showed consistent evidence for a causal effect of the genetic liability of IBS on neuroticism (ΔELPD=-3.6, SE=1.9, *P*=0.031), depression (ΔELPD=-5.9, SE=1.8, *P*=5.4E-03) and anxiety (ΔELPD=-2.9, SE=1.7, *P*=0.049). We also found evidence for reverse causality with a causal effect of the genetic liability of neuroticism and depression on IBS (ΔELPD=-7.3, SE=1.4, *P*=1.5E-07 and ΔELPD=-6.3, SE=1.4, *P*=1.8E-06 respectively) but there was no evidence for a causal relationship when anxiety was considered as exposure and IBS as outcome (Figure 2, Supplementary Table 15a and 15b and Supplementary Figure 6).

## DISCUSSION

In the present study we found extensive genetic sharing between IBS, neuroticism, depression and anxiety, and identified 42 genome-wide significant hits for IBS, of which 38 are novel. Our findings confirm the polygenic architecture of the disorder, with more than 12,000 variants explaining 90% of the *h*^*2*^_SNP,_ and represent a great advance over the previously reported six genome-wide risk loci ^18^. Significant signal enrichment was found in genes showing heightened expression in the brain during early embryonic development and playing prominent roles in mental and digestive disorders, autoimmune diseases and transcription regulation.

Our results confirm a role on IBS of genes involved in brain development and synaptic function as well as genes previously associated with psychiatric conditions ^18^. We detected 27 loci for IBS also associated with at least one of the three mental conditions under study, and found evidence supporting that IBS and neuroticism, which is genetically correlated with many psychiatric disorders ^39^, share a considerable proportion of their genetic background. The widespread common genetic risk sharing with mental conditions was further supported by the positive genetic correlation found between IBS and many psychiatric disorders (i.e. schizophrenia, ADHD, autism or depression) and by the IBS associated variants being located within genes significantly expressed in the brain. These results are in agreement with the higher burden of mental disorders often co-existing in IBS patients, add further evidence of substantial pleiotropy of contributing loci and underscore that genetic influences on IBS may transcend diagnostic boundaries.

Among top findings we identified genes associated with IBS in previous GWAS, such as *CADM2* and *NCAM1*, members of the synaptic cell adhesion molecules that play a role in synapse organization and plasticity ^40,41^. Interestingly, NCAM peptide mimetics have been proven to have both antidepressant and anti-inflammatory effects ^42,43^, pointing them as a potential therapeutic target for IBS. Novel loci for IBS include interesting genes previously associated with depression and other mental disorders, such as *RERE*, that regulates retinoic acid signaling during development ^44–46^, *PCLO*, involved in synaptic vesicle trafficking, *TMEM161B* ^47^, a brain-expressed transmembrane protein ^48^, *RBFOX1*, a splicing regulator mainly expressed in neurons, that is one of the most pleiotropic genes among psychiatric disorders ^49^ or *DRD2*, encoding the dopamine receptor D2R and one of the strongest candidates for psychiatric disorders and traits ^50^ Interestingly, several studies in animal models suggested an important role for dopamine signaling both in the development and progression of inflammatory bowel disease ^51^ and treatment with D2R agonists decreased the severity of ulcerative colitis in mice and rats ^52^. We also provide new insights underlying IBS, showing strong evidence of transcriptional regulation mechanisms playing a role in the disorder, including non-coding RNAs and histone modification. We found genes encoding histones and histone modifying enzymes among top findings, and enrichment of IBS associations in histone acetylation and methylation peaks and in target genes for the histone deacetylase inhibitor belinostat ^53^. These findings are in agreement with previous results involving chromatin modifications in maintenance of anxiety behavior and nociception and in visceral hypersensitivity induced by early-life stress ^54,55^. Additionally, top findings also include non-coding RNAs, an epigenetic mechanism that has been involved in, regulation of genes related with visceral pain response and intestinal permeability ^56–58^. These results add additional evidence towards the role of epigenetic programming in inflammation, visceral pain as well as in intestinal permeability, sensibility and motility in both humans and animal models of IBS ^54,55,59,60^.

Despite many of the findings pointing out neurobiological processes and mental disorders, we also detected links between IBS and gastrointestinal-related phenotypes. Fine mapping showed that 38% of the credible variants were eQTLs for at least one digestive tissue and that credible sets were located in genes enriched in different digestive disorders, including ulcerative colitis and inflammatory bowel disease. In addition, positive genetic correlations were found between IBS and gastric reflux, diverticulitis, hiatus hernia, cholelithiasis/gallstones and gastric/stomach ulcers, among others, which adds evidence on the overlap between the genetic risk for IBS and for other digestive-related disorders and traits. These findings may reflect the multi-factorial etiology proposed for IBS involving psychological factors, abnormal brain functioning and dysregulation of brain-gut interactions ^15,61–63^, as previously proposed in different psychiatric disorders such as depression ^64^.

IBS-associated signals were also enriched in target genes of relevant drugs, including l-lysine or S-adenosylmethionine. L-lysine acts as partial serotonin 5-HT4 receptor antagonist and inhibits serotonin-mediated intestinal pathologies in rats, including anxiety and stress-induced fecal excretion and severity of diarrhea ^65^. Interestingly, l-lysine, and other 5-HT4 receptor antagonists, are promising targets for the treatment of diarrhea-predominant IBS ^66,67^ and may aminorate serotonin disturbances in gut and brain that account for part of intestinal and mental disorders ^65^. Additional drugs of interest include S-adenosylmethionine, involved in neurotransmission signaling that has a putative antidepressant effect^68,69^ or allopurinol that improves inflammatory bowel disease clinical outcomes ^70^, among others.

Despite the high prevalence of psychiatric comorbidities reported in patients with IBS, particularly anxiety and depression, a clear temporal relationship between them has not been well established. We found evidence for a bidirectional causal effect between IBS and neuroticism or depression when accounting for correlated pleiotropy, which strengthens previous evidence ^18^. In addition, we found evidence for a causal effect of the genetic liability of IBS on anxiety. These findings support that IBS increases the risk of subsequent depressive and anxiety disorders described in longitudinal study designs ^71^ and also previous evidence supporting that prior depression raises the risk of developing IBS ^72,73^. We found, however, no evidence for a causal effect of the genetic liability of anxiety on IBS when accounting for correlated pleiotropy, in line with previous results ^18^. Although the sample size for anxiety was more limited and these results may also reflect lack of statistical power. Long term follow-up studies as well as larger datasets and sensitivity analyses are required to confirm the robustness of these results and to better understand the temporal relationship between IBS and comorbid mental conditions.

A major strength of our study is the substantial larger sample size compared with previous studies. By conducting meta-analysis of GWAS summary statistics for IBS and comorbid mental conditions with MTAG we increased the effective sample size from 486,601 in the original IBS dataset to 887,490 individuals and the number of IBS genome-wide significant loci from six in the single-trait analysis to 42. Thirty-eight of them were novel for IBS and 11 were not associated with any of the mental conditions under study, which highlight that MTAG combining GWAS on IBS and mental conditions is a robust strategy to identify trait specific genetic associations. In addition, four of the previously six identified loci were also significant in the present study ^18^. Even though two identified loci demonstrated less association here, their associations were still suggestive (*P*<5E-07) and in concordance in the direction of the effect with the original GWAS study on IBS, which supports validity of the findings across studies.

The study, however, should be considered in the context of some limitations: (i) We did not account for phenotypic overlap and cannot discard that comorbid conditions may have biased the observed results. Also, IBS is considered a highly heterogenous disorder with pathophysiological differences observed among clinical subtypes, between genders, and across age groups and geographic locations ^1^. Accounting for such factors may contribute to better characterize the disorder, capture its genetic background and identify overlap with other comorbid disorders that may impact on IBS risk, prognosis and clinical outcome ^6^; (ii) Despite the strong genetic correlation between IBS and the three mental conditions under study, MiXeR was unable to assess the genetic overlap between IBS, depression and anxiety probably due to the high polygenicity and low SNP heritability estimates for these traits (0.083 and 0.099, respectively) and the limited sample size of the original GWAS on anxiety. We cannot discard, either, that due to lack of power we did not detect IBS signals previously reported for anxiety in the original GWAS or evidence for anxiety increasing the risk for IBS in the causality analyses; (iii) Combining GWAS that differ a great deal in power may lead to inflation of FDR, according to MTAG authors ^24^. In this study we combined GWAS with different sample sizes, however their mean chi-squared was similar and accordingly the max-FDR estimated in our IBS analysis was 0.02, which suggested no inflation of our results. Moreover, despite increasing considerably the effective sample size for IBS through the addition of multiple mental conditions, a number of outcomes were related with gastrointestinal-related phenotypes, which further supports this approach.

In summary, we identified novel risk loci for the IBS, reveal new insights of its polygenic architecture and extended previous knowledge on the genetic overlap and causal relationships between IBS, neuroticism, depression and anxiety. Overall, we advance our understanding of the biological mechanisms underlying IBS, highlighted candidate genes related to brain development and function as well as transcriptional regulation and provide insight into the association between IBS and comorbid mental disorders.

## Data Availability

All data used in the current study is publicly available. Summary statistics for IBS can be download from European Bioinformatics Institute GWAS Catalog (https://www.ebi.ac.uk/gwas/). Summary statistics for neuroticism can be downloaded from https://ctg.cncr.nl/software/summary_statistics/ and http://www.ccace.ed.ac.uk. Summary statistics for depression can be downloaded from https://datashare.ed.ac.uk/handle/10283/3203. Summary statistics for anxiety can be downloaded from http://www.nealelab.is/uk-biobank. Genotype tissue expression (GTEx v8) portal: http://www.gtexportal.org/home/datasets. BRAINEAC: http://www.braineac.org. eQTL catalogue: https://www.ebi.ac.uk/eqtl/Methods/. PsychENCODE: http://resource.psychencode.org. CommonMind Consortium (CMC/CMC): https://www.synapse.org//#!Synapse:syn5585484. WEB-based GEne SeT AnaLysis Toolkit (WebGestAlt): http://www.webgestalt.org.
SNP heritability and genetic correlations: https://github.com/bulik/ldsc. MiXeR: https://github.com/precimed/mixer. Conditional analysis: https://yanglab.westlake.edu.cn/software/gcta/#COJO. Multi-Trait Analysis of GWAS (MTAG): https://github.com/omeed-maghzian/mtag). Fine-mapping: https://github.com/mulinlab/CAUSALdb-finemapping-pip. Functional Mapping and Annotation of Genome-Wide Association Studies (FUMA): https://fuma.ctglab.nl/. Partitioned heritability: https://github.com/bulik/ldsc/wiki/Partitioned-Heritability. MR-Base database: https://github.com/MRCIEU/mrbase_casestudies. Causal Analysis Using Summary Effect estimates (CAUSE): https://jean997.github.io/cause/pipeline.html. The use of each software tools has been described in the Methods section. Analysis code and scripts used in the current study are available upon request from the corresponding

https://ctg.cncr.nl/software/summary_statistics/

http://www.ccace.ed.ac.uk

https://datashare.ed.ac.uk/handle/10283/3203

http://www.nealelab.is/uk-biobank

http://www.gtexportal.org/home/datasets

http://www.braineac.org

https://www.ebi.ac.uk/eqtl/Methods/

http://resource.psychencode.org

https://www.synapse.org//#!Synapse:syn5585484

## Abbreviations

ADHD: attention-deficit/hyperactivity disorder
AIC: Akaike Information Criterion
ASD: autism spectrum disorders
ATC: Anatomical Therapeutic Chemical
CAUSE: Causal Analysis Using Summary Effect estimates
IBS: irritable bowel syndrome
FDR: false discovery rate
ELPD: expected log pointwise posterior density
FUMA: Functional Mapping and Annotation of GWAS
GCTA: Genome-wide Complex Trait Analysis
GWAS: Genome-wide association study
*h*^*2*^_SNP_: SNP heritability
LDSC: Linkage disequilibrium score regression
MAGMA: Generalized gene-set analysis of GWAS data
MTAG: Multi-Trait Analysis of GWAS

## Declarations

### Ethics approval and consent to participate

This article contains results derived from data from human participants collected by several studies performed by previous studies. All participants gave informed consent in all the corresponding original studies. Our study is based on the large-scale GWAS datasets, and not the individual-level data. Hence, ethical approval was not applicable.

### Consent for publication

Not applicable.

### Availability of data and material

All data used in the current study is publicly available. Summary statistics for IBS can be download from European Bioinformatics Institute GWAS Catalog (https://www.ebi.ac.uk/gwas/). Summary statistics for neuroticism can be downloaded from https://ctg.cncr.nl/software/summary_statistics/ and http://www.ccace.ed.ac.uk. Summary statistics for depression can be downloaded from https://datashare.ed.ac.uk/handle/10283/3203. Summary statistics for anxiety can be downloaded from http://www.nealelab.is/uk-biobank. Genotype tissue expression (GTEx v8) portal: http://www.gtexportal.org/home/datasets. BRAINEAC: http://www.braineac.org. eQTL catalogue: https://www.ebi.ac.uk/eqtl/Methods/. PsychENCODE: http://resource.psychencode.org. CommonMind Consortium (CMC/CMC): https://www.synapse.org//#!Synapse:syn5585484. WEB-based GEne SeT AnaLysis Toolkit (WebGestAlt): http://www.webgestalt.org. SNP heritability and genetic correlations: https://github.com/bulik/ldsc. MiXeR: https://github.com/precimed/mixer. Conditional analysis: https://yanglab.westlake.edu.cn/software/gcta/#COJO. Multi-Trait Analysis of GWAS (MTAG): https://github.com/omeed-maghzian/mtag). Fine-mapping: https://github.com/mulinlab/CAUSALdb-finemapping-pip. Functional Mapping and Annotation of Genome-Wide Association Studies (FUMA): https://fuma.ctglab.nl/. Partitioned heritability: https://github.com/bulik/ldsc/wiki/Partitioned-Heritability. MR-Base database: https://github.com/MRCIEU/mrbase_casestudies. Causal Analysis Using Summary Effect estimates (CAUSE): https://jean997.github.io/cause/pipeline.html. The use of each software tools has been described in the Methods section. Analysis code and scripts used in the current study are available upon request from the corresponding authors.

## Authors’ contributions

SA, MSA, JC, LVR, NLL and MR designed and supervised the study. SA analyzed the data. SA, MSA, JC and MR wrote the manuscript draft. All authors contributed to the interpretation of results and critically reviewed the manuscript.

