## Supplementary materials for "Genome-wide multi-trait analysis of irritable bowel syndrome and related mental conditions identifies 38 new genetic hits"

Online methods

**Online methods**

S1. Summary statistics

We used publicly available SNP-level GWAS summary statistics for IBS [1], neuroticism [2], depression [3] and anxiety (Table 1). Summary statistics for IBS were obtained from the most recent and largest GWAS conducted to date in 53,400 cases and 433,201 controls [1]. For neuroticism, depression and anxiety we searched for the summary statistics of the largest datasets publicly available to date (December 2021). Quality control for genetic variants for all datasets included removal of duplicated and/or ambiguous SNPs and insertions or deletions, MAF<0.01, INFO score <0.90 (INFO score was not available for depression and anxiety summary statistics), SE<0, *P*-value out of bounds and N=0.

S2. SNP-based heritability genetic correlation and overlap

SNP heritability (h^2^_SNP_) and pair-wise genetic correlation between IBS and each mental condition (i.e. neuroticism depression or anxiety) was calculated using linkage disequilibrium score regression (LDSC) analysis [4]. Conversion of h^2^_SNP_ estimates from observed to liability scale was done using a population prevalence of 11% , 25%, 30% and 14% for IBS, neuroticism, depression and anxiety, respectively. Polygenic overlap, irrespective of genetic correlation, between IBS and each mental condition was quantified using MiXeR [5]. MiXeR provides univariate estimates of the number of trait-influencing loci for each trait as well as bivariate estimates of genome-wide genetic overlap between pairs of traits. We calculated the Dice coefficient which estimates the proportion of SNPs shared by two traits. MiXeR also calculates the proportion of trait-influencing variants with concordant direction of effects for both traits. Model fit was assessed using the Akaike Information Criterion (AIC). In the univariate model, AIC negative values indicate that there is not enough power in the input summary statistics and MiXeR is not recommended in this situation [5]. In the bivariate models, the best model (i.e. the model estimating the number of shared variants between the traits) is compared with two models representing the maximal and minimal possible polygenic overlap. AIC positive values indicate that the best model explain the GWAS signal better than the maximal or minimal model while negative values indicate poor model fit.

S3. Multi-Trait Analysis of GWAS (MTAG)

To identify new loci for IBS, SNP-level GWAS for IBS, neuroticism, depression and anxiety were meta-analyzed using MTAG [6], which integrates summary statistics across correlated traits and generates new trait-specific effect estimates and P-values. To discard inflation in our results we calculated the max-false discovery rate (max-FDR). A primary assumption of MTAG is that the variance-covariance matrix of effects is identical across SNPs. Violation of this assumption can lead to an inflated false discovery rate (FDR). MTAG simulates FDR under a worst-case scenario to provide an FDR upper bound (max FDR) of how severely deviations will affect FDR. Acceptable FDR should be below 5% [6,7].

Independent lead SNPs from MTAG-IBS results (*P*-value<5-E08) were identified through clumping (r2 = 0.05, kb = 5000) using the 1000 Genomes Project Phase 3 European reference panel (<http://www.internationalgenome.org/>) and PLINK1.09 as described by Eijsbouts et al. [1]. We carried out conditional analyses to evaluate independence between secondary (within 5000 kb and r2 < 0.2) and index variants within each locus. For loci with more than two secondary lead variants, we further confirmed whether secondary lead variants were independent among each other conditioning on the secondary lead variant with the lowest multi-trait analysis P-value. Conditional analyses were performed using COJO implemented in Genome-wide Complex Trait Analysis (GCTA) [8]. Overlap between MTAG-IBS and previous genome-wide significant independent lead SNPs for each trait was assessed according to distance and linkage disequilibrium (previous lead SNPs within +/-5000kb from any of the MTAG-IBS lead SNPs and r2>0.2). Independent signals identified were further confirmed using conditional analysis using COJO as described above.

S4. Credible variants and functional annotation

Sets of credible variants (credible-sets) were identified by fine-mapping the independent lead SNPs of MTAG-IBS using three different tools , FINEMAP 1.3.1 [9], PAINTOR v3.0 [10] and CAVIARBF v.0.2.1 [11] following the pipeline available elsewhere [12]. Variants located in a region of 5000 kb around the lead SNPs were included in the analysis and we assumed that there was only one causal variant per locus. We used the recommended parameters of each tool and only variants identified by all three methods were considered.

Functional annotation was conducted using ANNOVAR [13] on the credible variants as implemented in FUMA[14]. The categories used to predict the SNP functional consequences included CADD scores [15], RegulomeDB scores [16] and chromatin states [17,18]. CADD scores predict how deleterious the SNP effect is on protein structure/function based on 63 functional annotations. A threshold of CADD >= 12.37 was considered for detecting deleterious variants [15]. The RegulomeDB score is based on information from eQTLs and chromatin marks and predicts the likelihood of regulatory functionality. Lower RegulomeDB scores indicate increasing evidence of having regulatory function [19]. The chromatin state represents the accessibility of genomic regions considering 15 categorical states with lower state indicating higher accessibility and states from 1 to 7 referring to open chromatin states. Traits showing suggestive evidence of association (*P*<5E-07) with the SNPs in credible sets were identified using the NHGRI-EBI GWAS catalog [20]. We queried SNPs for known eQTLs using the genotype tissue expression (GTEx v8) portal [21], BRAINEAC [22], eQTL catalogue [23], PsychENCODE [24] and CommonMind Consortium (CMC/CMC) [25]. The specific databases used can be found in Supplementary Note 3.

SNPs in credible sets were annotated to genes based on physical proximity (using the default parameters), eQTL (based on GTEx v8, BRAINEAC, eQTL catalogue, PsychENCODE and CMC/CMC using the databases previously described) and chromatin interaction using FUMA (databases detailed in Supplementary Notes 3 and 4). These genes were used in gene-set enrichment analyses in the GENE2FUNC module of FUMA. Enrichment was tested among the predefined sets of differentially expressed genes in GTEx v8 (54 tissue types) and Brainspan (29 different ages of samples and 11 general developmental stages) using hypergeometric test with protein coding genes as background genes. Genes were also tested for enrichment in gene-sets from the Molecular Signatures Database (MSigDB version v6.2) including Biocarta, gene ontology (GO), KEGG, Reactome and GWAS Catalog. We corrected for multiple comparisons using FDR.

S5. Datasets used in FUMA:

S5.1. SNPs in eQTLs

eQTLcatalogue/BrainSeq_ge_brain.txt.gz

PsychENCODE/PsychENCODE_eQTLs.txt.gz

CMC/CMC_SVA_cis.txt.gz

CMC/CMC_SVA_trans.txt.gz

CMC/CMC_NoSVA_cis.txt.gz

CMC/CMC_NoSVA_trans.txt.gz

BRAINEAC/CRBL.txt.gz

BRAINEAC/FCTX.txt.gz

BRAINEAC/HIPP.txt.gz

BRAINEAC/MEDU.txt.gz

BRAINEAC/OCTX.txt.gz

BRAINEAC/PUTM.txt.gz

BRAINEAC/SNIG.txt.gz

BRAINEAC/TCTX.txt.gz

BRAINEAC/THAL.txt.gz

BRAINEAC/WHMT.txt.gz

BRAINEAC/aveALL.txt.gz

GTEx/v8/Brain_Amygdala.txt.gz

GTEx/v8/Brain_Anterior_cingulate_cortex_BA24.txt.gz GTEx/v8/Brain_Caudate_basal_ganglia.txt.gz GTEx/v8/Brain_Cerebellar_Hemisphere.txt.gz

GTEx/v8/Brain_Cerebellum.txt.gz

GTEx/v8/Brain_Cortex.txt.gz

GTEx/v8/Brain_Frontal_Cortex_BA9.txt.gz

GTEx/v8/Brain_Hippocampus.txt.gz

GTEx/v8/Brain_Hypothalamus.txt.gz GTEx/v8/Brain_Nucleus_accumbens_basal_ganglia.txt.gz GTEx/v8/Brain_Putamen_basal_ganglia.txt.gz

GTEx/v8/Brain_Spinal_cord_cervical_c-1.txt.gz

GTEx/v8/Brain_Substantia_nigra.txt.gz

GTEx/v8/Colon_Sigmoid.txt.gz GTEx/v8/Colon_Transverse.txt.gz GTEx/v8/Esophagus_Gastroesophageal_Junction.txt.gz GTEx/v8/Esophagus_Mucosa.txt.gz GTEx/v8/Esophagus_Muscularis.txt.gz GTEx/v8/Small_Intestine_Terminal_Ileum.txt.gz

GTEx/v8/Stomach.txt.gz

S5.2. Chromatin interaction datasets used for gene mapping

EP/PsychENCODE/EP_links_oneway.txt.gz:

HiC/PsychENCODE/Promoter_anchored_loops.txt.gz:

HiC/Giusti-Rodriguez_et_al_2019/Adult_Cortex.txt.gz:

HiC/Giusti-Rodriguez_et_al_2019/Fetal_Cortex.txt.gz:

HiC/GSE87112/Dorsolateral_Prefrontal_Cortex.txt.gz

HiC/GSE87112/Hippocampus.txt.gz

HiC/GSE87112/Small_Bowel.txt.gz

Roadmap – brain: E067 E068 E069 E070 E071 E072 E073 E074 E081 E082 E075 E076 E106 E077 E078 E079 E084 E085 E109 E101 E102 E103 E092 E094 E110 E111

S5.3. Tissue specific gene expression datasets used for MAGMA gene-property analysis

GTEx/v8/gtex_v8_ts_avg_log2TPM

GTEx/v8/gtex_v8_ts_general_avg_log2TPM

BrainSpan/bs_age_avg_log2RPKM

BrainSpan/bs_dev_avg_log2RPKM

S6. Causal analysis using summary effect estimates (CAUSE)

Causal relationships between IBS and correlated traits were assessed considering independent variants (r2 = 0.05; kb = 5000) associated with the exposure with *P*<1.0E-03 using CAUSE [30]. Bidirectional relationships were tested considering IBS as exposure and depression, anxiety or neuroticism as outcomes and vice-versa. Given that SE was not avaiable from the largest study on neuroticism to date [31], we used the GWAS dataset on neuroticism by Luciano et al. in 329,821 subjects as an alternative [32]. The strengths of CAUSE involve accounting for correlated horizontal pleiotropic effects (i.e. when a variant affects the outcome and the mediator through shared heritable factors) and using a less stringent significance threshold (*P*<1.0E-3) allowing the incorporation of more variants to the analyses. CAUSE compares two nested models, a sharing and a causal model. Both models allow for horizontal pleiotropy (correlated pleiotropy (eta)) but only the casual model includes a causal effect parameter (gamma). The sharing and the causal model are compared against a null model and against each other. Model comparisons are carried out using the expected log pointwise posterior density (ELPD), a Bayesian model comparison approach that estimates how well the posterior distributions of a particular model are expected to predict a new set data. When *P* <0.05 the second model fits the data better than the first model. There is evidence of causal effects when the causal model represents a significant improvement in the model fit of the sharing model.
