## Supplementary figures for "Genome-wide multi-trait analysis of irritable bowel syndrome and related mental conditions identifies 38 new genetic hits"

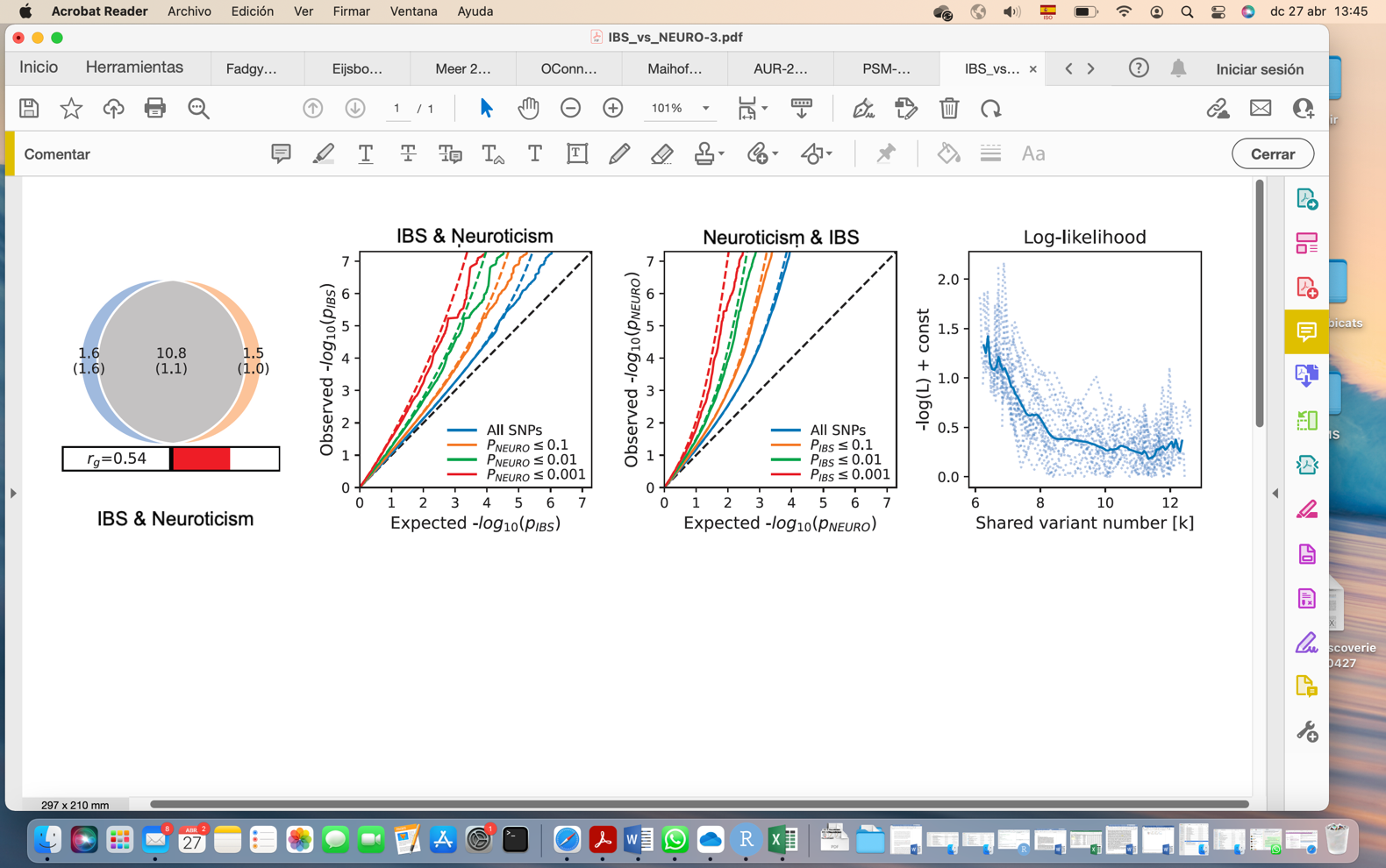

(A) (B)	(C)	(D)

**Supplementary Figure 1. MiXeR results for IBS and neuroticism.**

A) Venn diagram depicting the estimated number of trait-influencing variants shared (gray) between IBS and neuroticism. Unique variants for each trait are depicted in blue for IBS and orange for neuroticism. The number of trait-influencing variants in thousands is shown, with the standard error in thousands provided in parentheses. The size of the circles reflects the polygenicity of each phenotype, with larger circles corresponding to greater polygenicity. The estimated genetic correlation (r_g_) is shown in the bar. Red color indicates positive genetic correlation. B) and C) depict conditional Q–Q plots of observed versus expected −log10 p-values in the primary trait as a function of significance of association with a secondary trait at the level of p ≤ 0.1 (orange lines), p ≤ 0.01 (green lines), p ≤ 0.001 (red lines). Blue line indicates all SNPs. Dotted lines in blue, orange, green, and red indicate model predictions for each stratum. Black dotted line is the expected Q–Q plot under null (no SNPs associated with the phenotype). D) Log-likelihood curves highlighting the goodness of model fit. The minimum point indicates the best-fitting model estimate of the number of influencing variants shared between two traits (Supplementary Table 1).

**Chr1: rs301806**

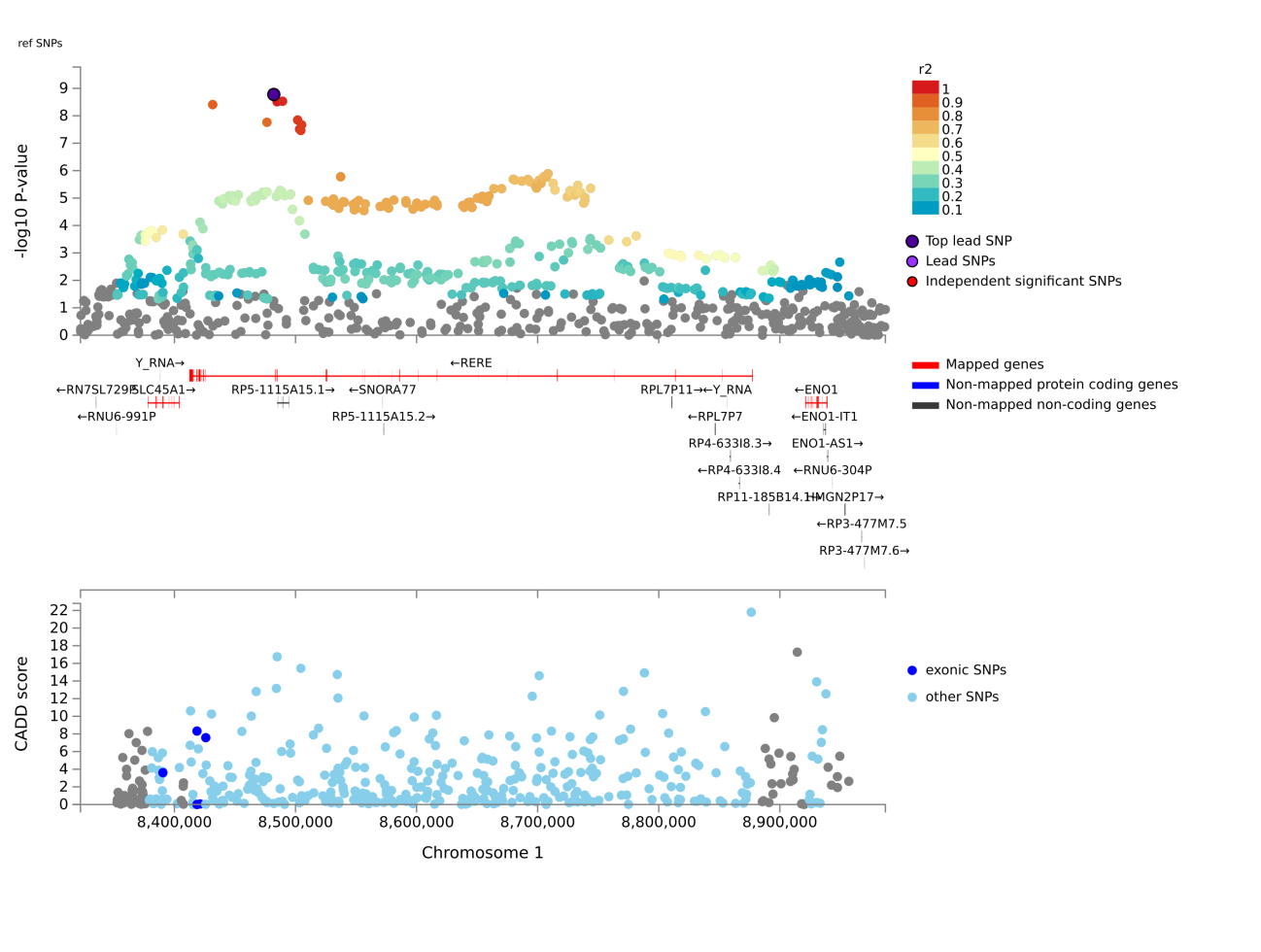

**Chr1: rs11206127**

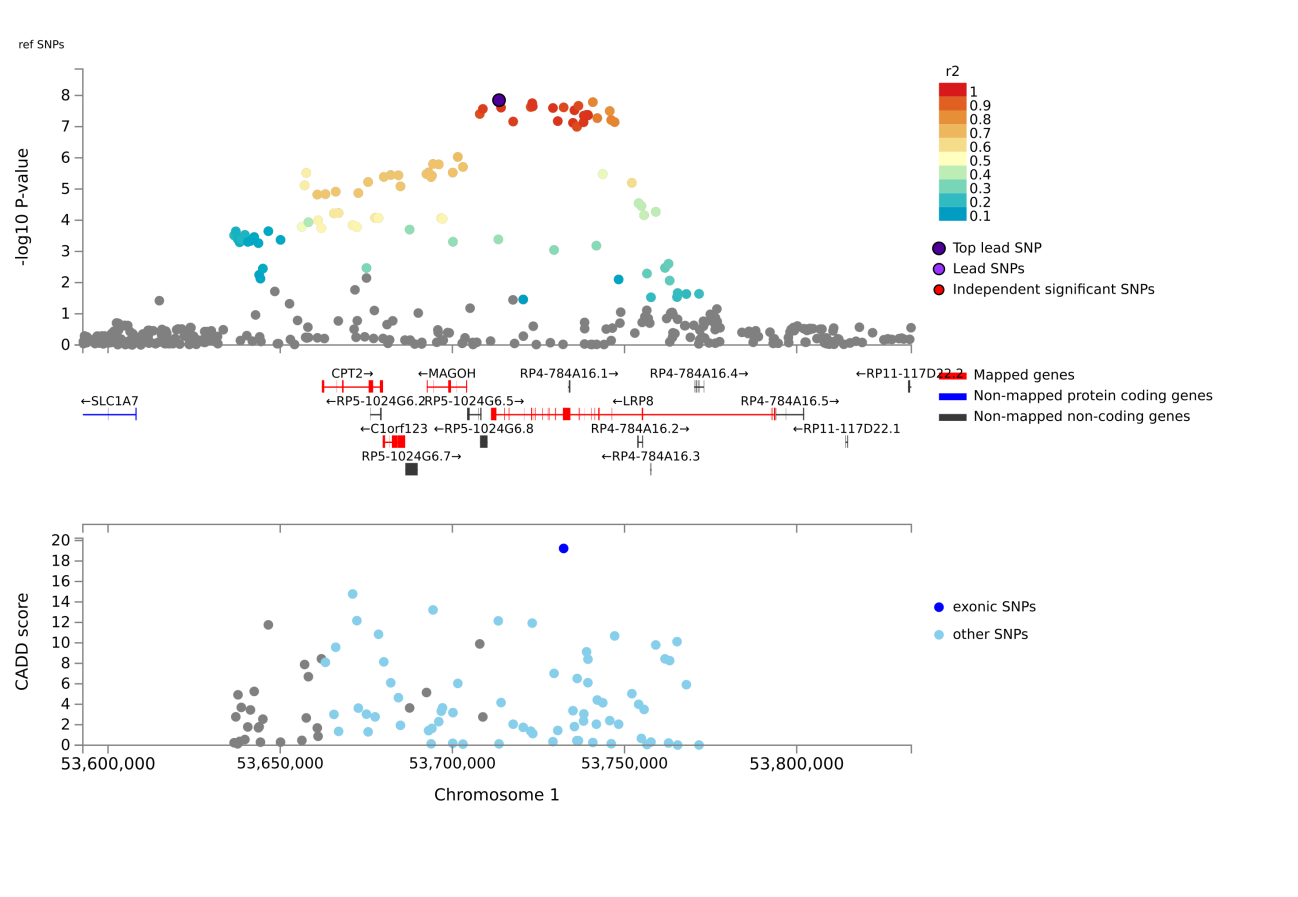

**Chr1: rs12755507**

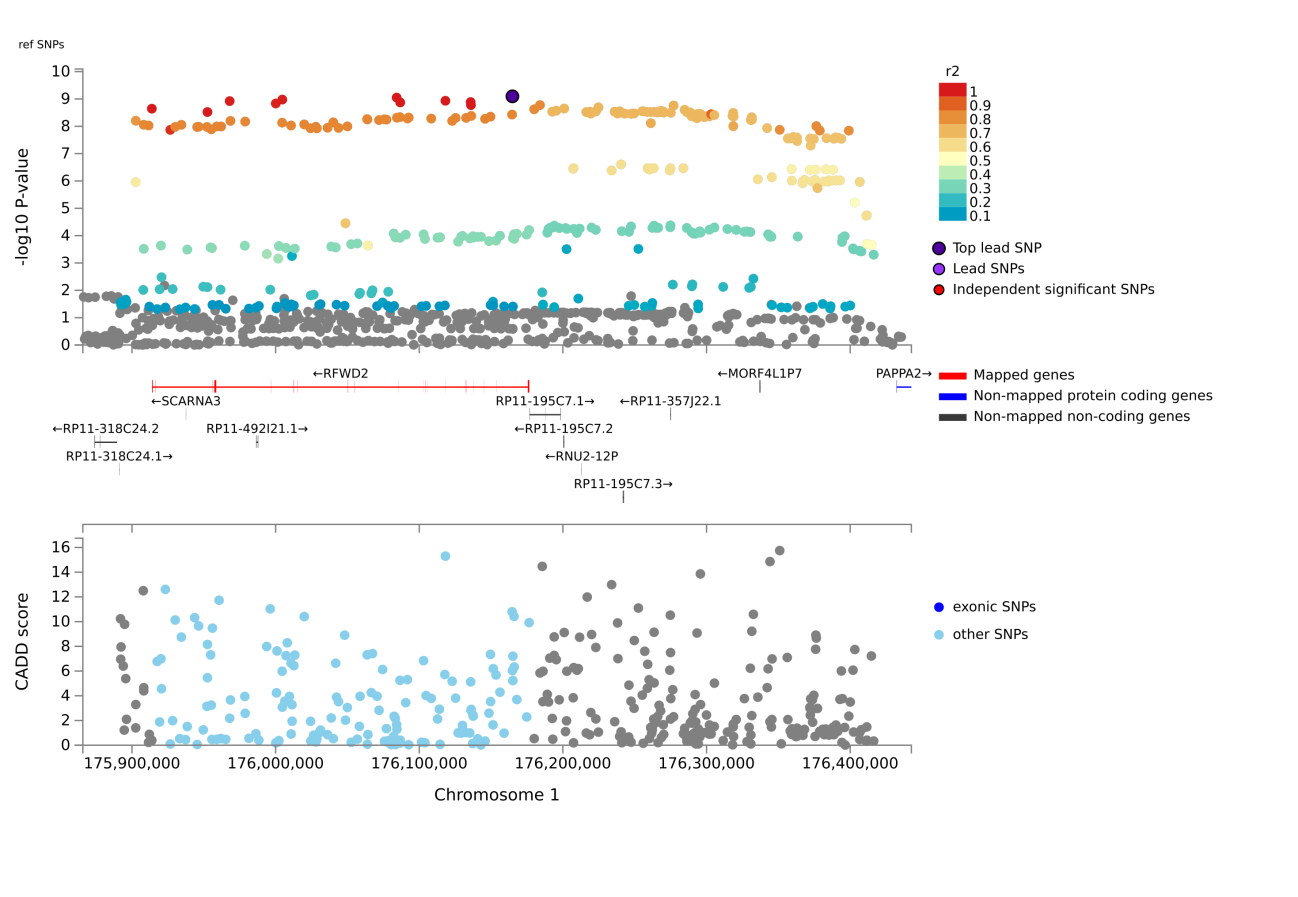

**Chr1: rs113198479**

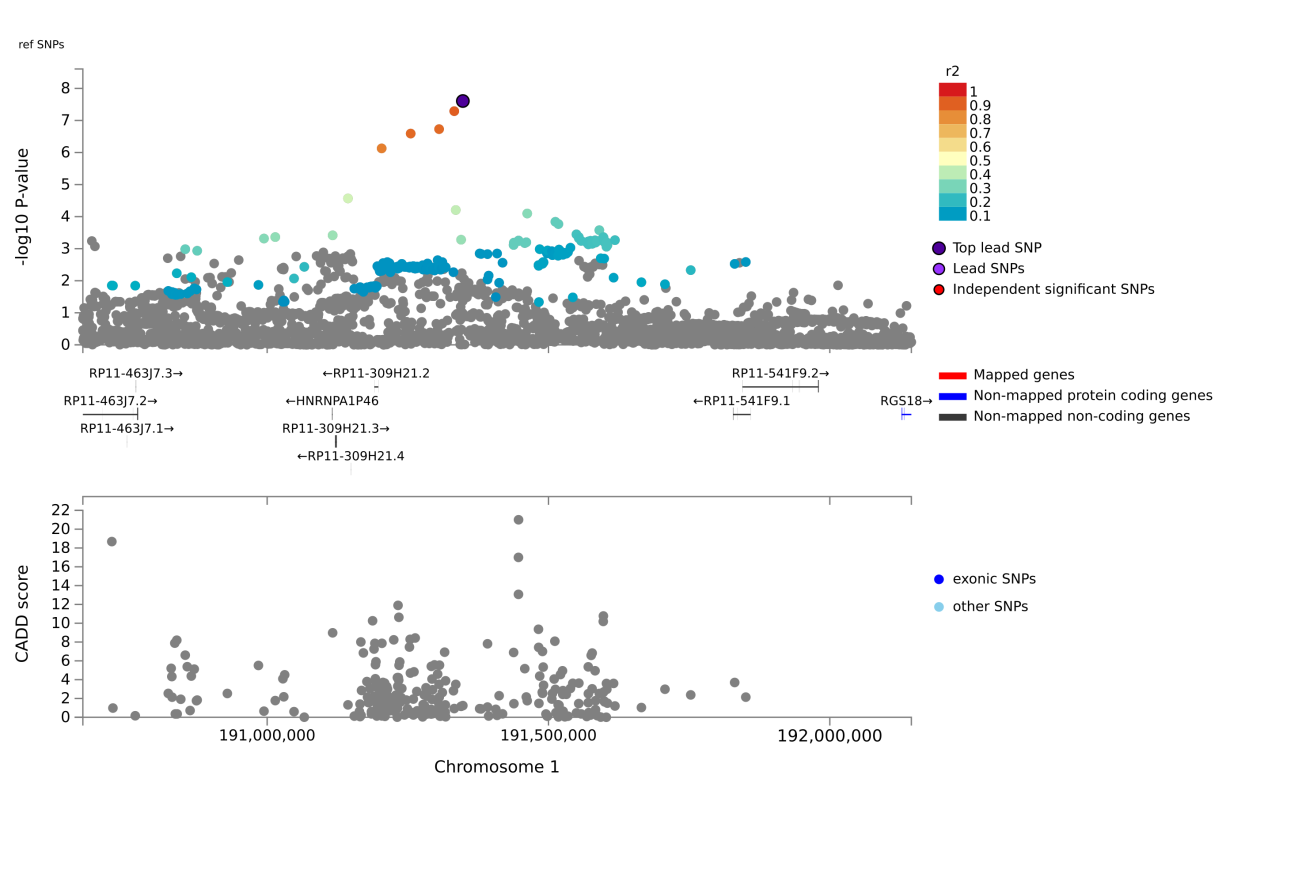

**Chr1:** **rs72740550**

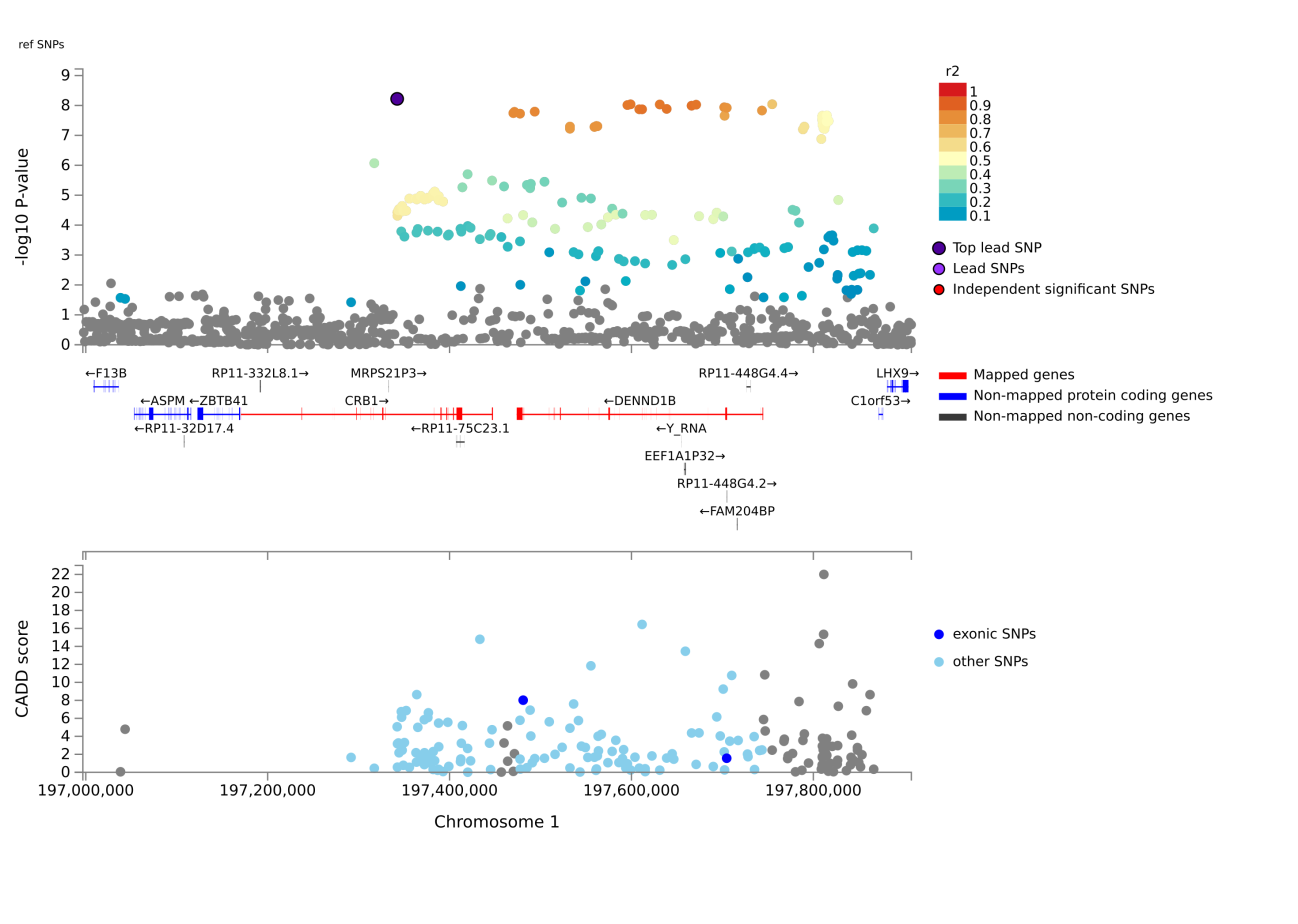

**Chr2: rs115962846**

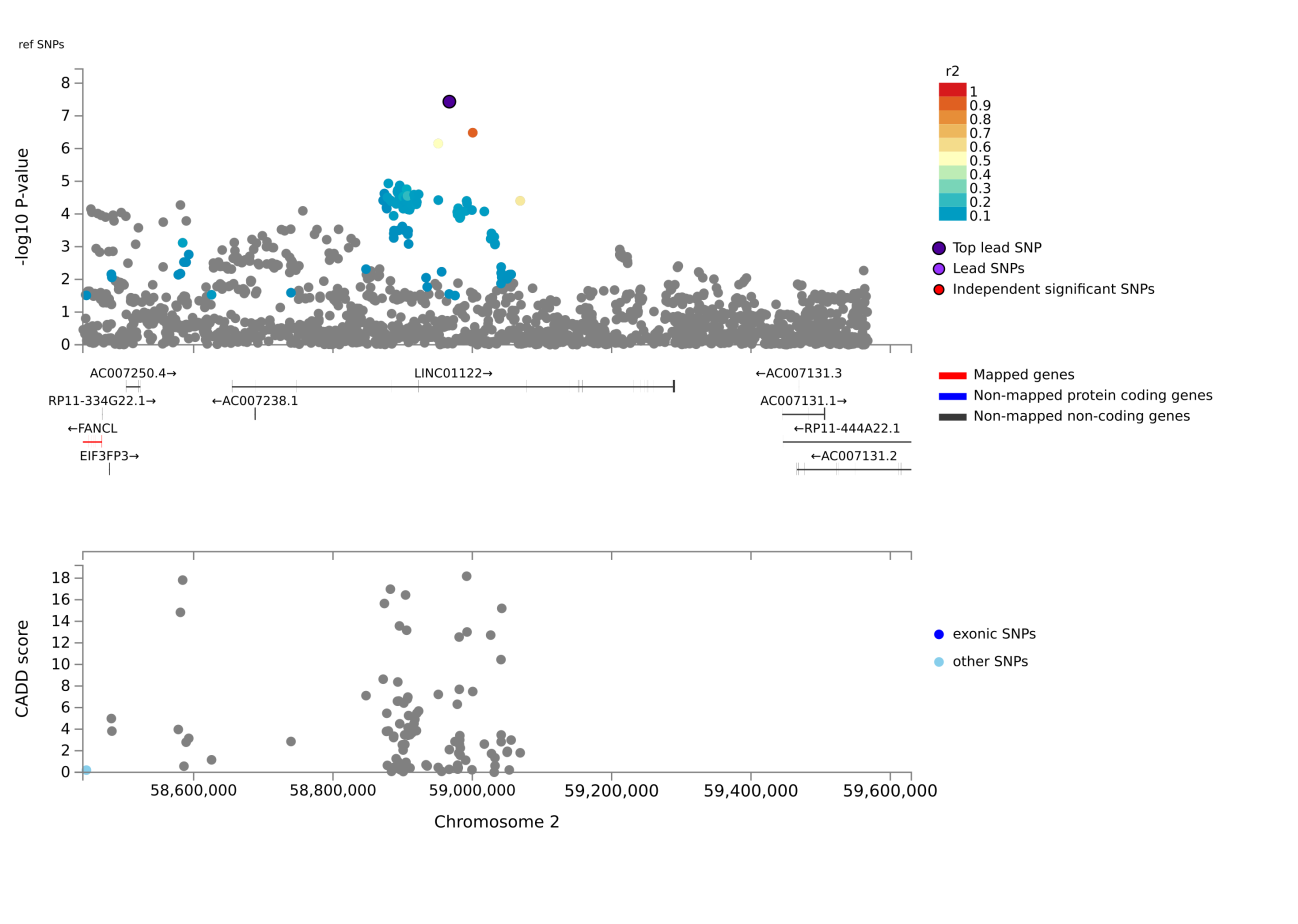

**Chr2: rs28496790**

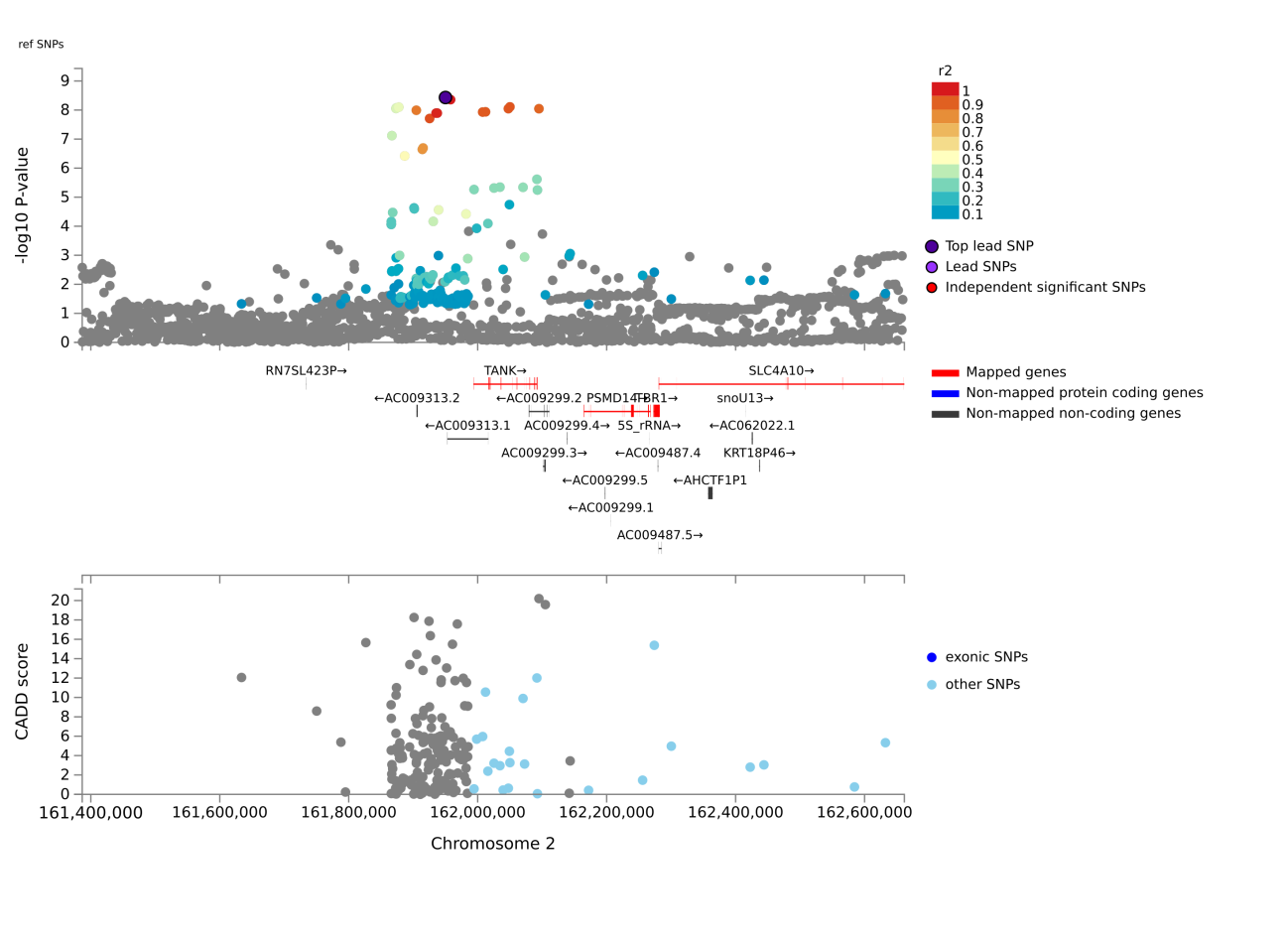

**Chr2: rs138218528**

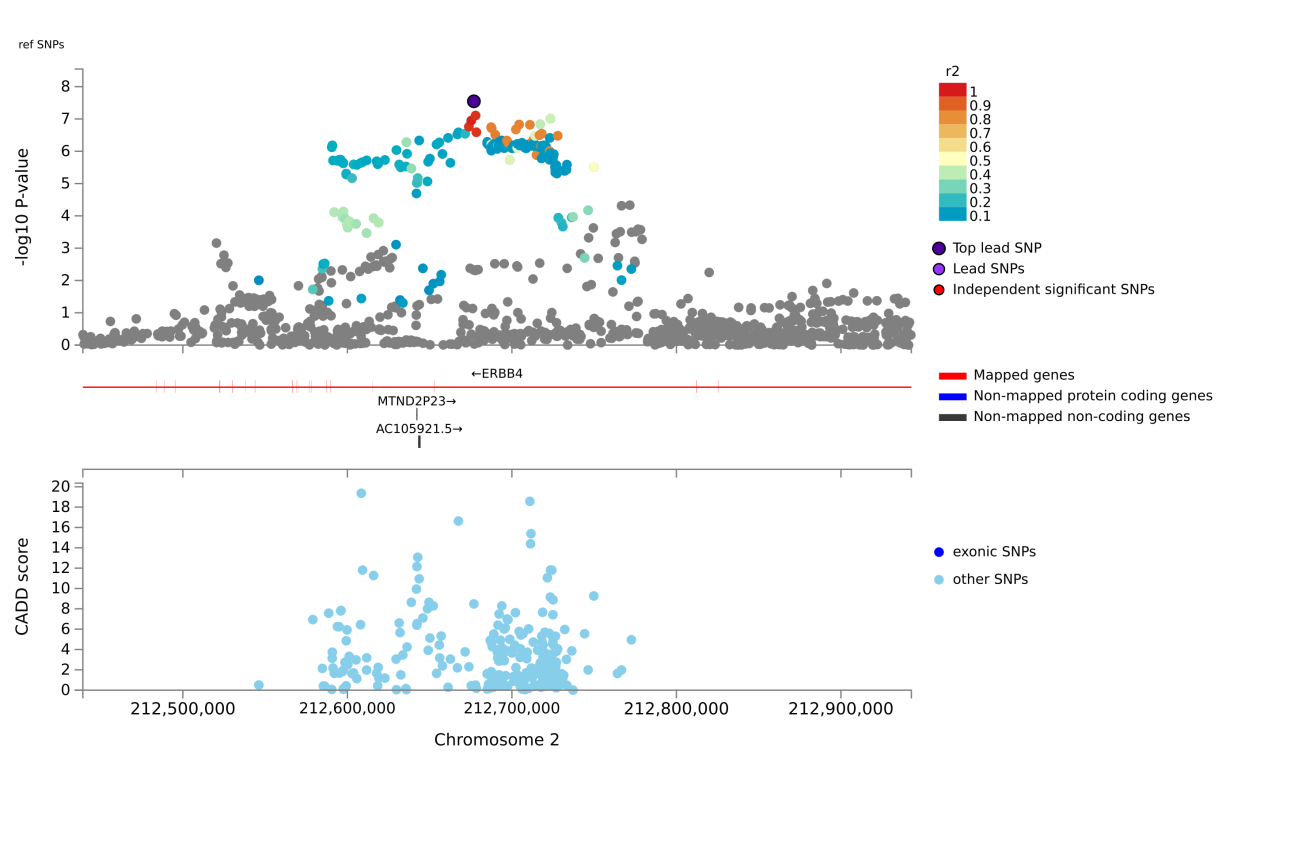

**Chr3: rs62246276**

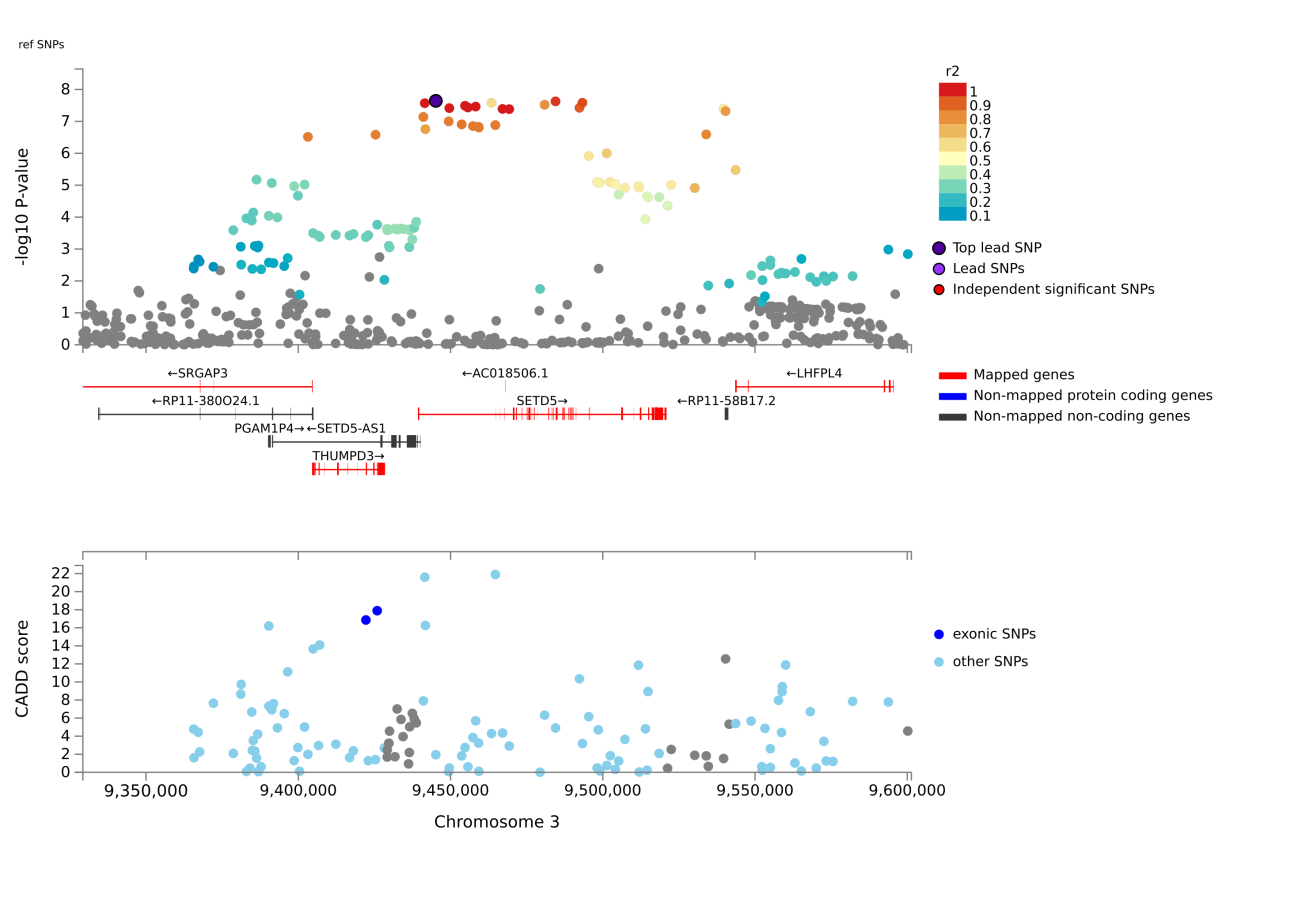

**Chr3: rs67416405**

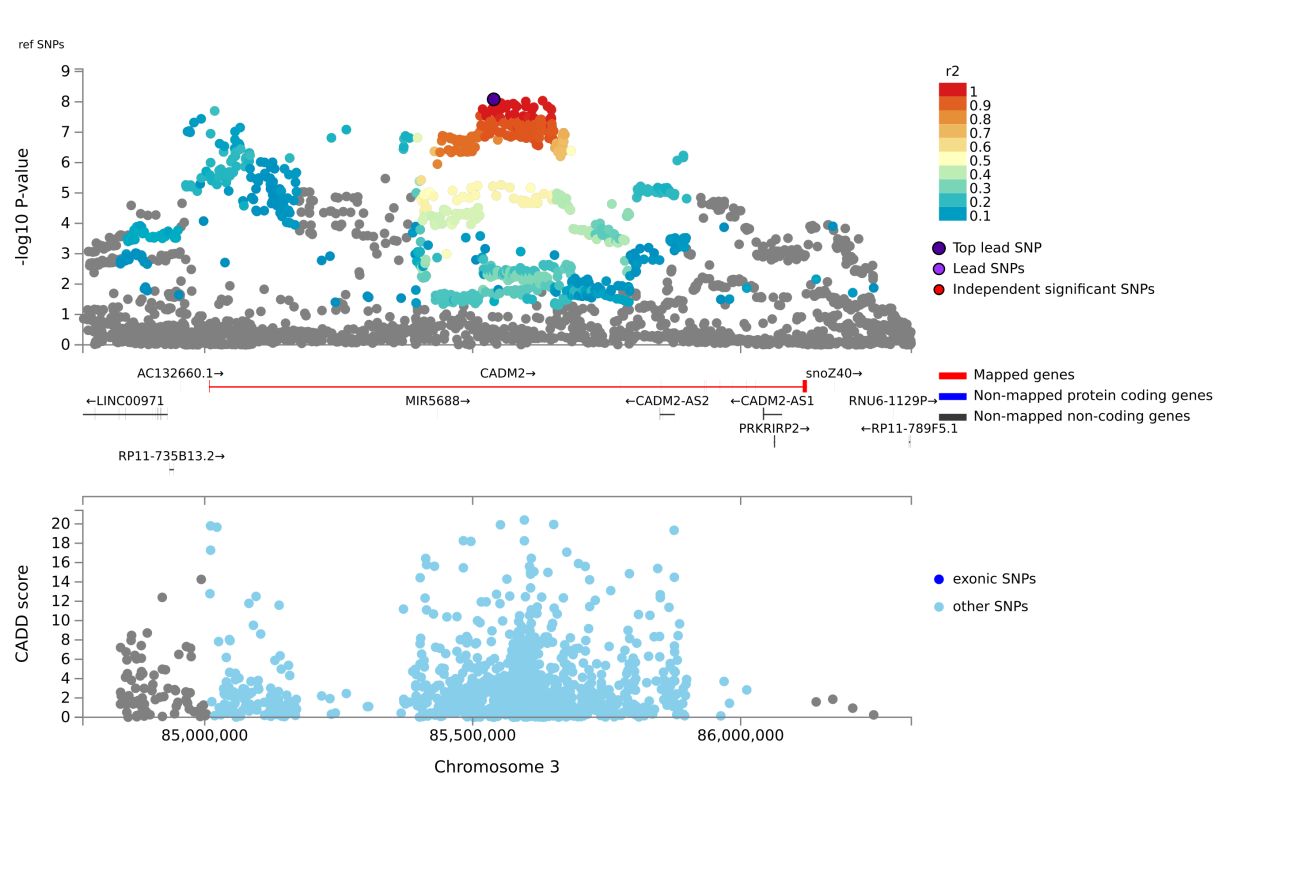

**Chr3: rs1729951**

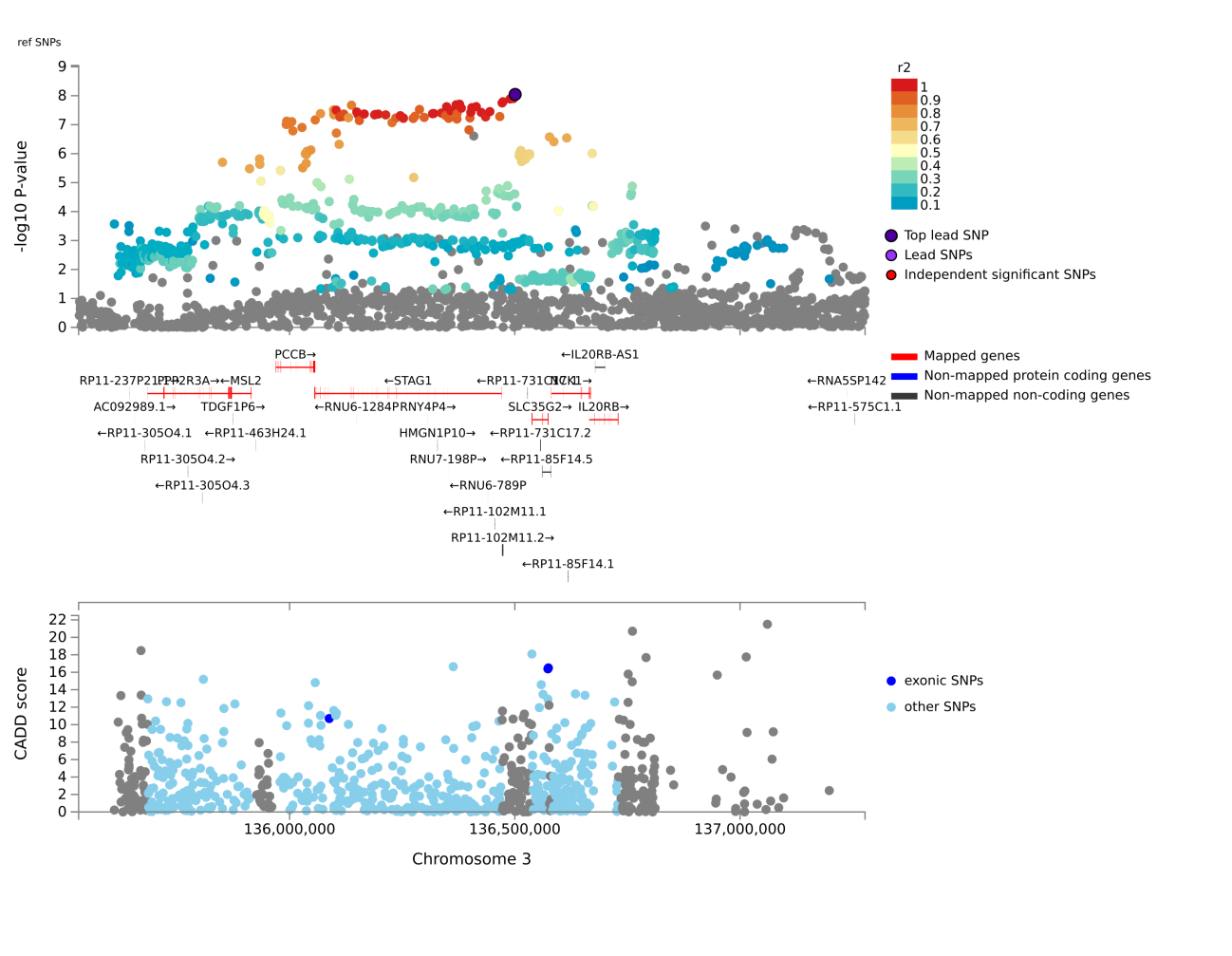
**Chr4: rs1442129**

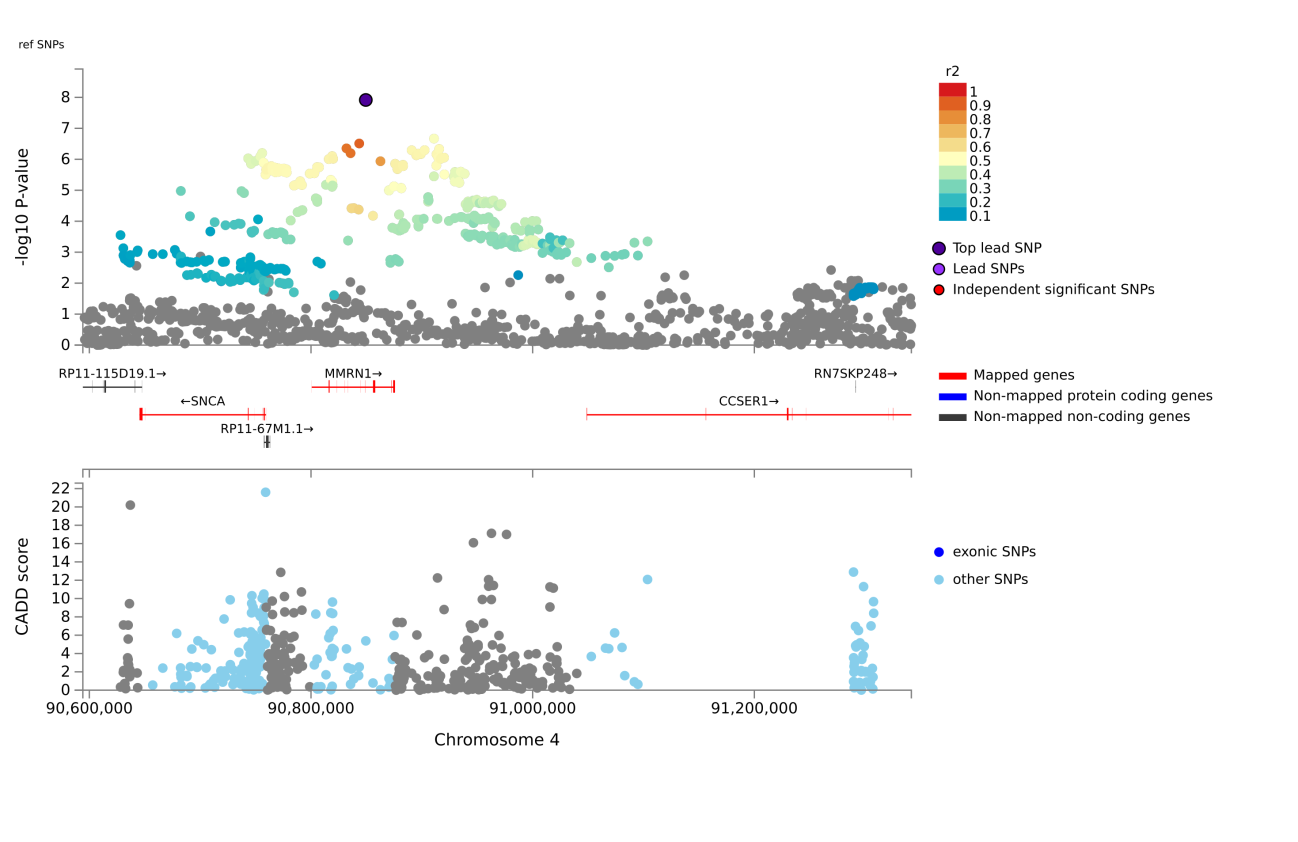

**Chr4: rs77087420**

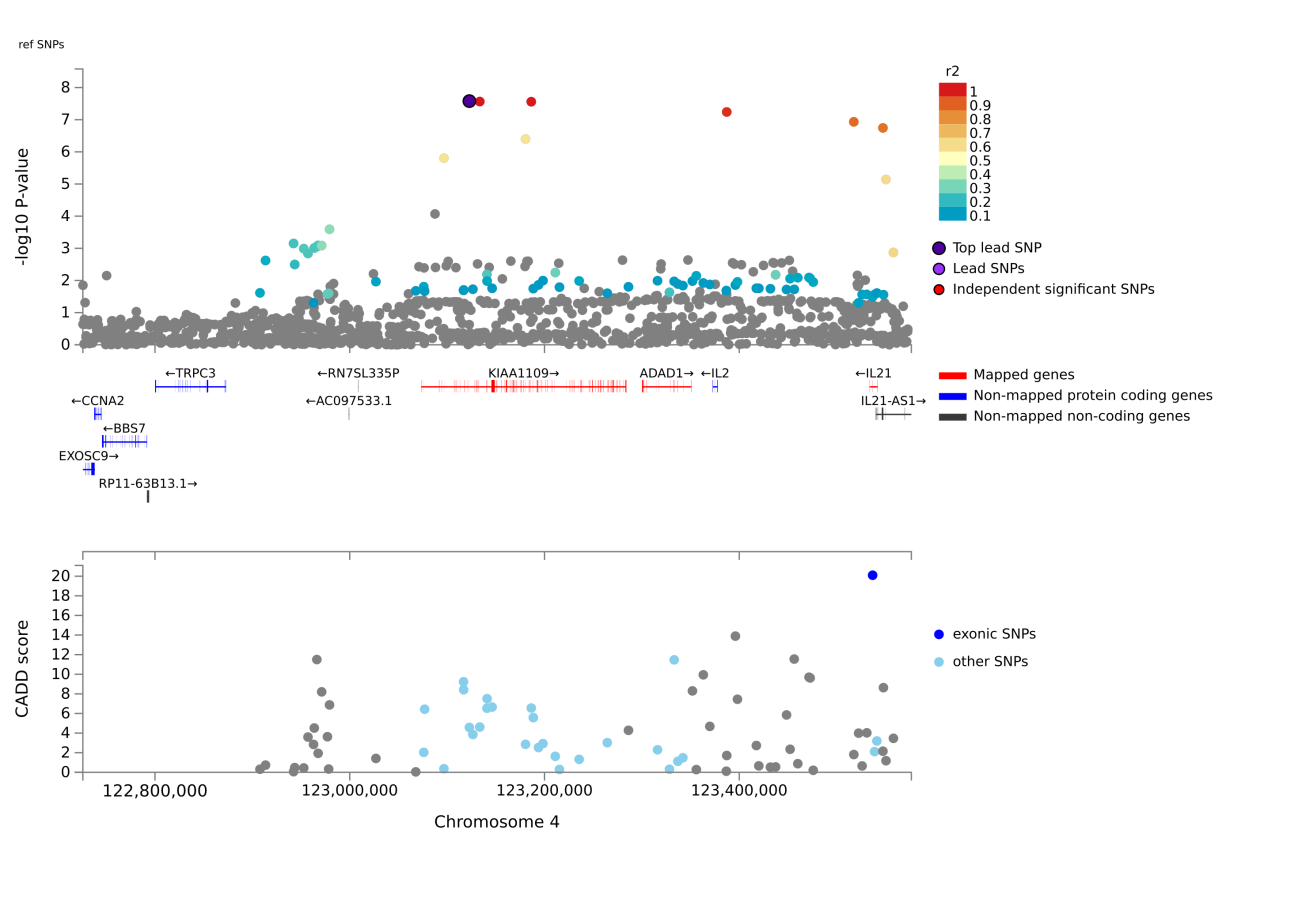

**Chr5: rs12513440**

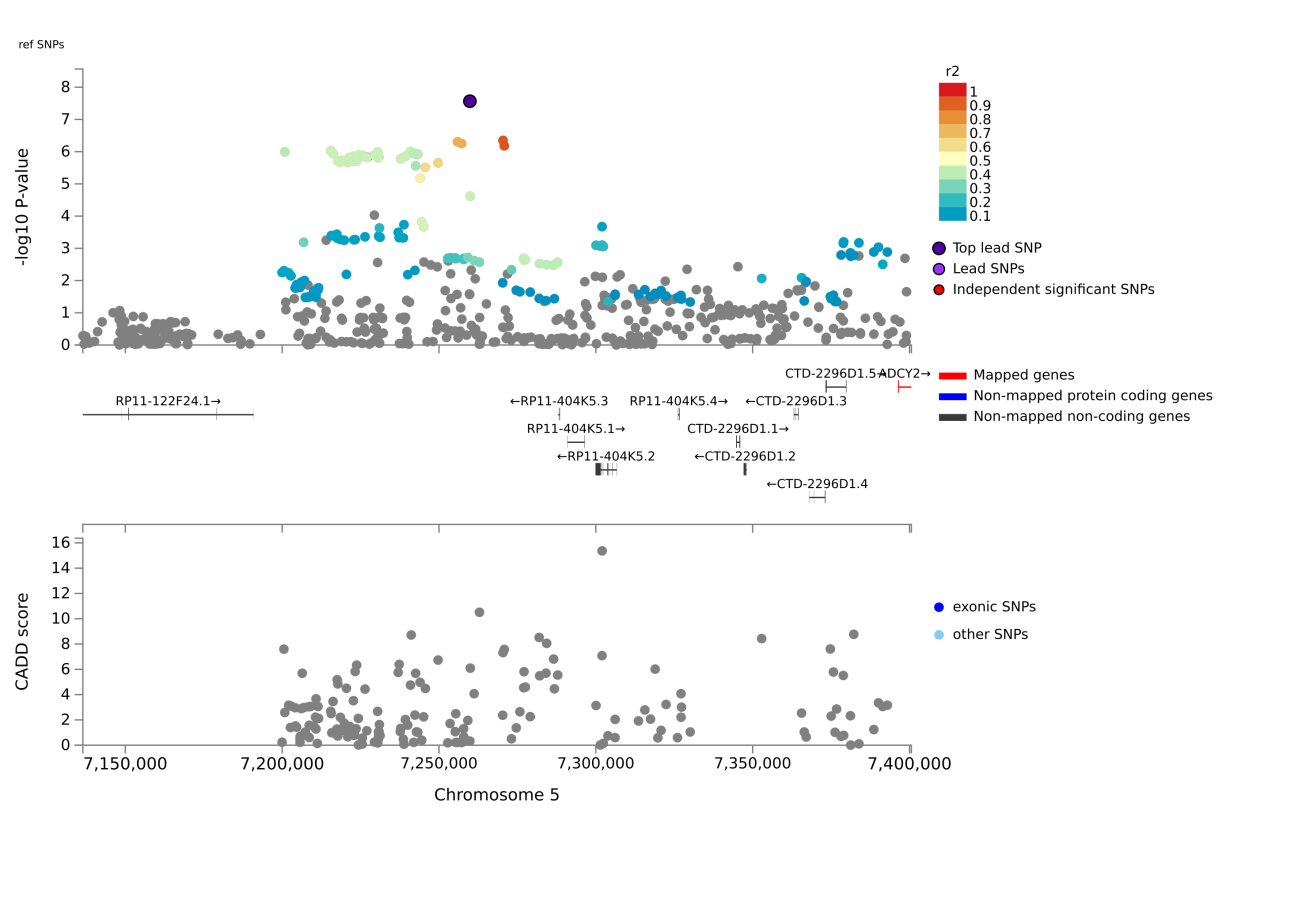

**Chr5: rs3099439**

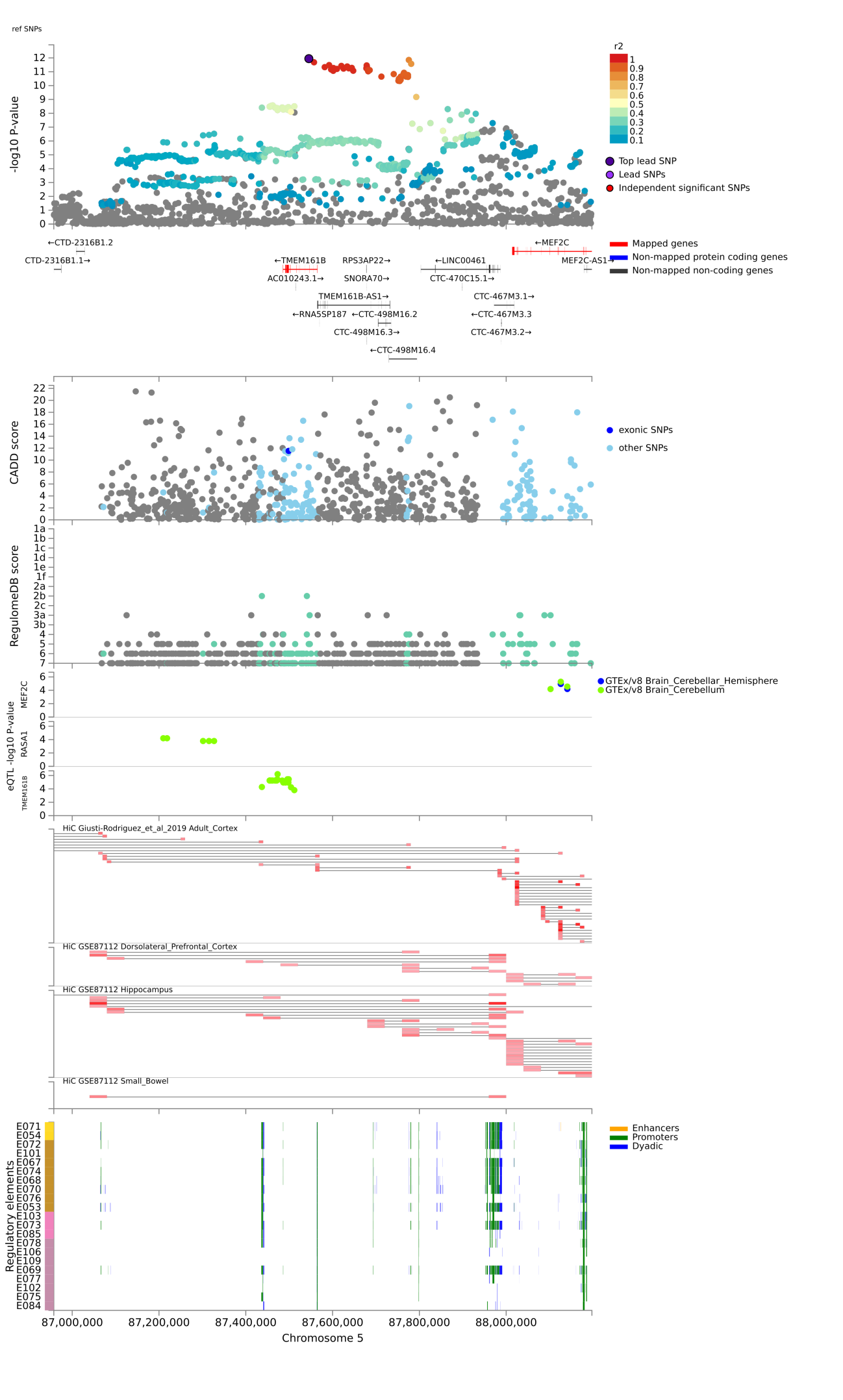

**Chr5: rs4481363**

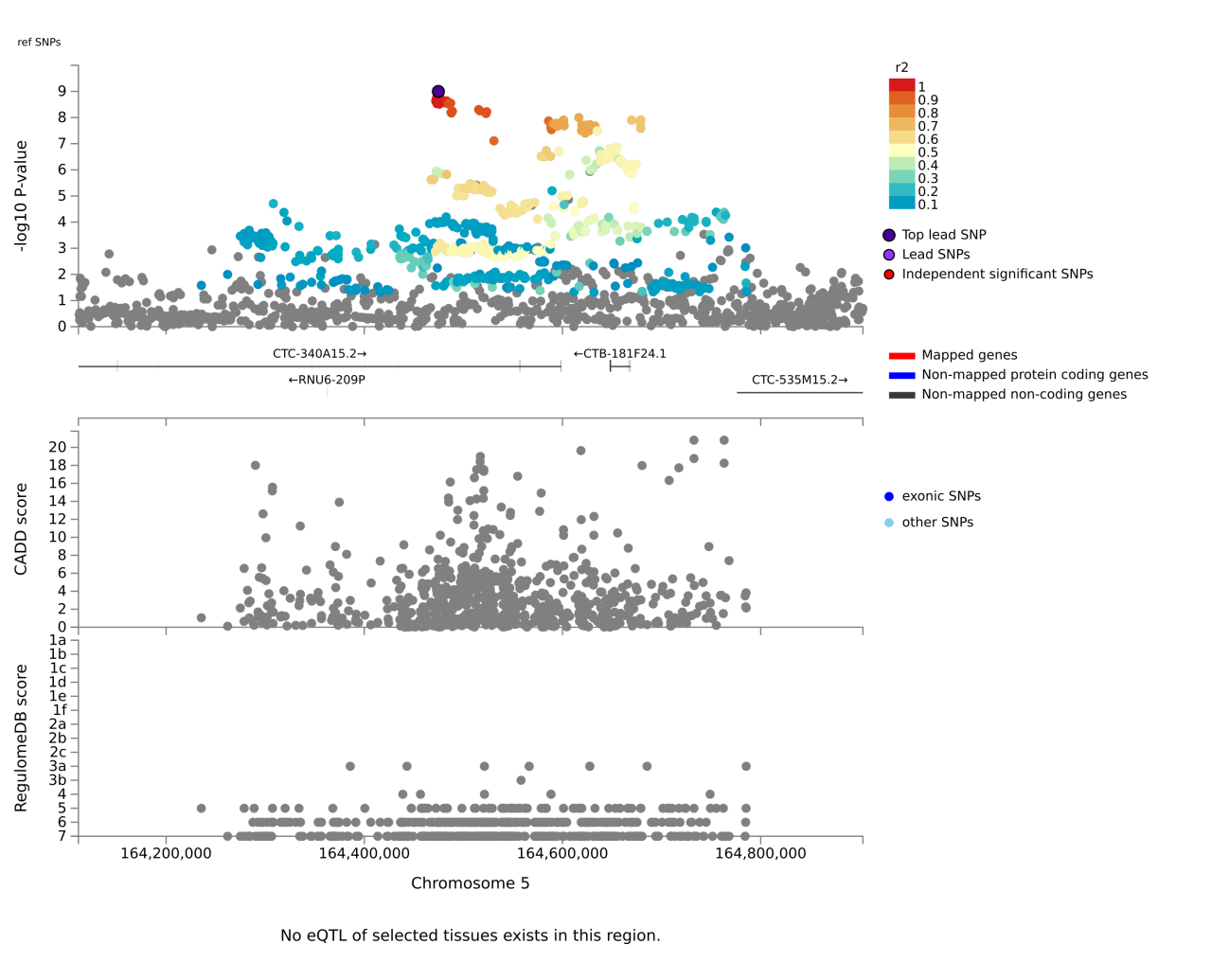

**Chr5: rs180928232**

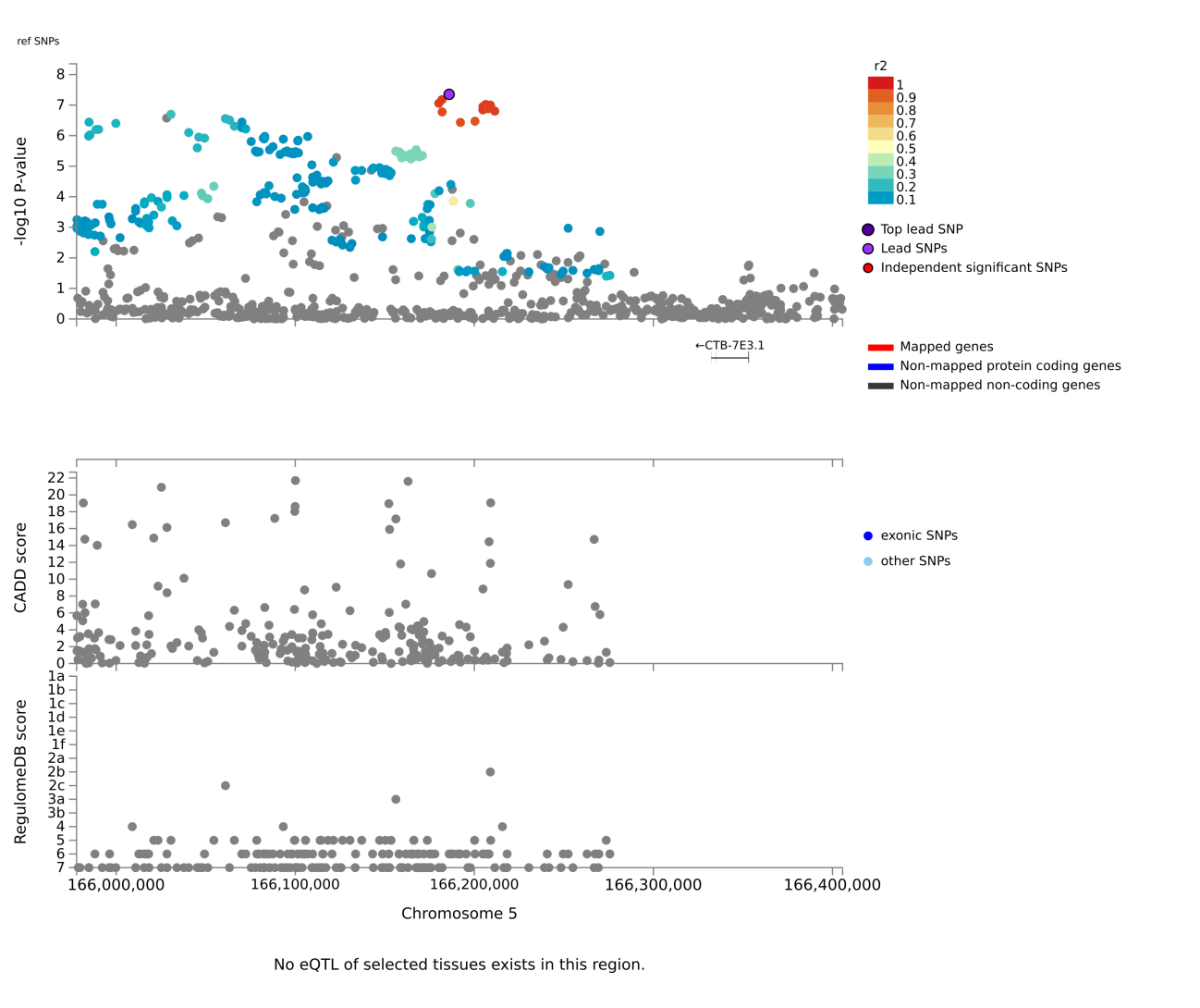

**Chr6: rs200977**

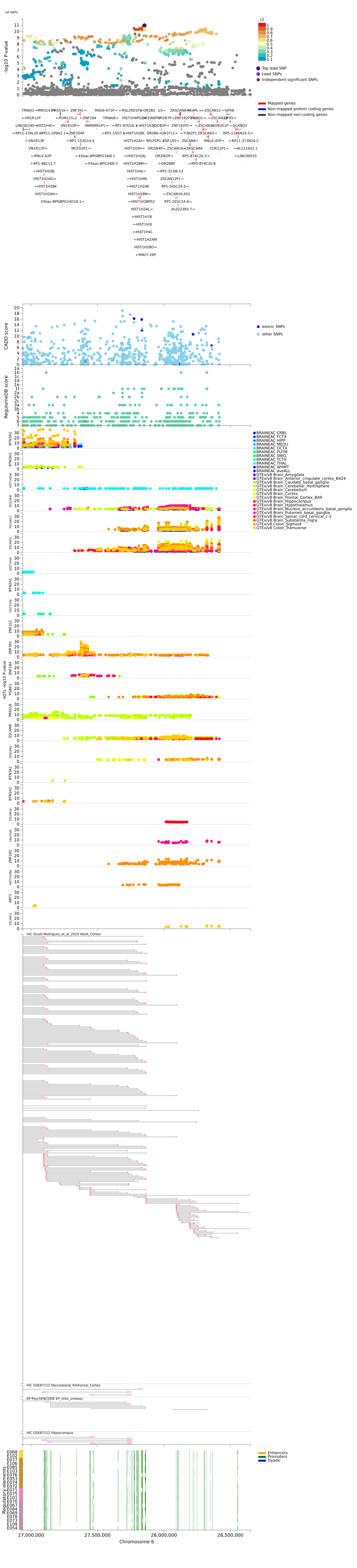

**Chr6: rs2534664**

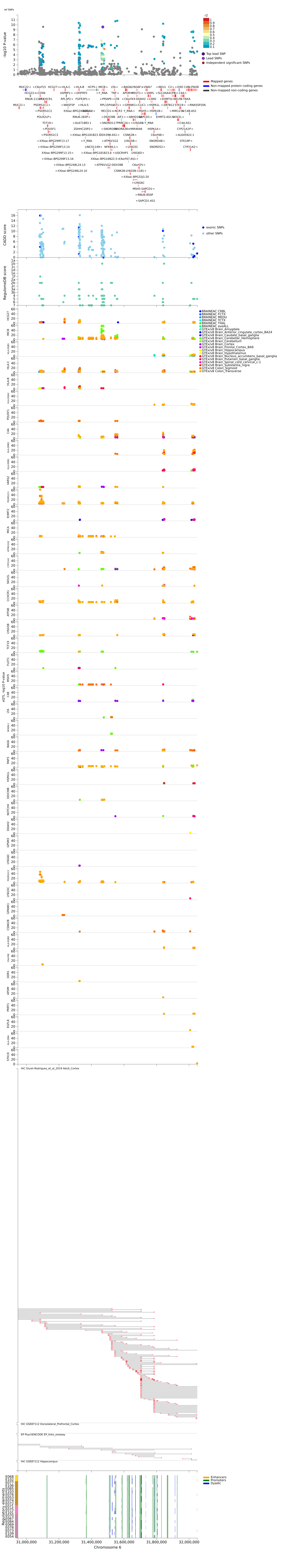

**Chr6: rs1144708**

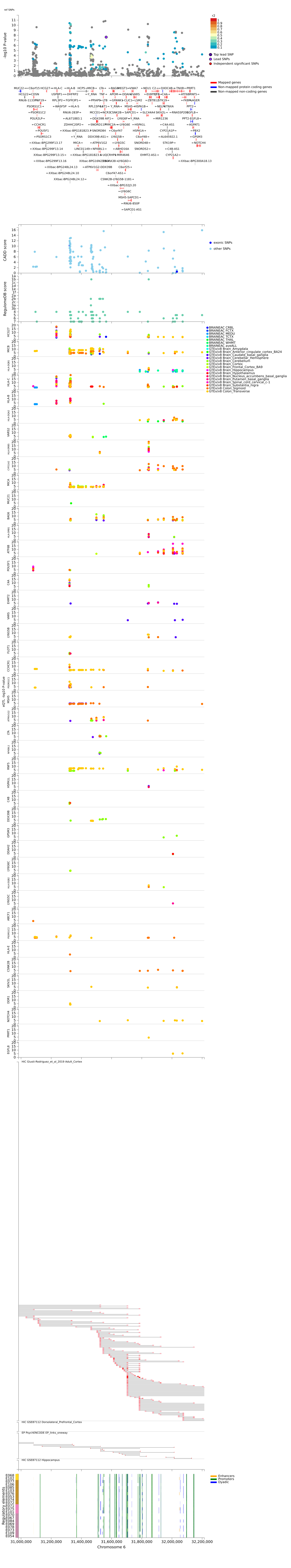

**Chr6: rs12374612**

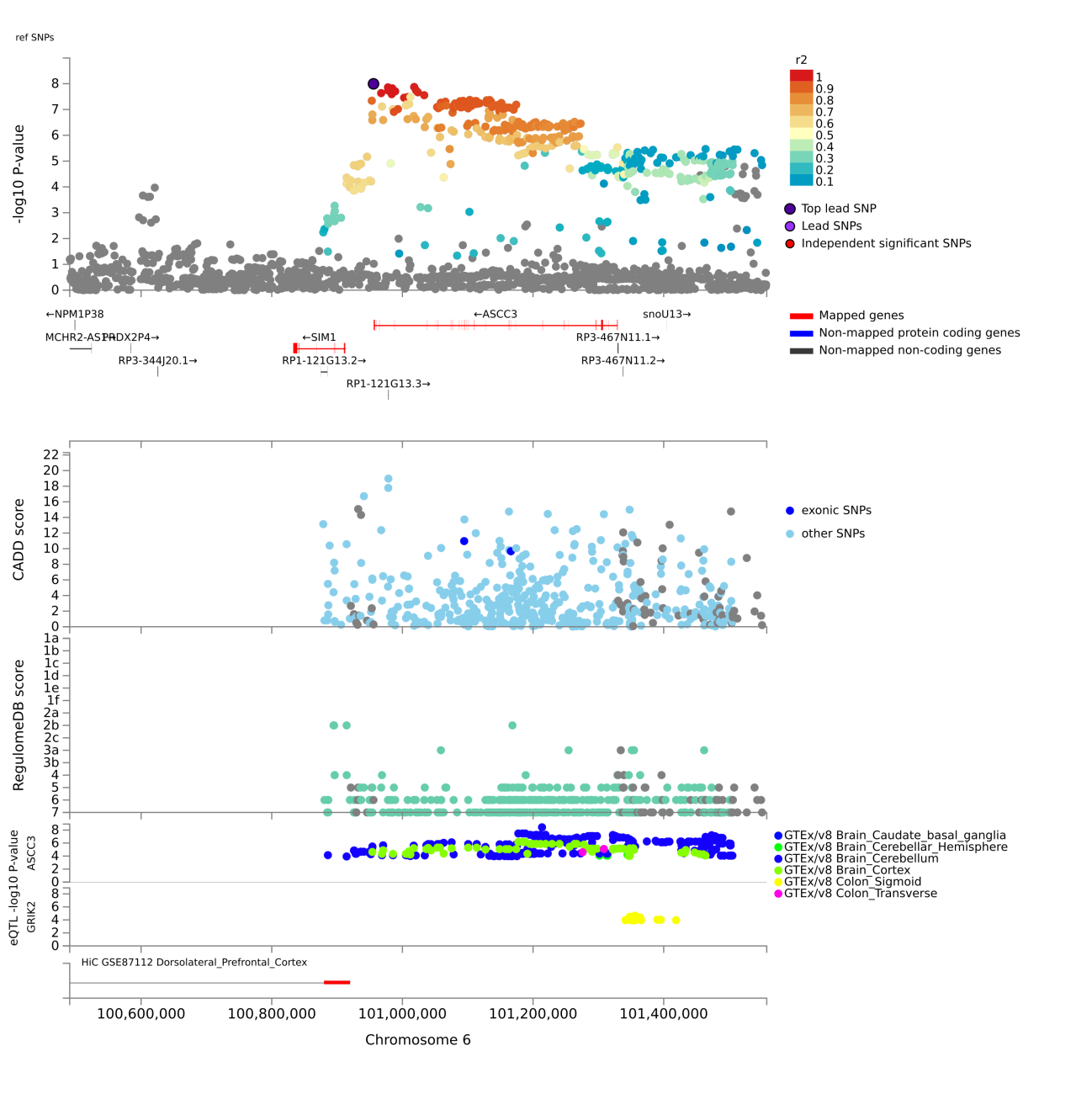

**Chr7: rs2189246**

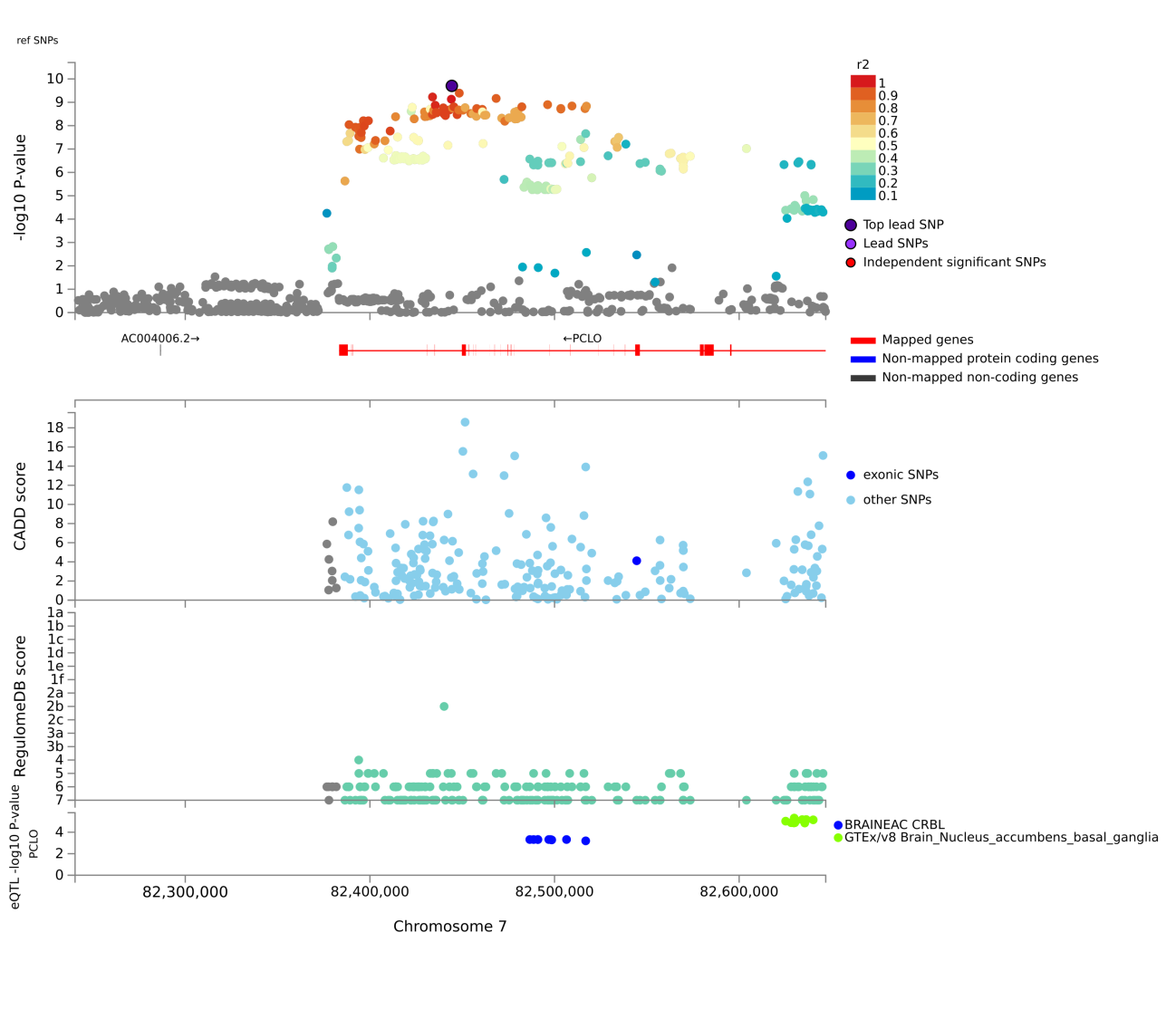

**Chr7: rs6956352**

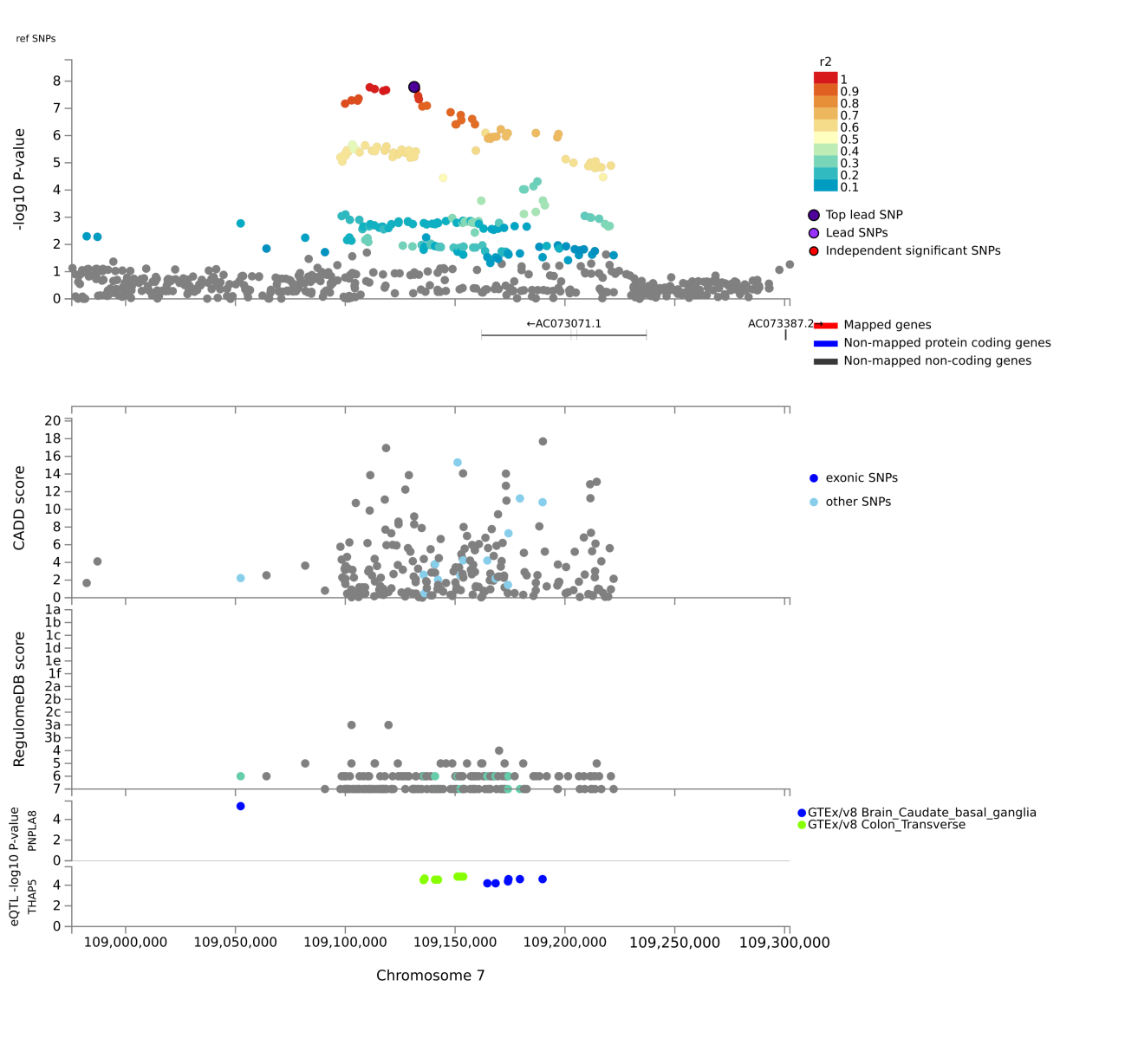

**Chr7: rs4726814**

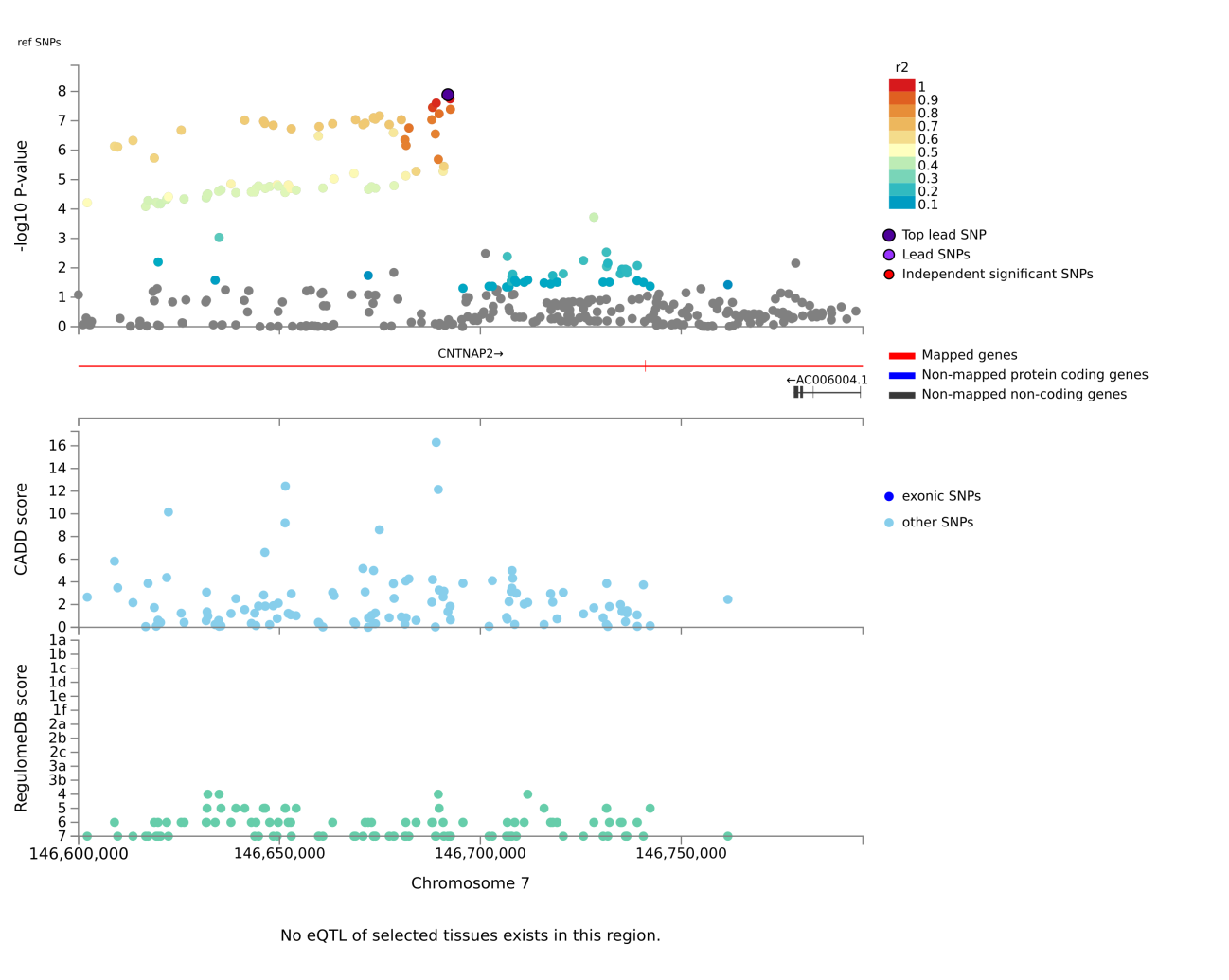

**Chr8: rs4478545**

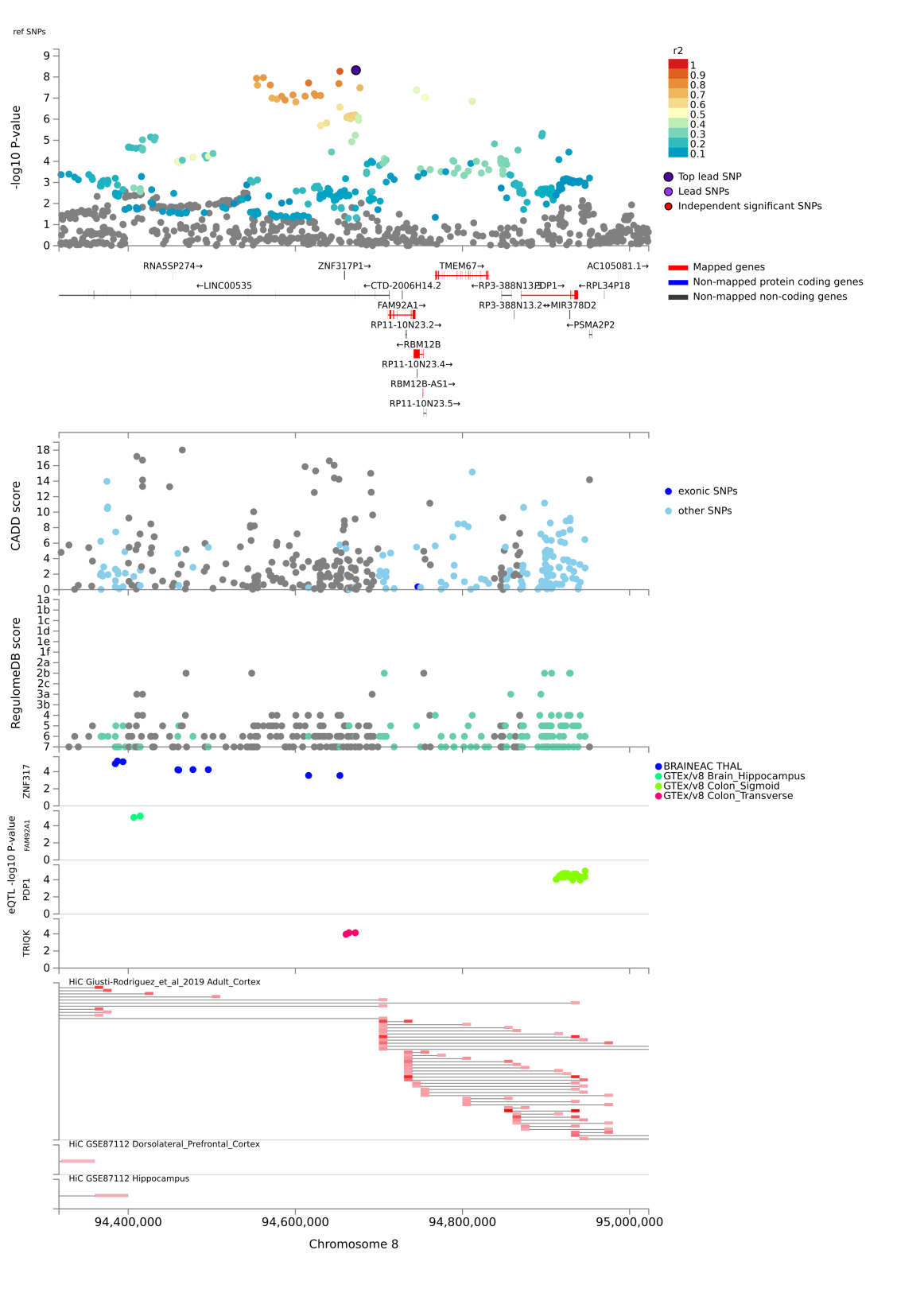

**Chr9: rs3793577**

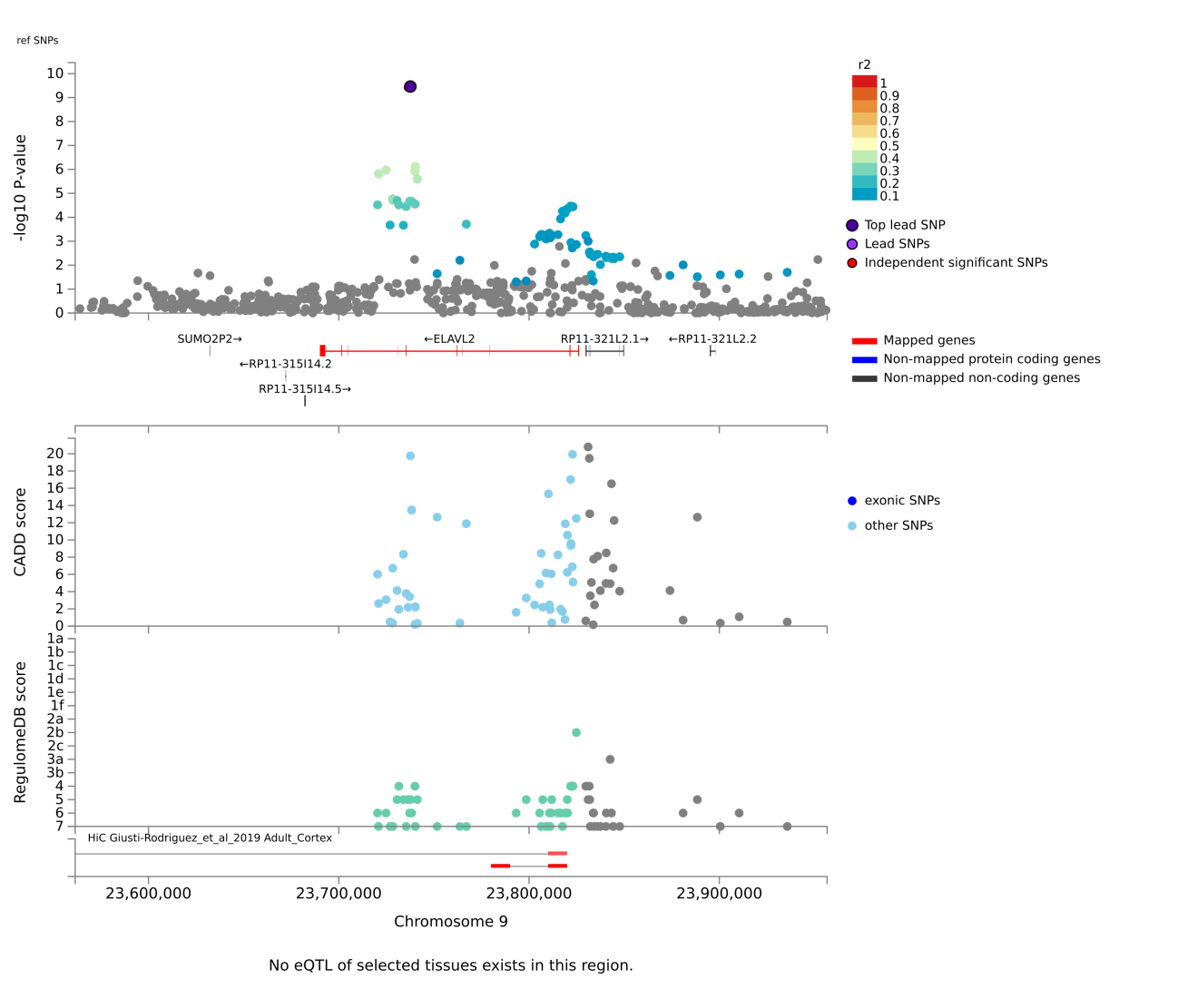

**Chr9: rs4744242**

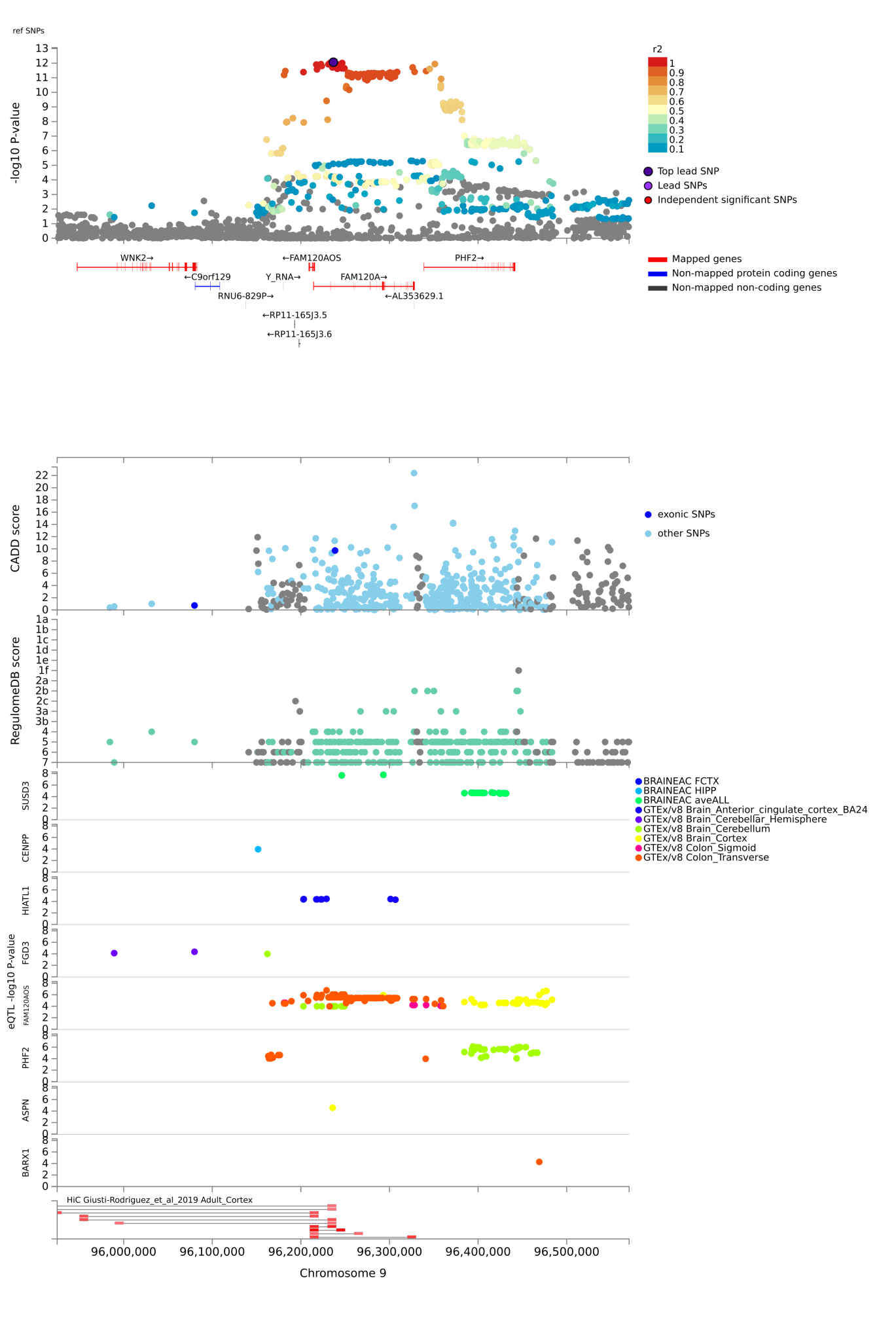

**Chr9: rs10123941**

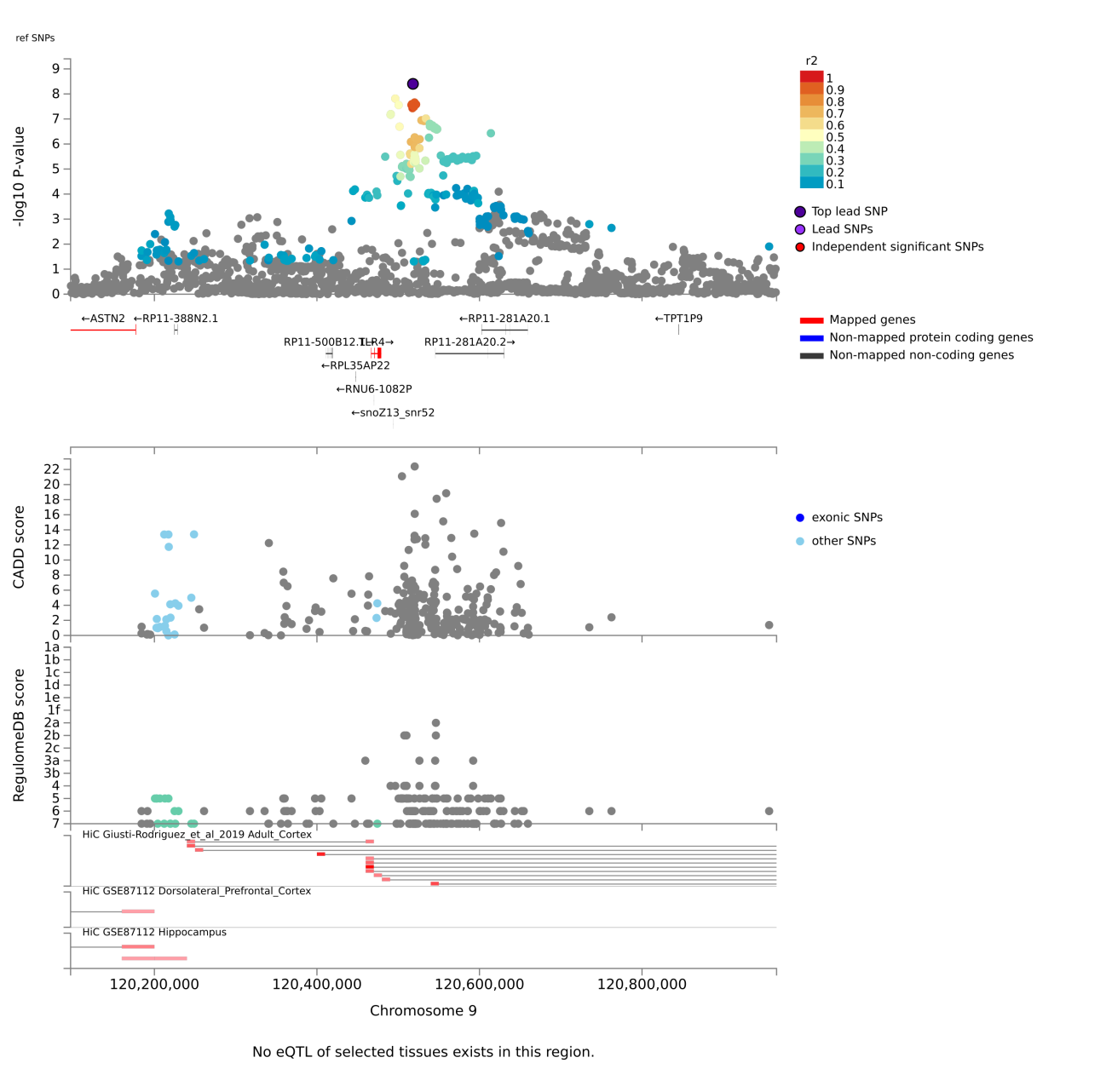

**Chr10: rs6584631**

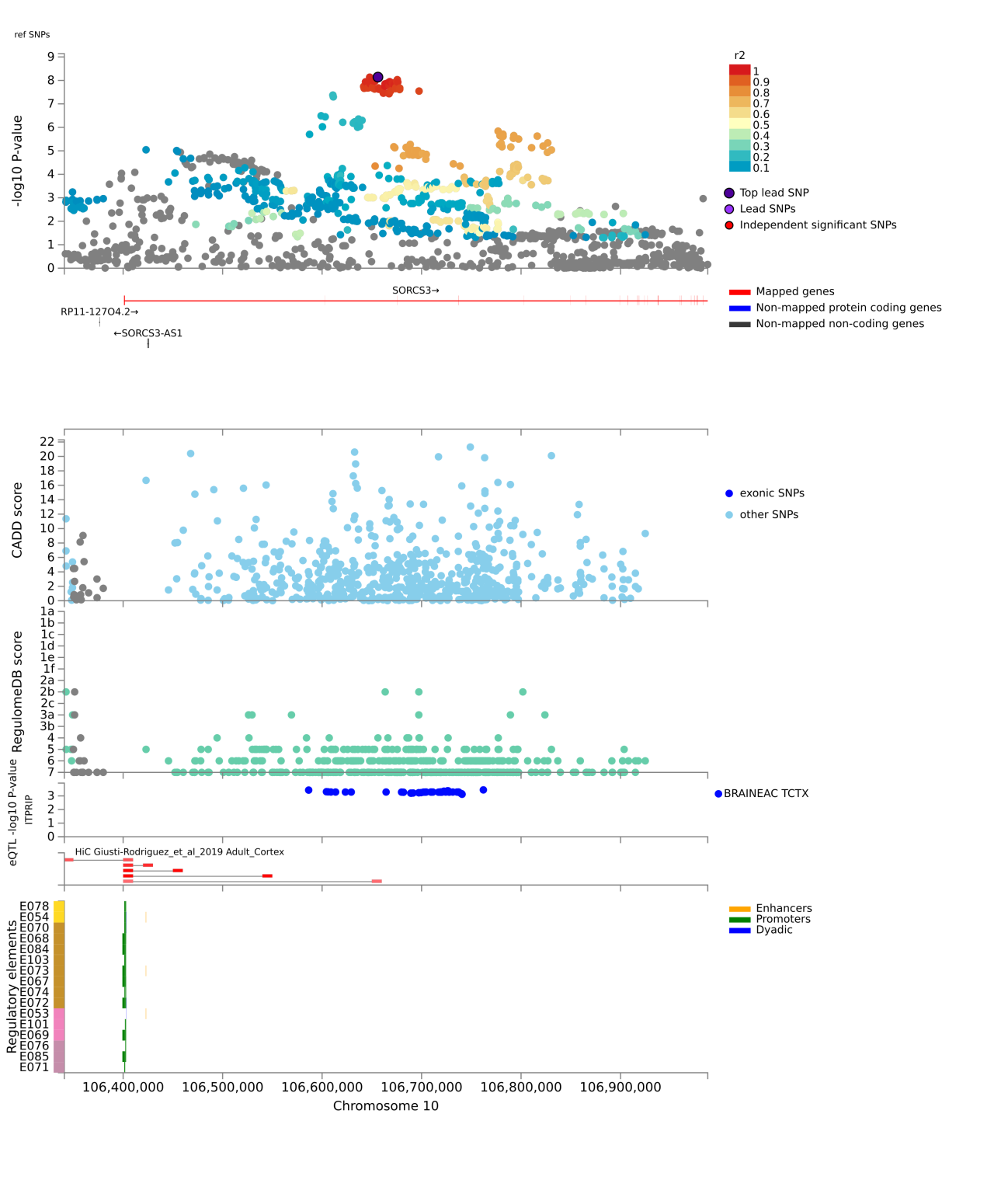

**Chr11: rs4937872**

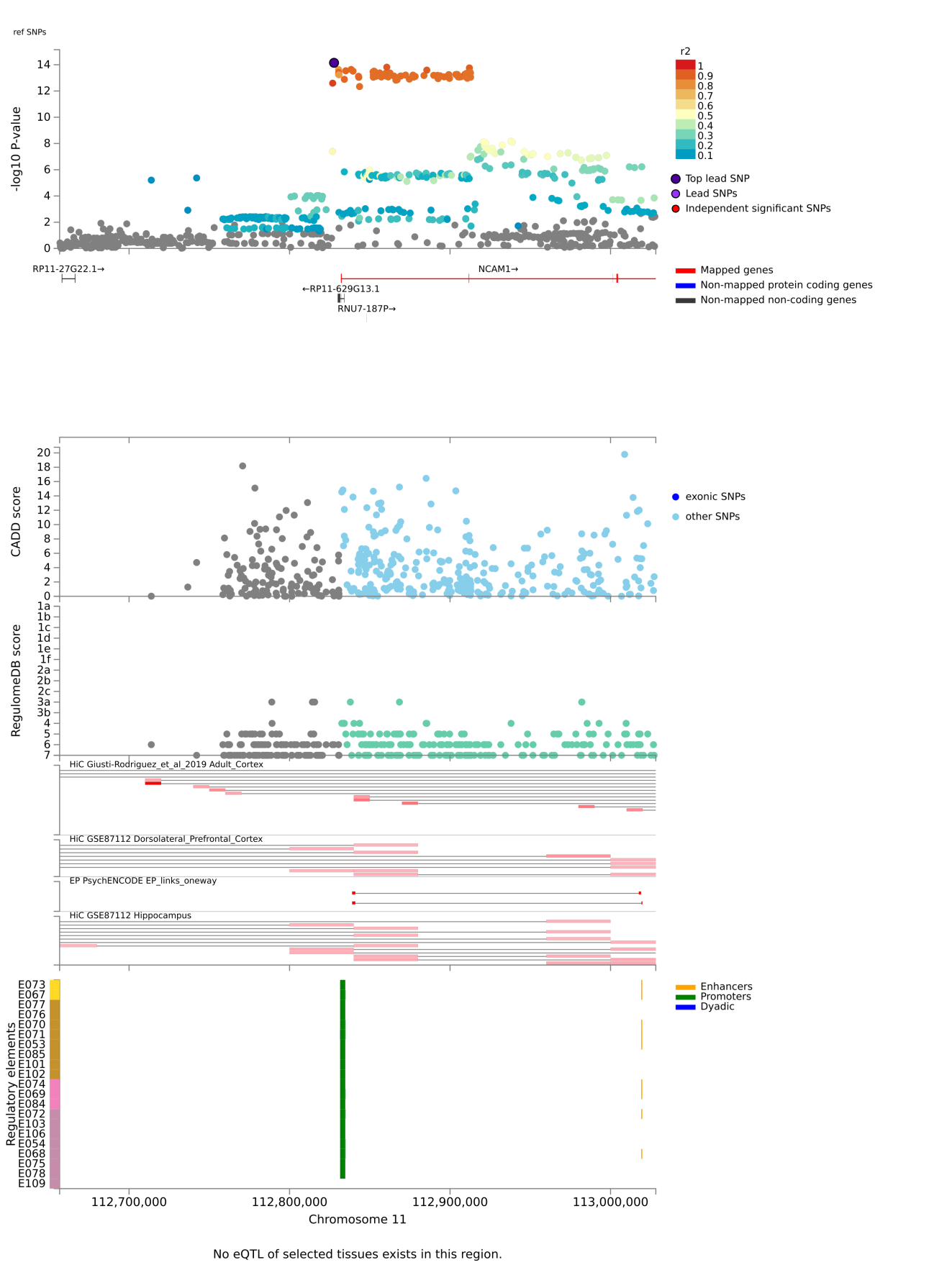

**Chr13: rs9530139**

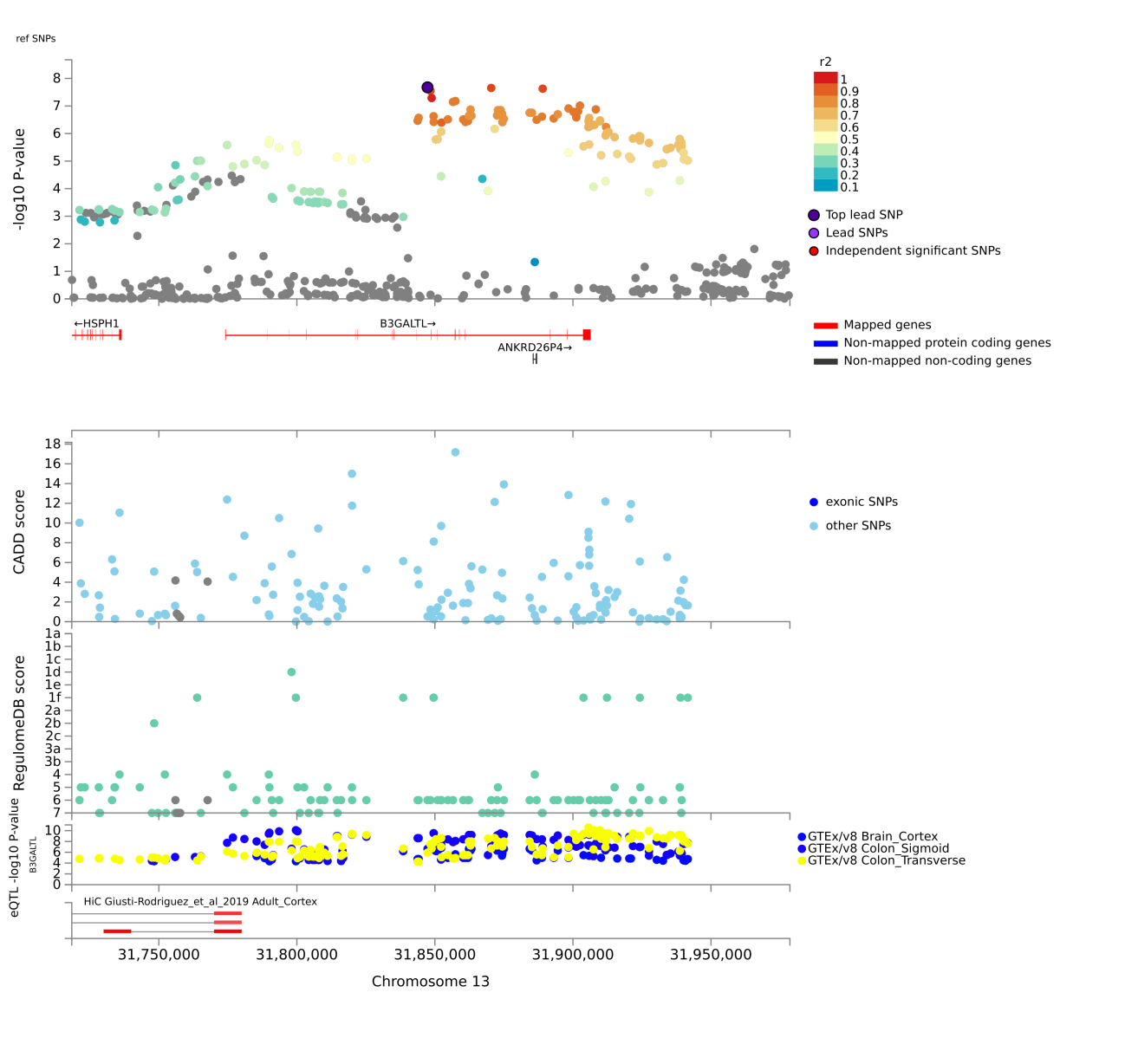

**Chr13: rs9597797**

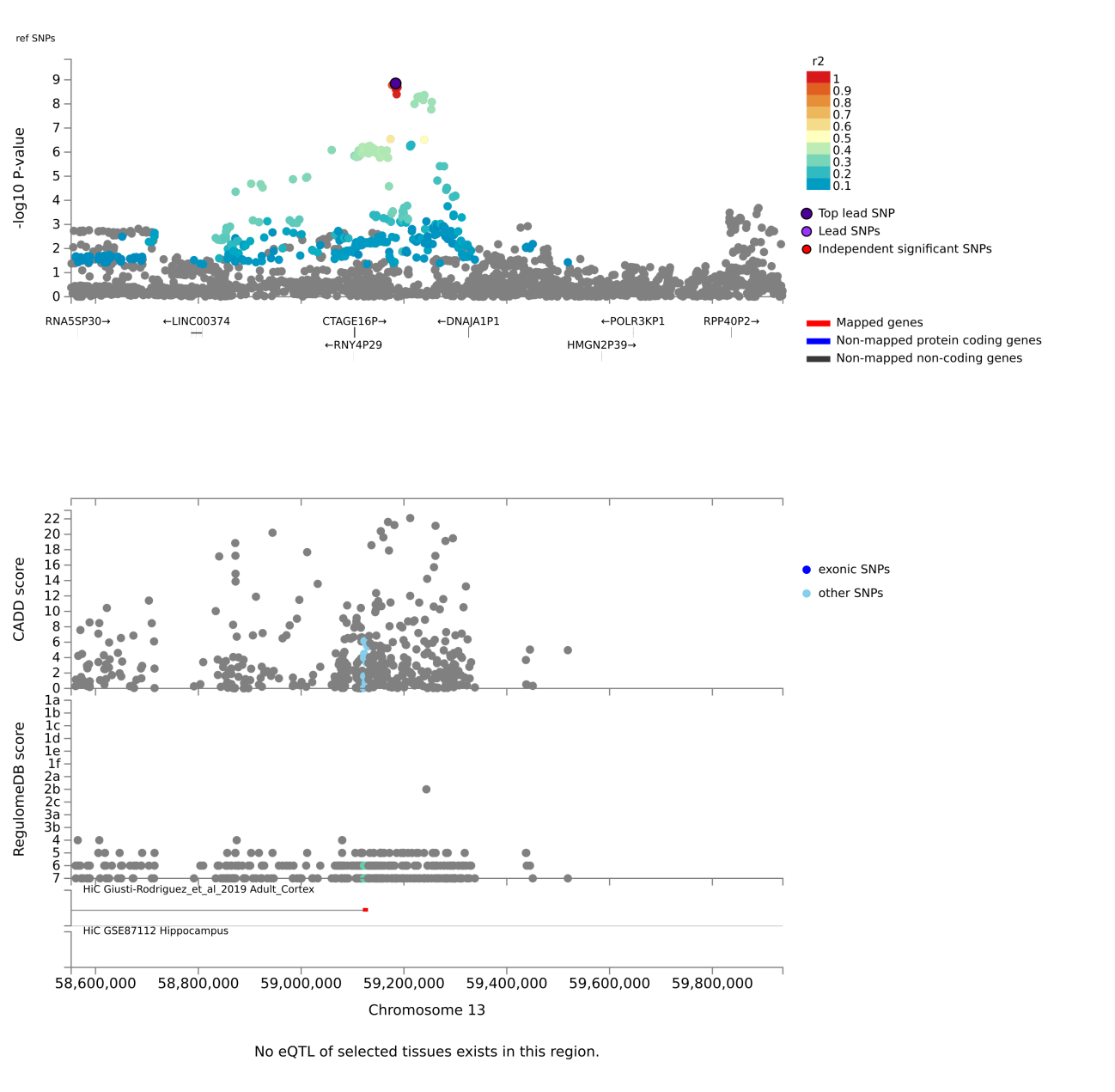

**Chr14: rs2121708**

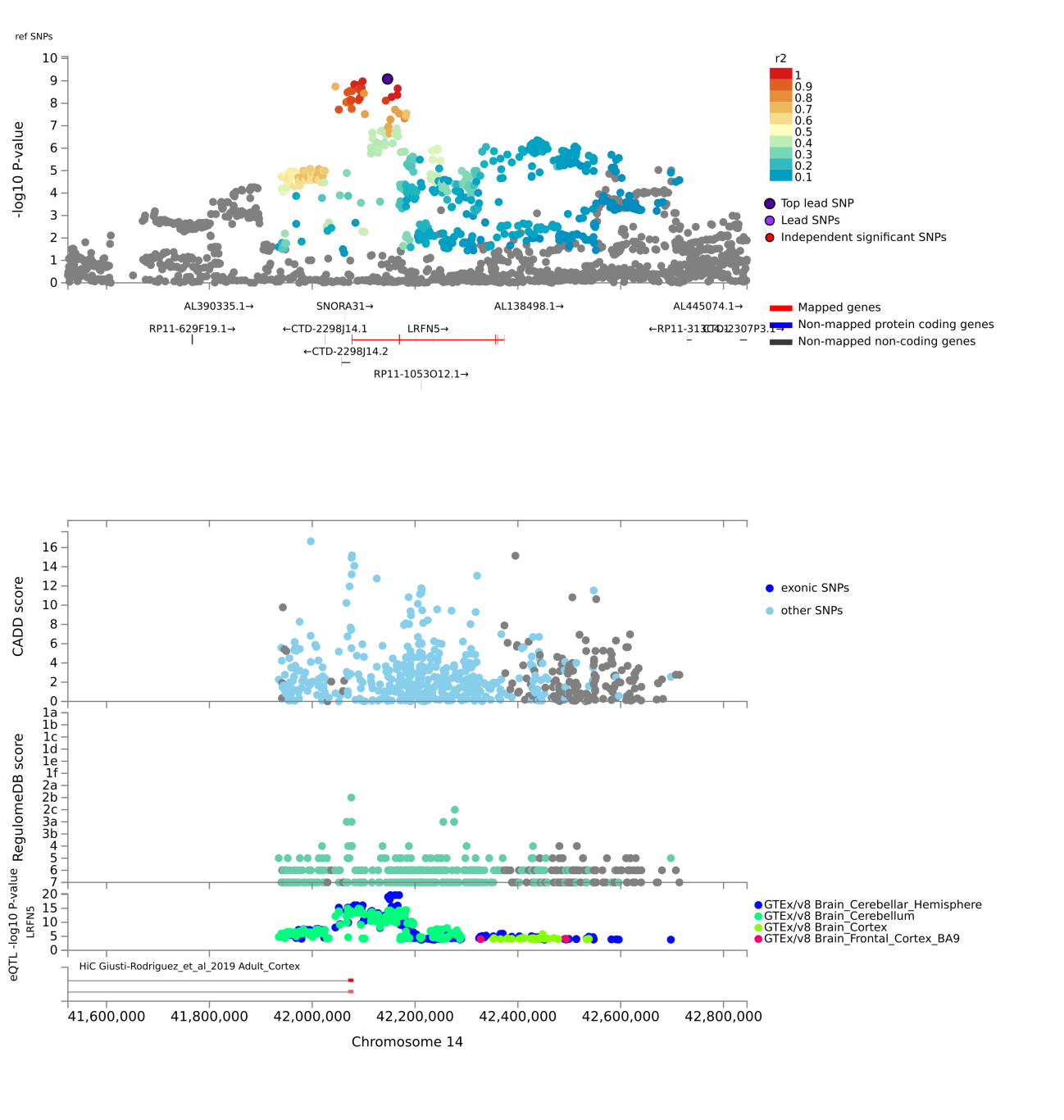

**Chr14: rs35641442**

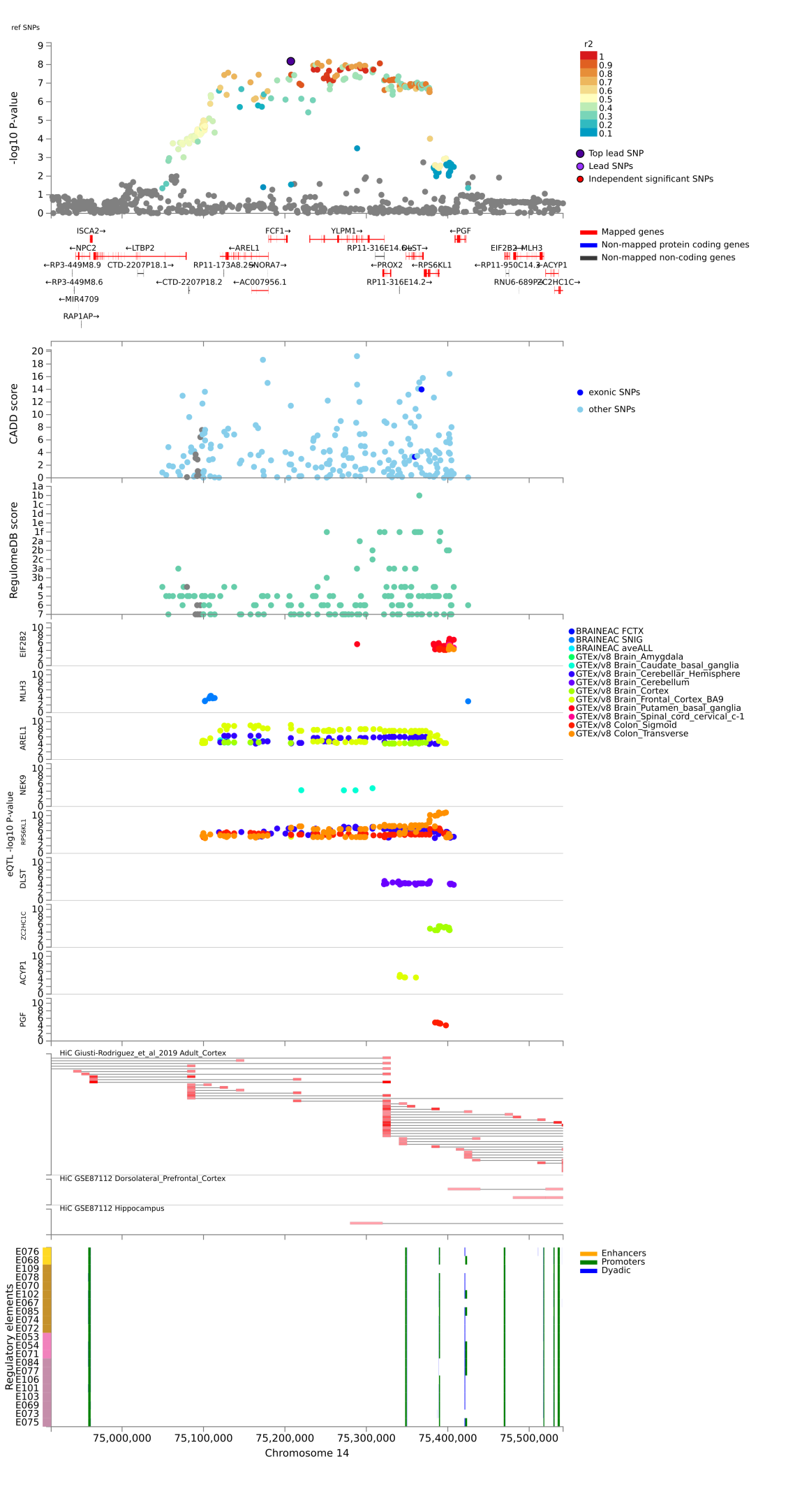

**Chr16: rs1862743**

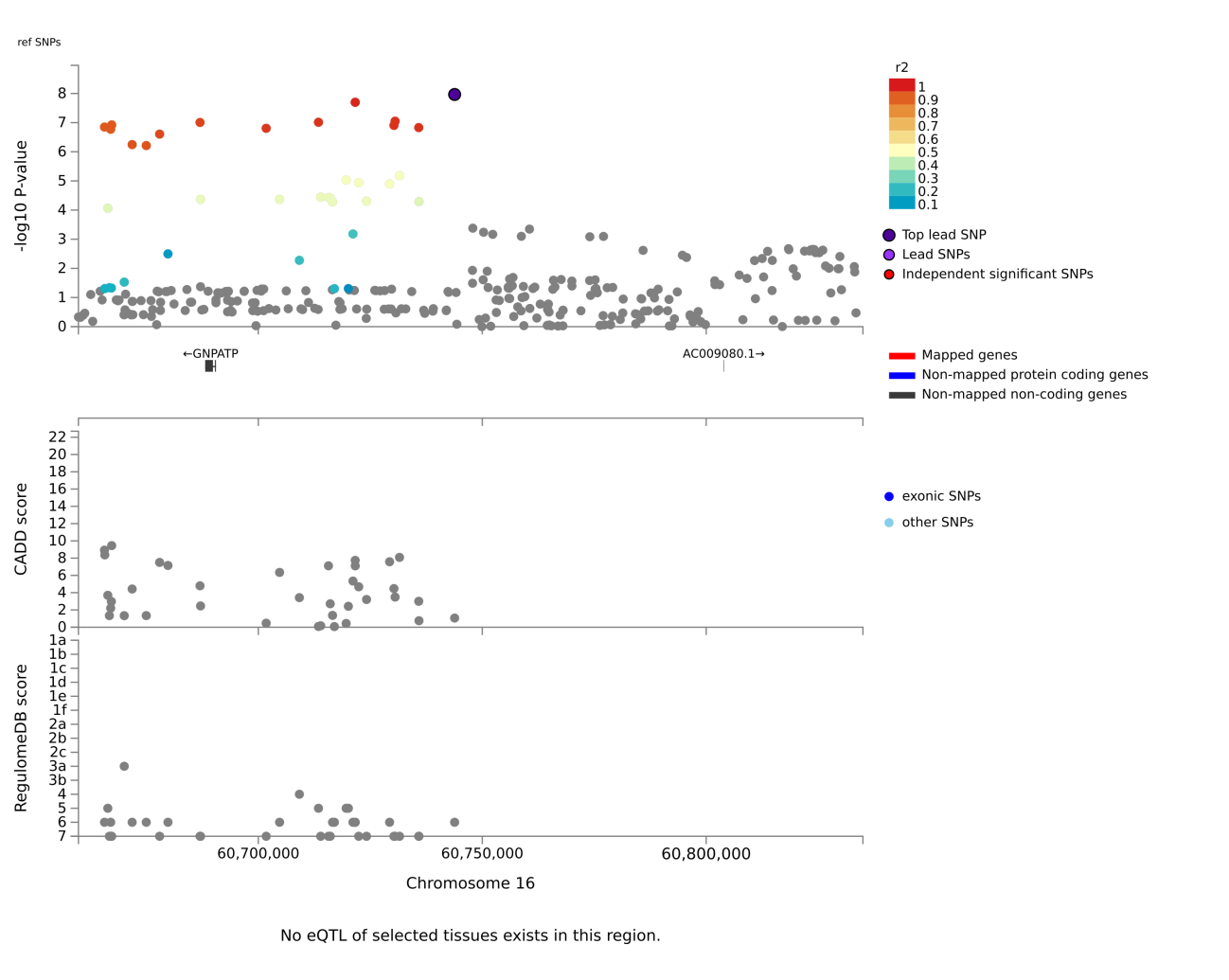

**Chr18: rs2978362**

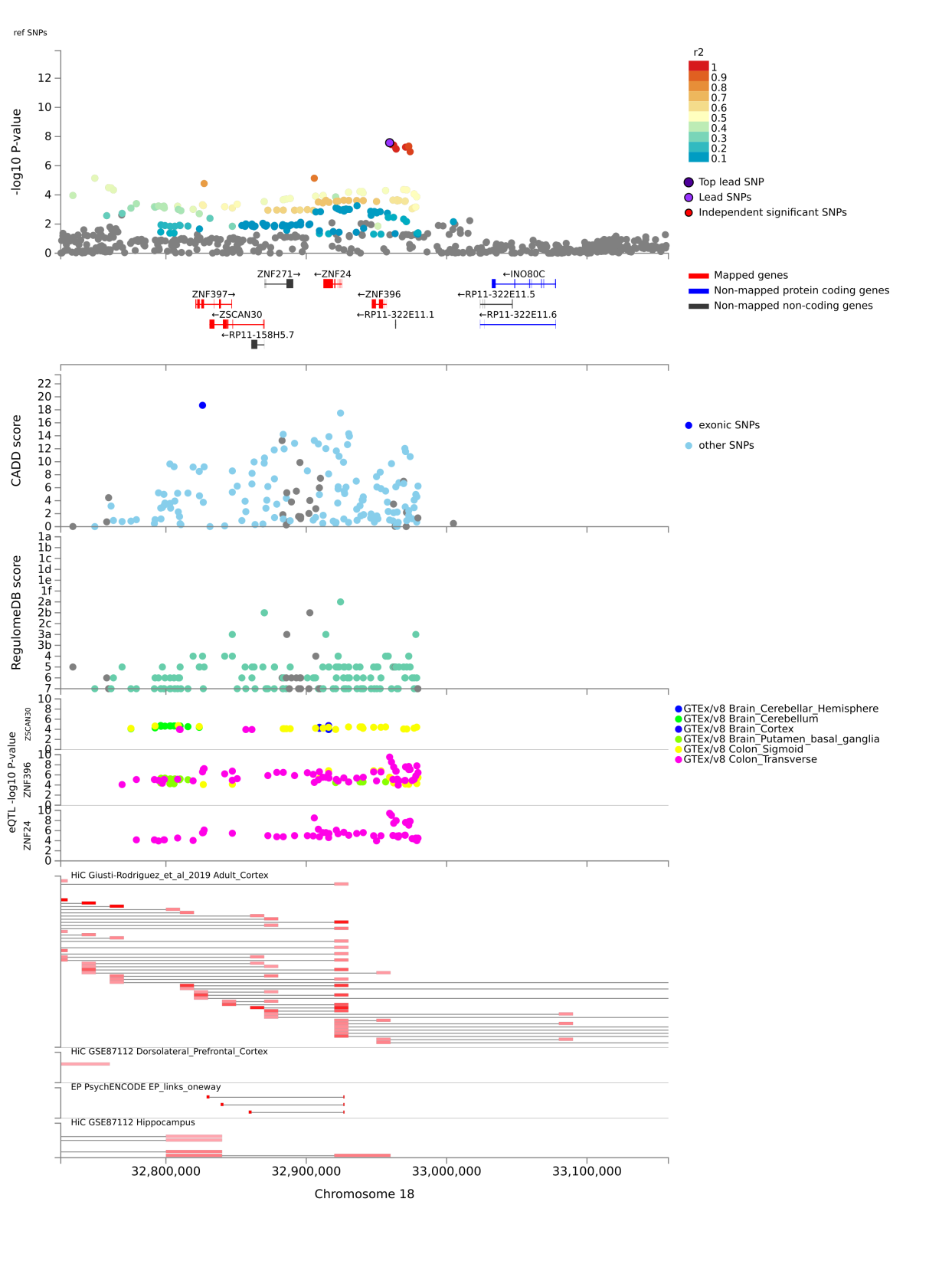

**Chr18: rs11877758**

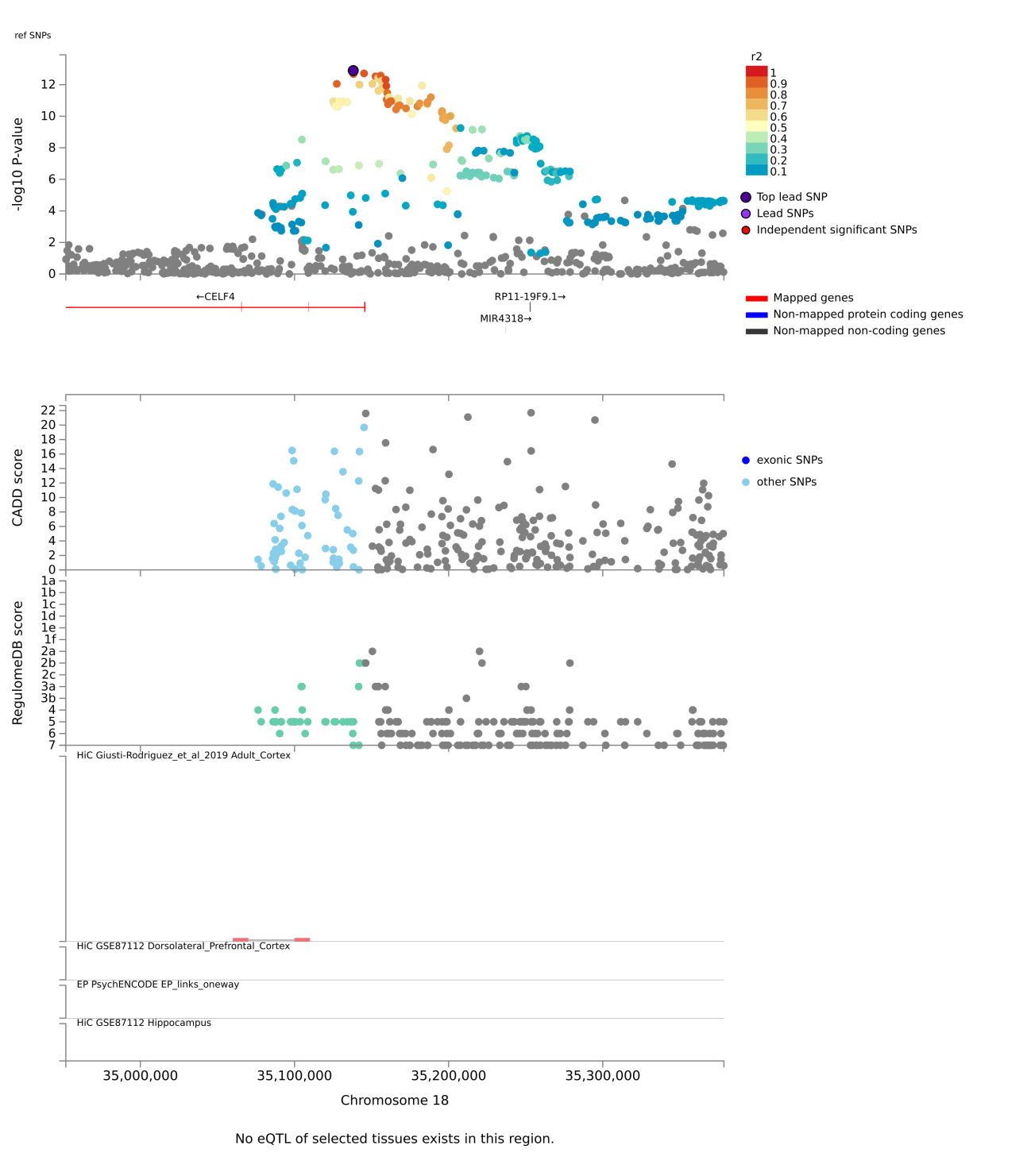

**Chr18: rs17410557**

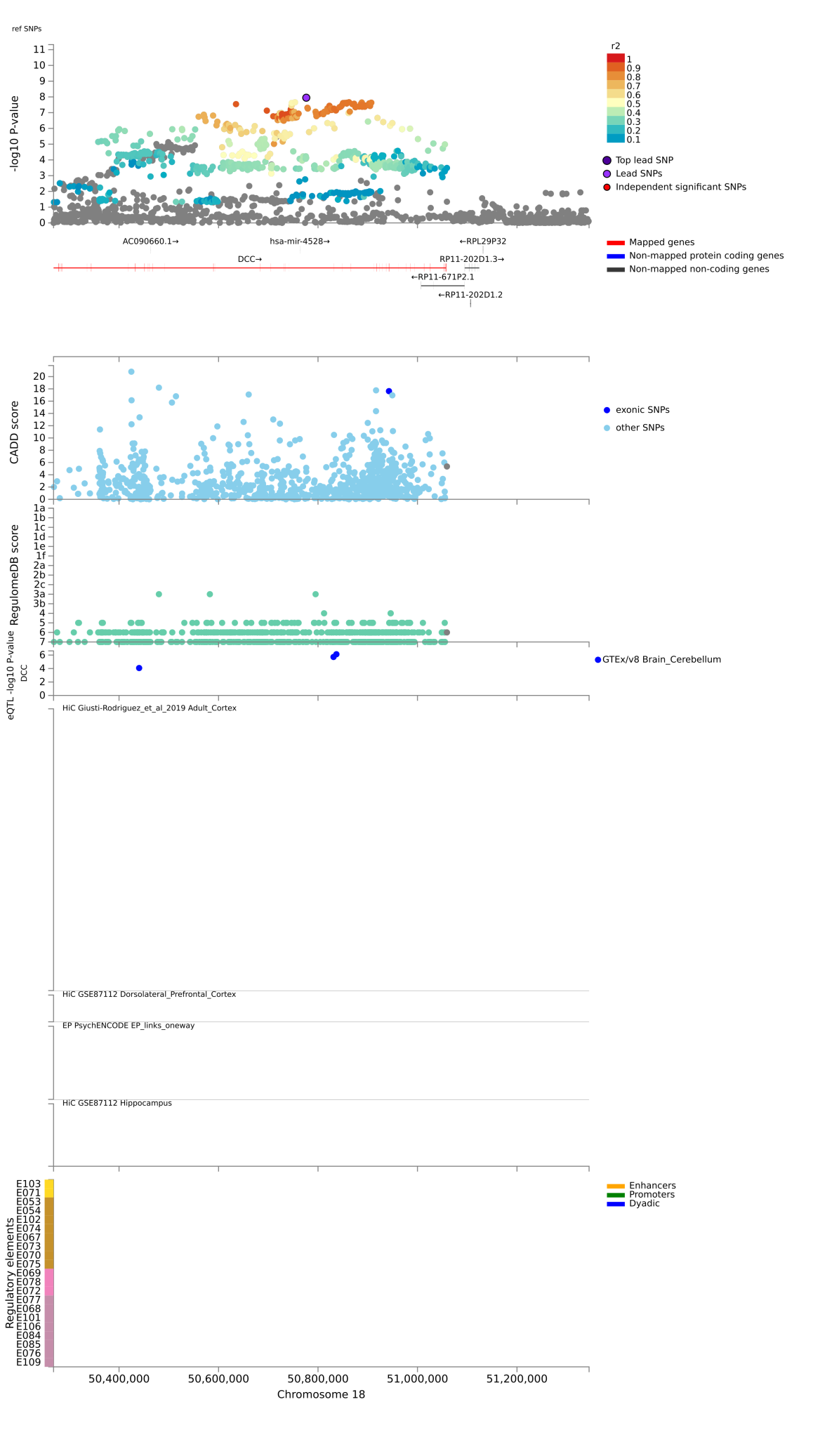

**Chr18: rs12958048**

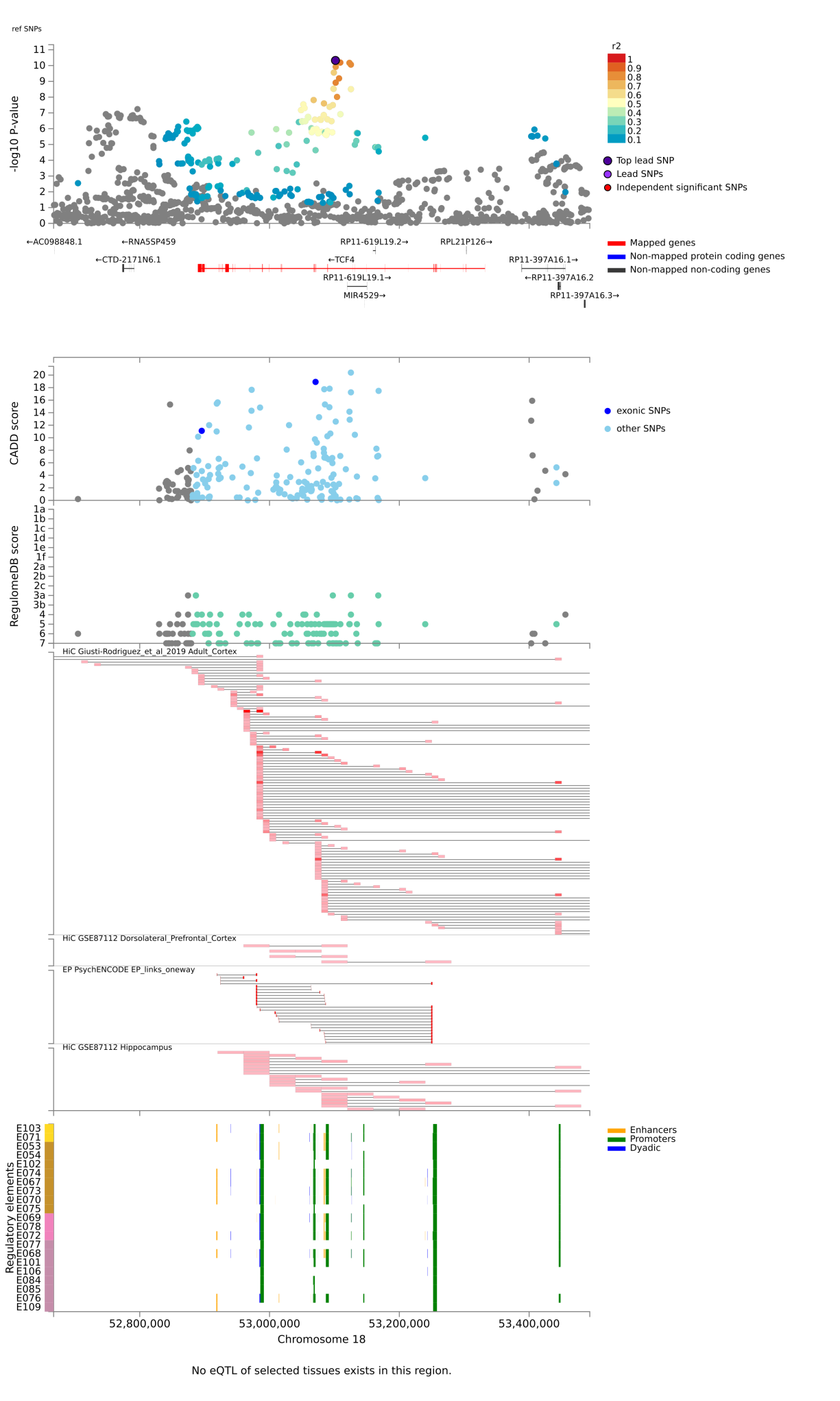

**Chr19: rs2111530**

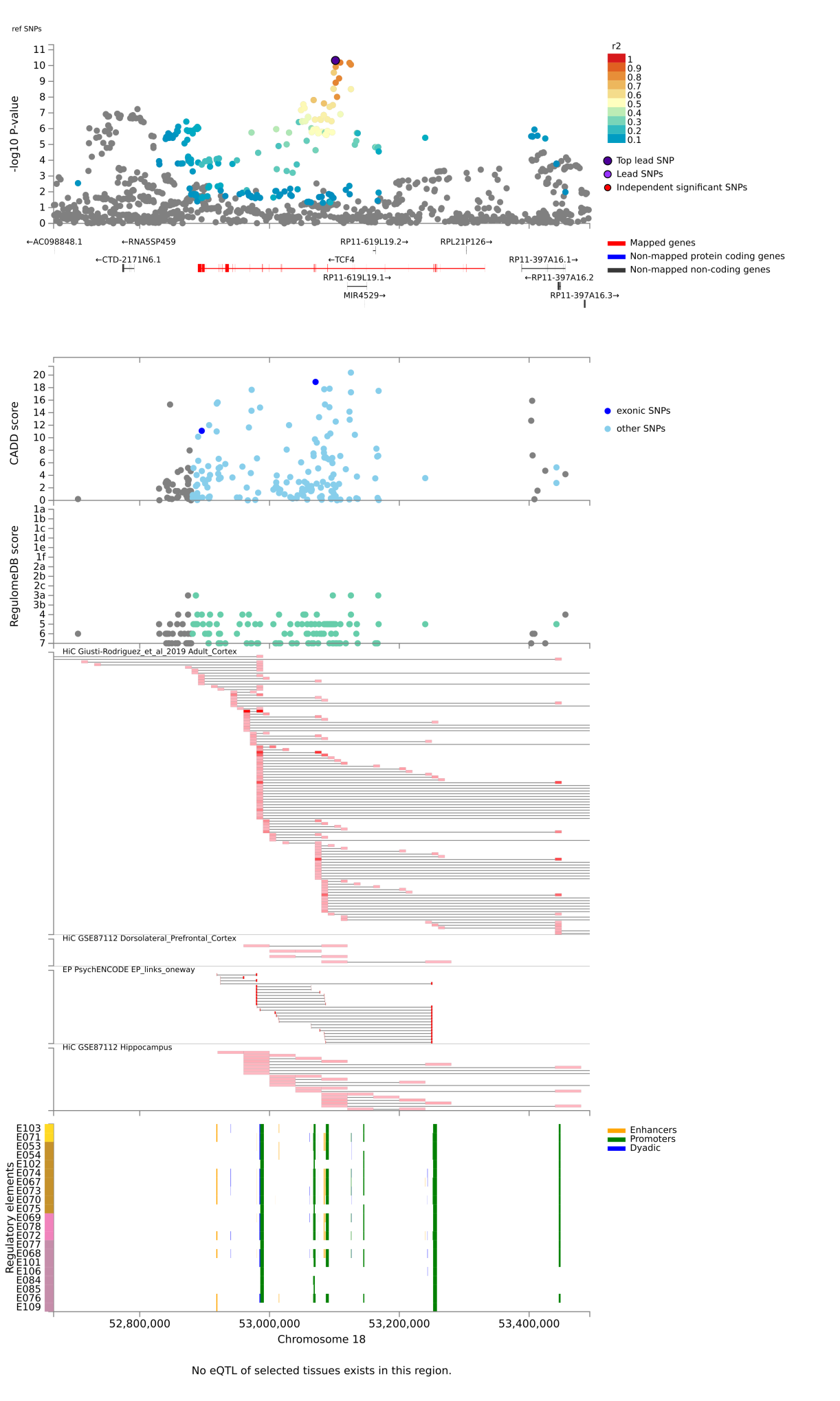

**Chr20: rs2024568**

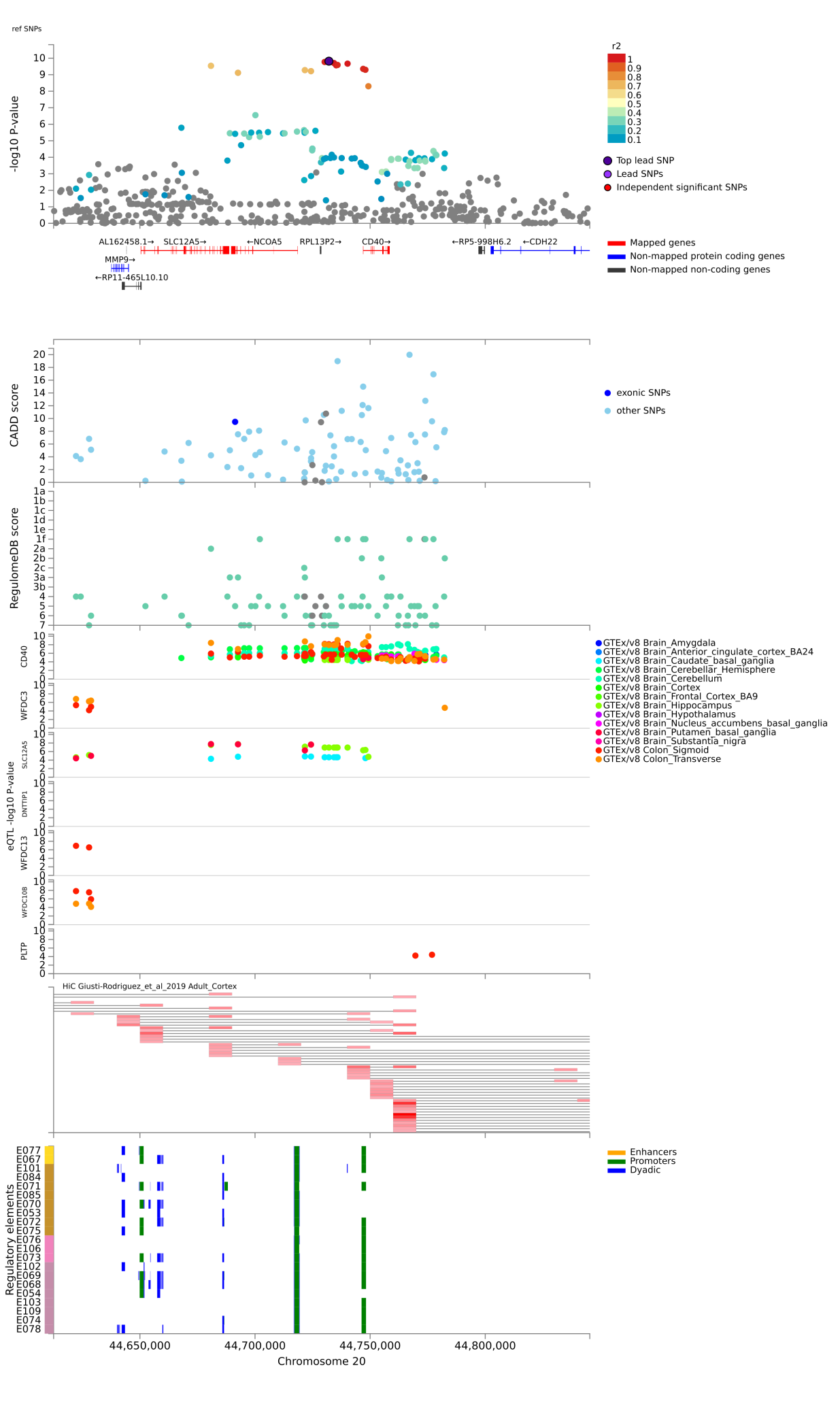

**Chr22: rs11090039**

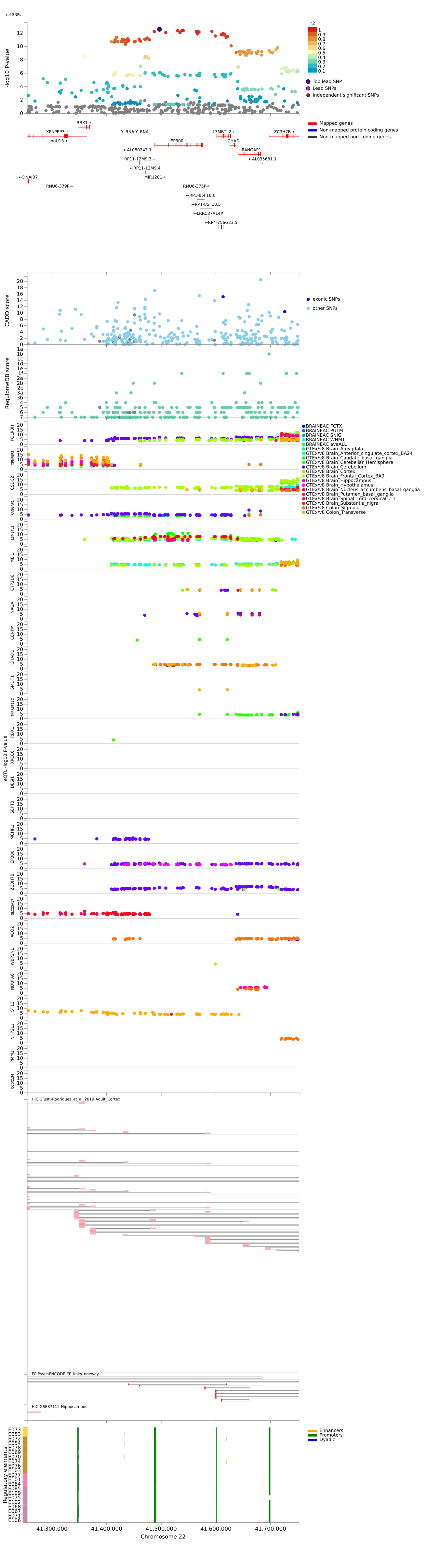

### **Supplementary Figure 2.** Regional Plots of the 42 lead SNPs identified in the MTAG-IBS analysis. In red, genes mapped by SNPs in the credible sets based on physical proximity, chromatin interaction and/or eQTLs using FUMA.

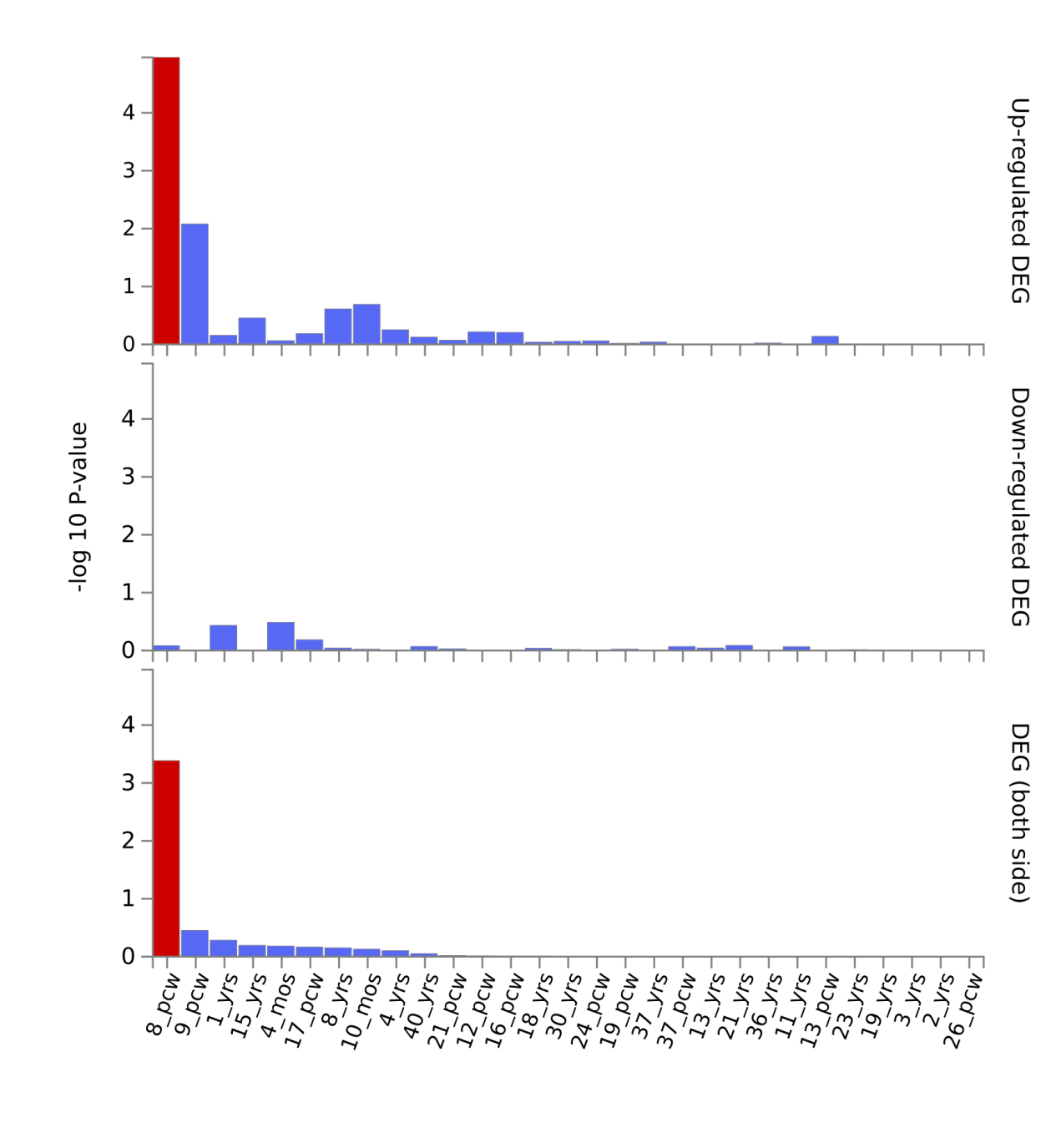
**Supplementary Figure 3. Enrichment of genes mapped to MTAG-IBS variants with credible sets on Differentially Expressed Genes (DEG) in brain tissue.**

Results from hypergeometric test evaluating enrichment of the 289 mapped genes by credible variants in DEG in brain tissue representing different brain developmental stages in BrainSpan. Significant enrichment at Bonferroni corrected P-value ≤ 0.05 are coloured in red.

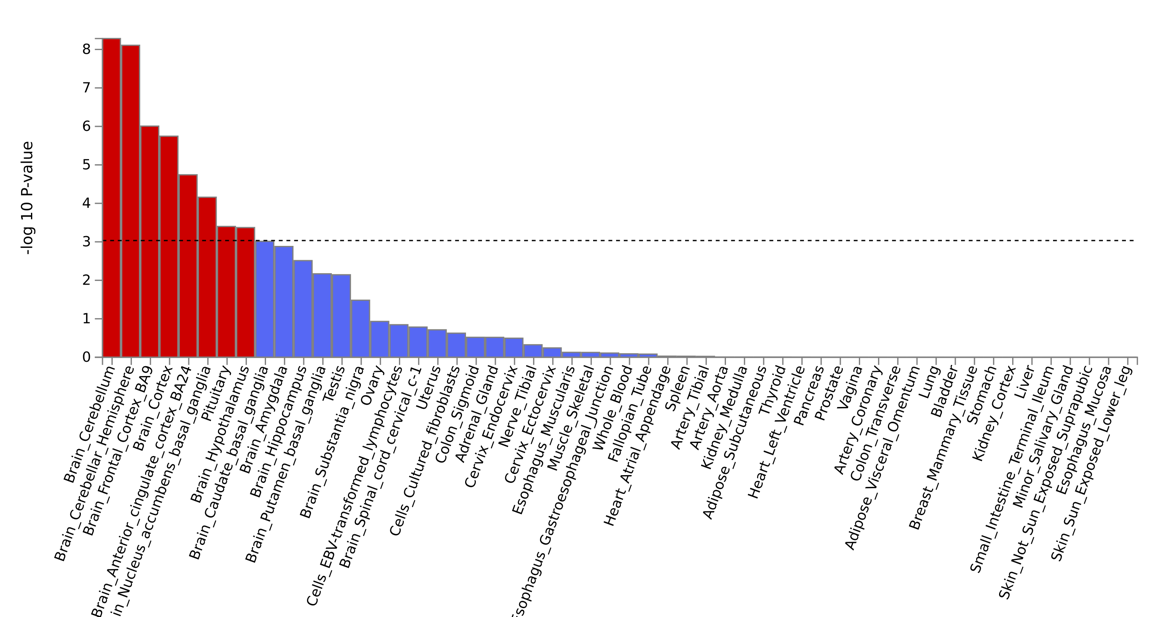
(A)

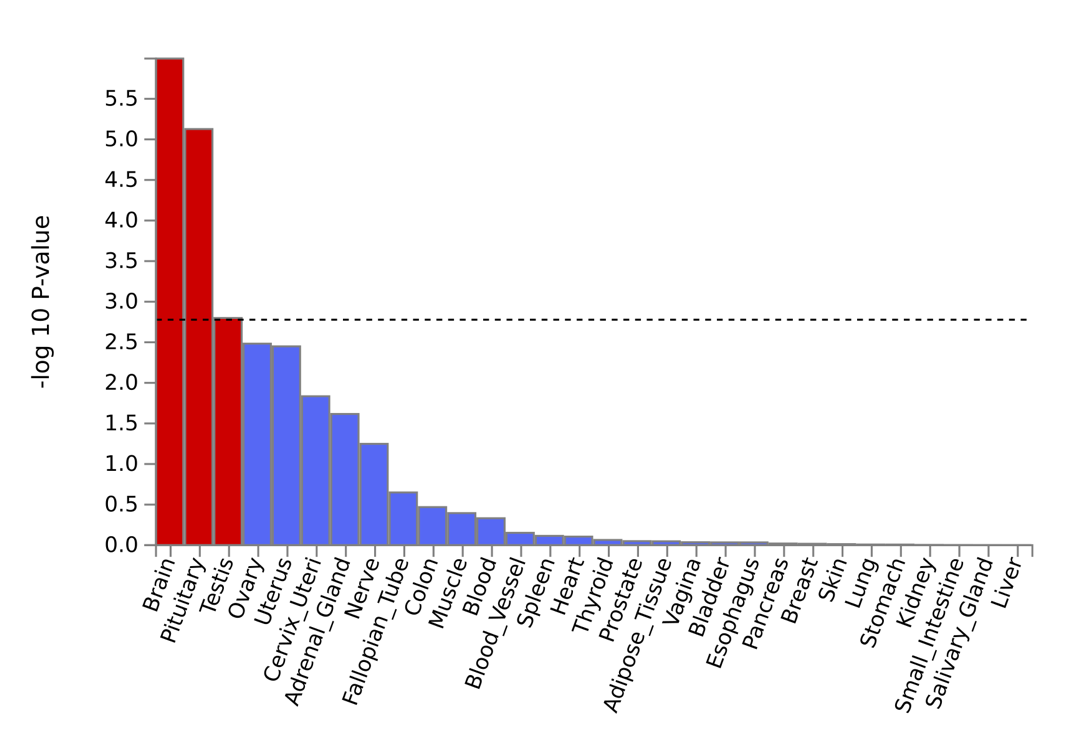
(A)

**Supplementary Figure 4.** **MAGMA tissue expression analysis using GTEx v.8.**

Results from MAGMA gene-property analysis between gene-based MTAG-IBS associations and tissue specific gene expression profiles. (A) GTEx v.8 54 tissues. (B) GTEx v.8 30 general tissues. Red bars indicate significant results.

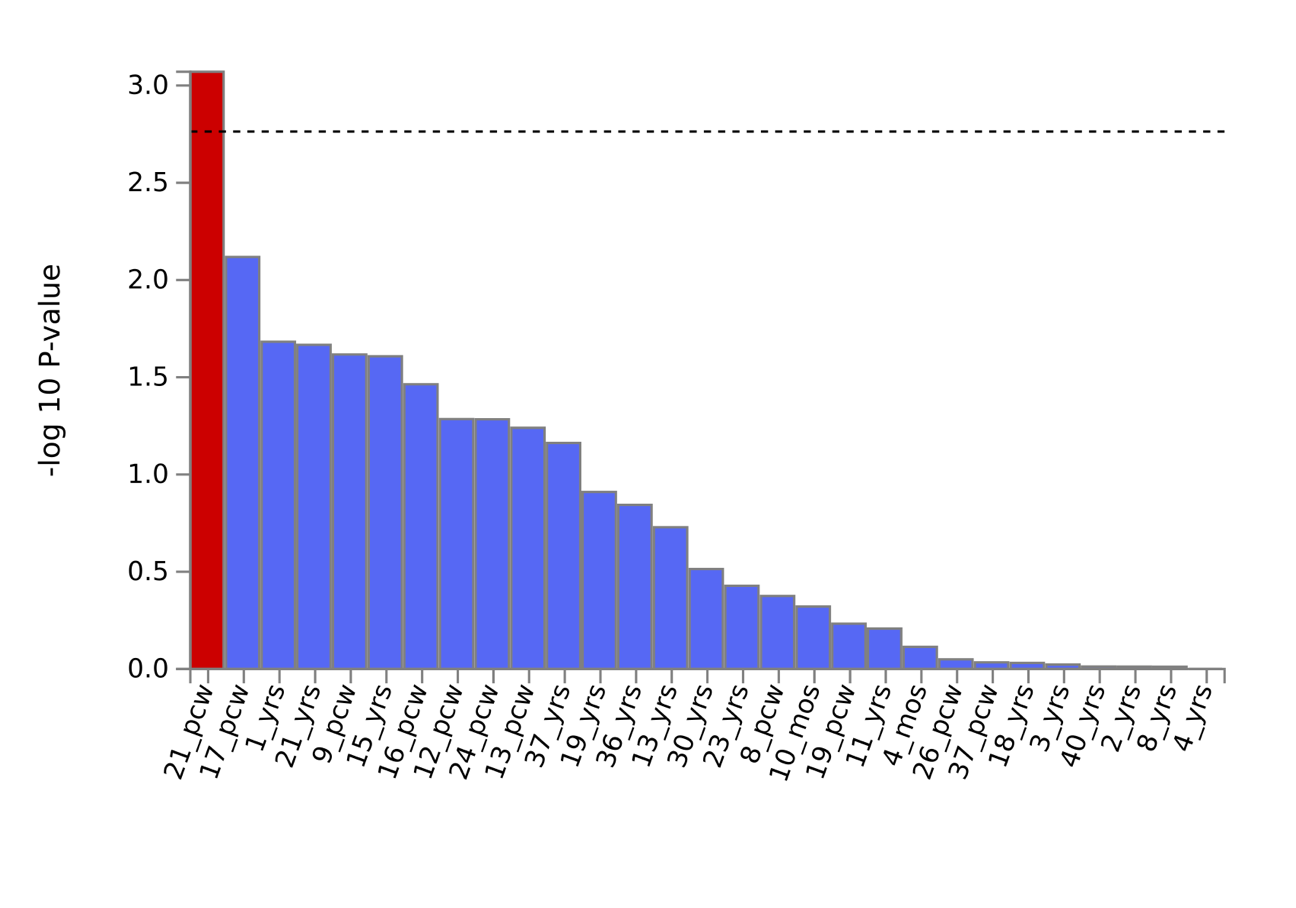
(A)

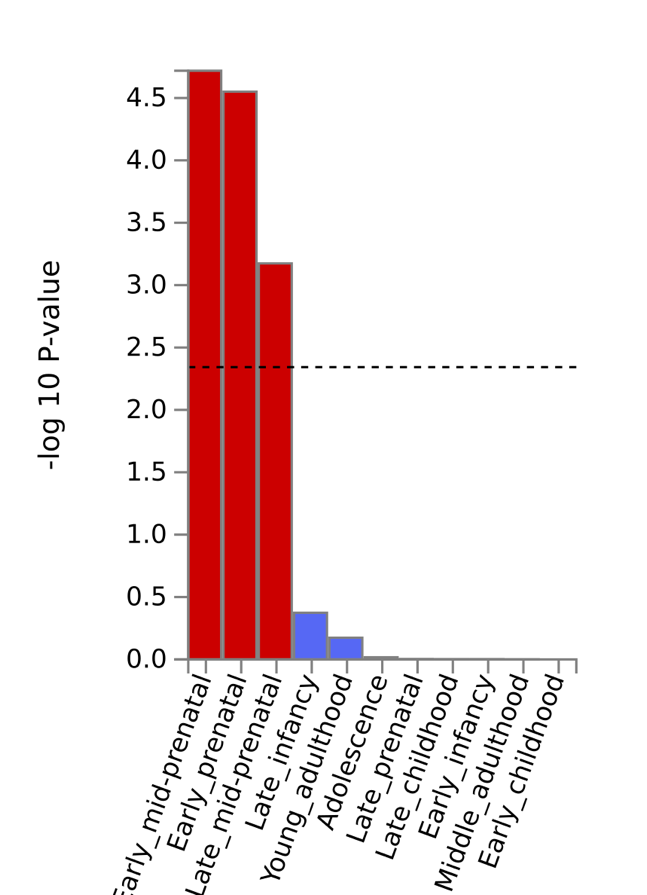

(B)

### **Supplementary Figure 5.** **MAGMA tissue expression analysis using Brainspan.**

Results from MAGMA gene-property analysis between gene-based MTAG-IBS results and tissue specific gene expression profiles in Brainspan. (A) BrainSpan 29 ages. (A) Brainspan 11 developmental stages. Red bars indicate significant results.

(A) IBS -> Neuroticism

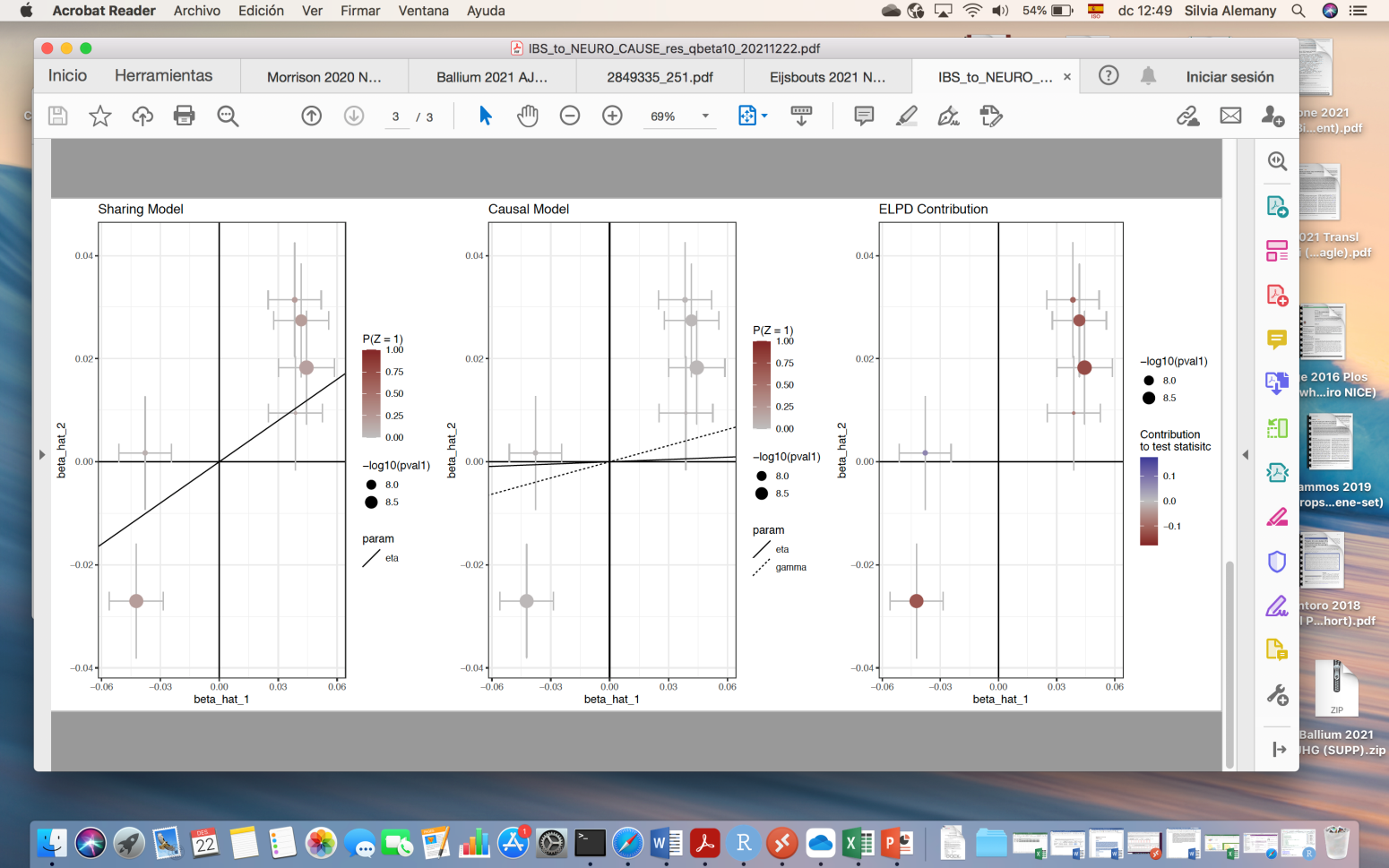

(B) Neuroticism -> IBS

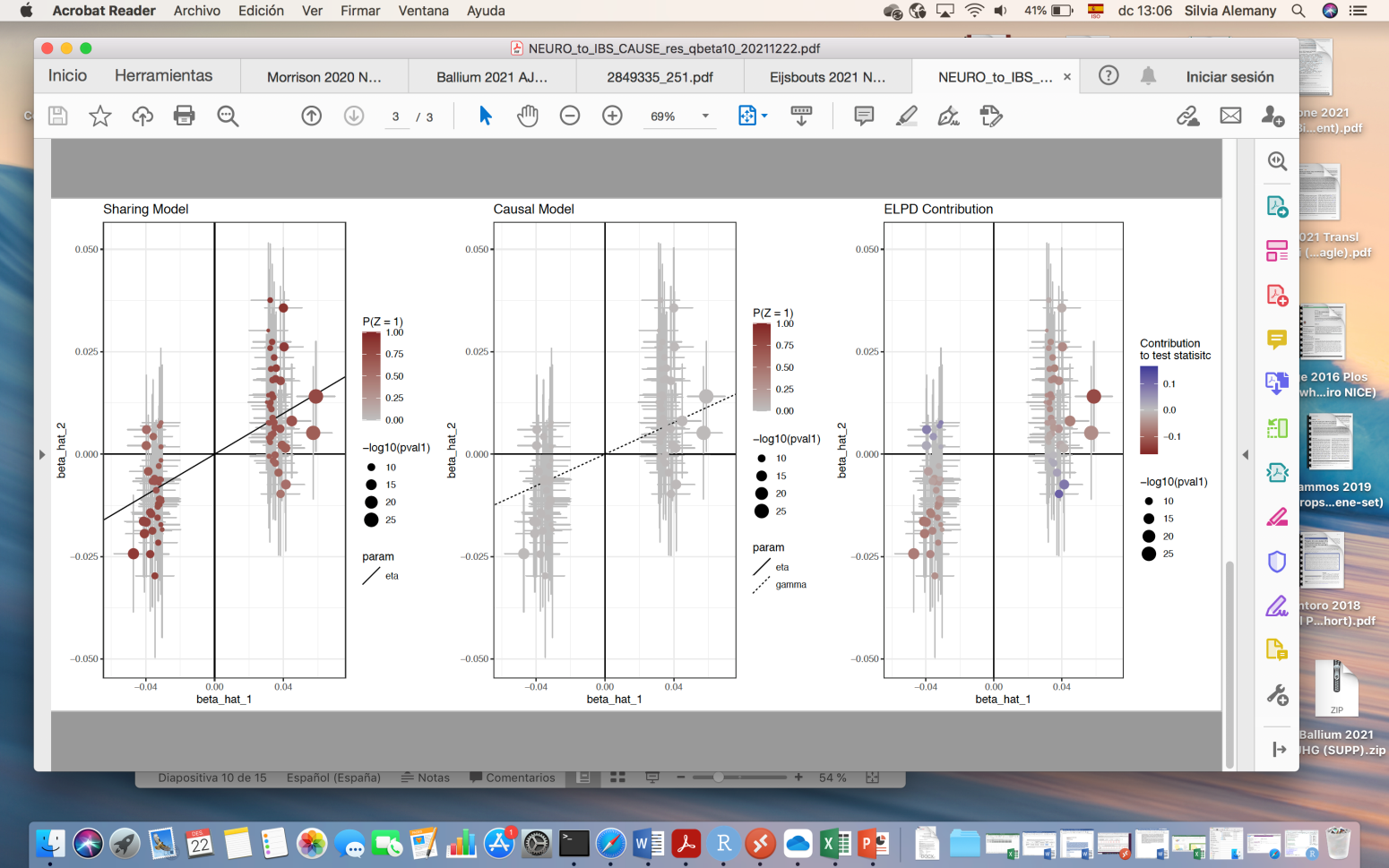

(B)
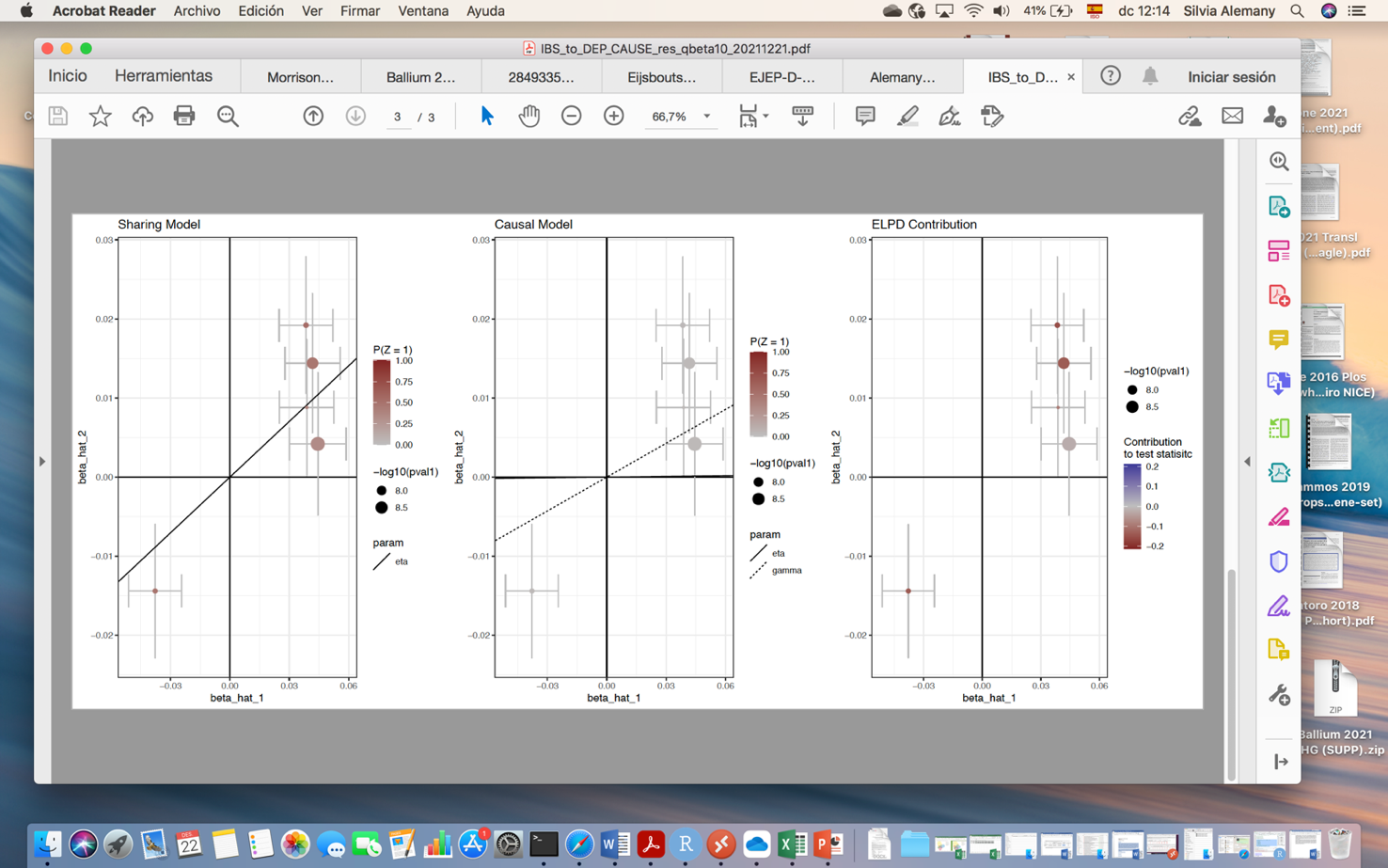
IBS-> Depression

(D) Depression -> IBS

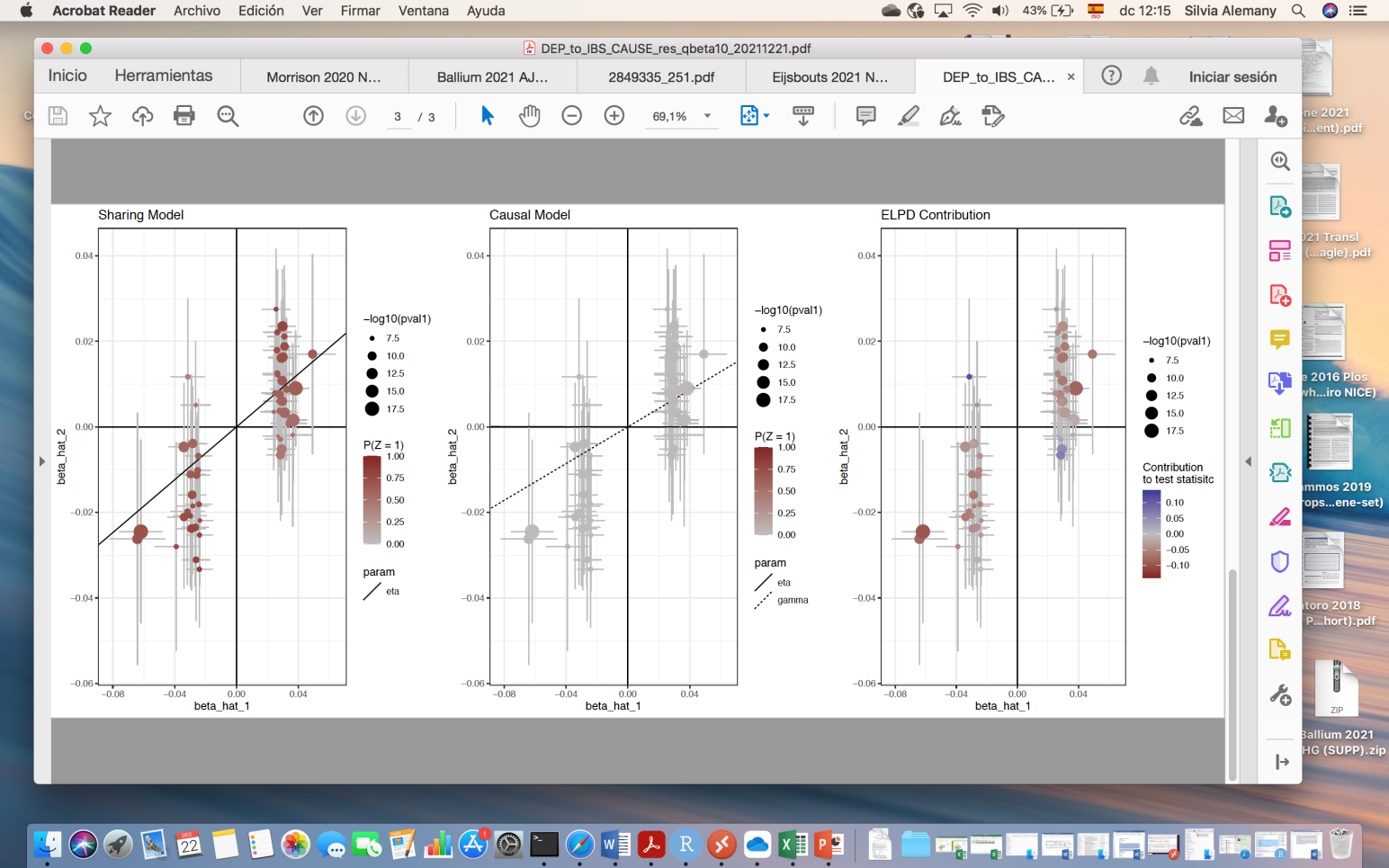

(E) IBS-> Anxiety

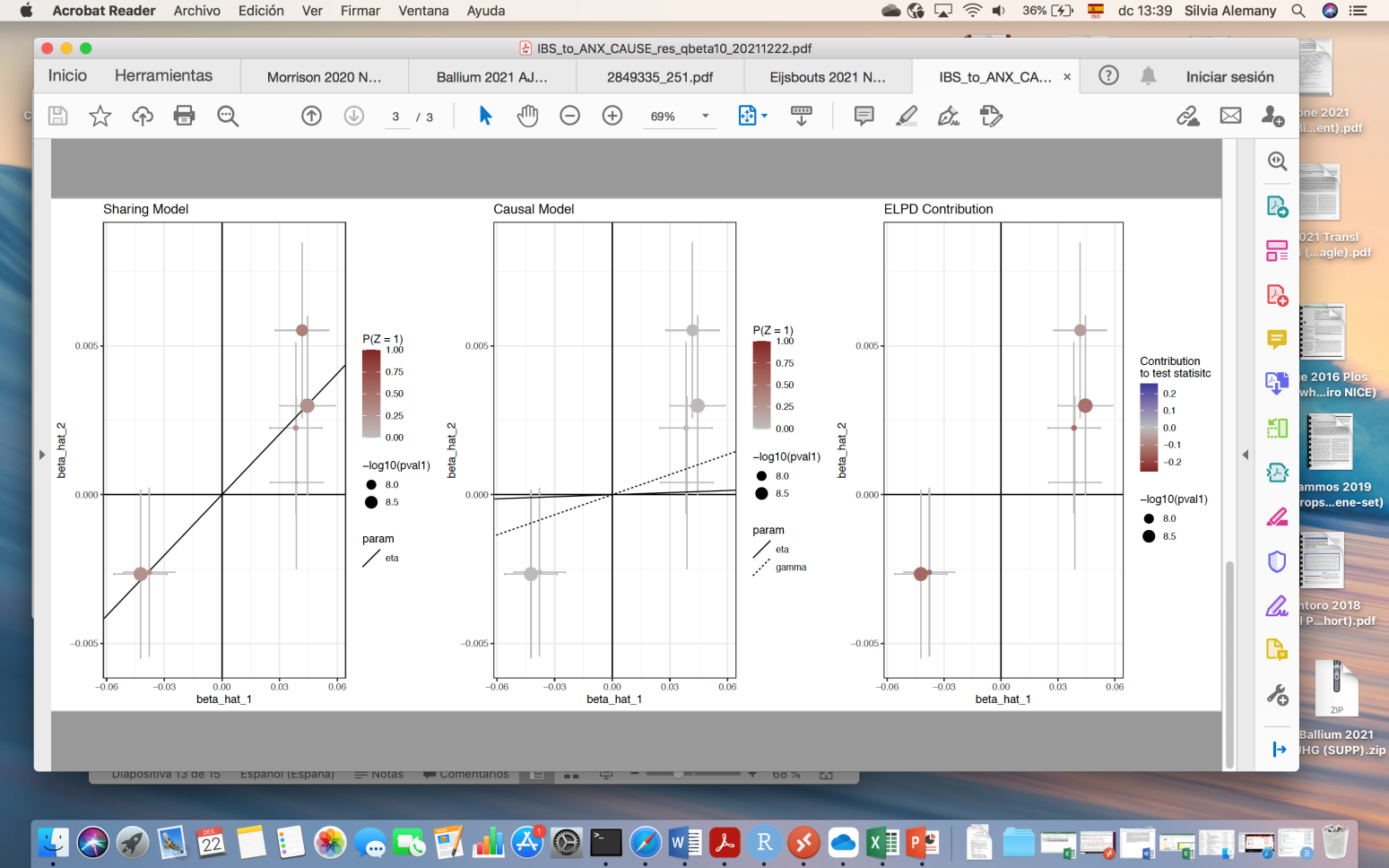

(F) Anxiety -> IBS

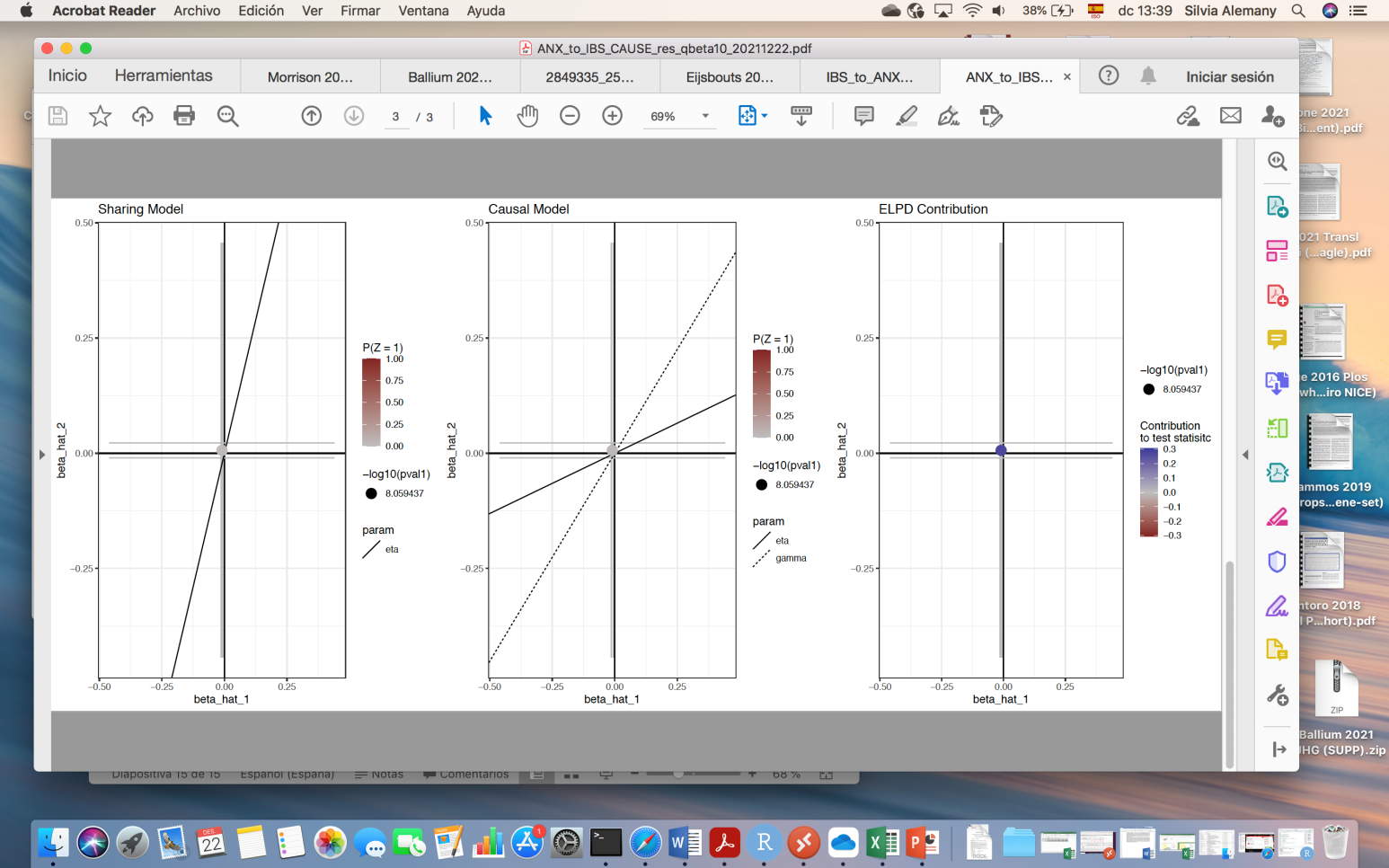

### **Figure S6. Scatter plots of the causal analysis.**

Scatter plots of exposure versus outcome effect sizes for: the sharing model (left) illustrating the pattern induced by a shared factor (correlated pleiotropy, eta) without a causal effect; the causal model (middle) illustrating the pattern induced when including also a causal effect (gamma); and the expected log pointwise posterior density (DEPLD) contribution from each variant for each causal relationship tested.
